# A New Vaccination Assessment Method and Strategy for COVID-19

**DOI:** 10.1101/2023.04.14.23288579

**Authors:** James Michaelson

**Affiliations:** Department of Pathology, Massachusetts General Hospital, USA; Department of Surgery, Massachusetts General Hospital, USA; Department of Pathology, Harvard Medical School, USA

## Abstract

Optimizing vaccination to reduce **COVID-19** death remains a challenge. A new method, ***Gompertzian Analysis***, examines numbers of infectious disease cases and deaths, by age, on log graphs, capturing **COVID-19 *Lethality*** (Deaths/Cases) by the ***Gompertz Mortality Equation***. ***Gompertzian Analysis*** revealed that each of the first 4 ***Vaccination Events*** (primary and boosters) led to a ∼1/3^rd^ reduction in **COVID-19 *Lethality***. These vaccination reductions in **COVID-19 *Lethality*** were cumulative, persistent, and undiminished by variants, while vaccination’s impact on **COVID-19 *Infectivity*** (Cases/Population) was fleeting. Primary vaccination and 3 boosters gave an ∼85% reduction in **COVID-19 *Lethality***, with projections suggesting ∼68% fewer deaths (∼267,000 to ∼85,000). Projections also suggest that 6 boosters may offer a ∼96% reduction in **COVID-19 *Lethality***, to the familiar level of influenza, with a ∼91% reduction in **COVID-19** deaths, to ∼25,000, fewer than automobile deaths. ***Gompertzian Analysis*** provides rational vaccination guidelines by age. ***Gompertzian Analysis*** points to a strategy molded by multiple vaccinations reducing **COVID-19 *Lethality***, which is persistent, rather than focusing on reducing **COVID-19 *Infectivity***, which is fleeting. Such a strategy, based on accumulating the necessary number of vaccinations (∼7), and possibly no more, would accept vaccination’s limited ability to prevent infection in exchange for its power to prevent death.

## INTRODUCTION

How might **COVID-19** be brought under control? Here, a new method, ***Gompertzian Analysis***, is outlined, which uses data on the numbers of infectious disease cases and deaths to reveal hidden insights into how vaccines can be more effectively utilized to save many more lives.

## METHODS

Let us call the examination of numbers of infectious disease cases and deaths on log graphs, ***Gompertzian Analysis***, taking advantage of the ∼10,000-fold exponential increase in the risk of **COVID-19 *Lethality*** (Deaths/Cases) from infancy to old age (**FIGURE 1**). This relationship between **COVID-19 *Lethality*** (***D***) and age (***t***) can be captured mathematically by the ***Gompertz Mortality Equation***, and visualized graphically by sorting patients by age, and then examining deaths and cases on log graphs, yielding straight rows of data points, called here ***Gompertz Lines*** (**FIGURE 1**). Values for the two parameters of the ***Gompertz Mortality Equation***, the ***Gompertzian Slope, G***_***s***_, and the ***Gompertzian Height***”, ***G***_***H***_, emerge by fitting the data to the exponential equation.

**FIGURE 1.**
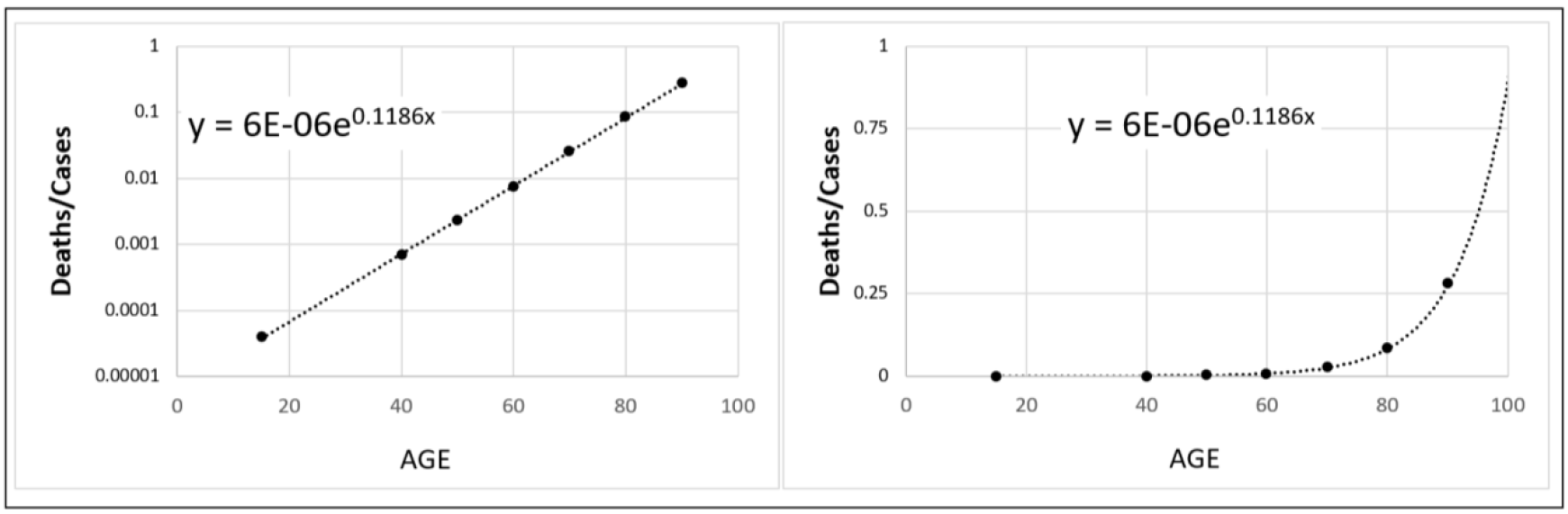
**COVID-19 *Lethality*** (Deaths/Cases) in the Pre-Vaccination era, visualizes the ***Gompertz Mortality Equation: log*(*D*)** = (***G*_*S*_ · *t***) **+ log (*G*_*H*_**), which is equivalent to 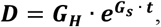, for the relationship between **COVID-19 *Lethality, D***, the risk of death if infected, that is, (Deaths/Cases), vs age, ***t***, captured by the parameters of the ***Gompertzian Slope, G***_***s***_ (the rate of increase with age), and ***Gompertzian Height, G***_***H***_ (the location of the ***Gompertz Line*** on the log graph, up or down).^3^ For values from various datasets, see the APPENDIX.

As we shall see here, and in detail in the **APPENDIX**, ***Gompertzian Analysis*** of many datasets has revealed that vaccination, and other forces, do not appear to make a marked impact on the ***Gompertzian Slope, G***_***s***_, of the ***Gompertz Mortality Equation***, of **COVID-19 *Lethality*** (Deaths/Cases), giving us a simple single number, ***G***_***H***_, the ***Gompertzian Height***, with which we can examine, predict, and test, the combined impacts of morbidity factors, ***Vaccination Events***, infections, and age, into a single measure of **COVID-19 *Lethality***. The cellular mechanism behind this exponential quality of mortality is examined in the companion manuscript to this communication^1^.

The results of ***Gompertzian Analysis*** described were from publicly available datasets, which can be found in the references here, and in the **APPENDIX**. For the terminology used here, see the **APPENDIX**.

## RESULTS AND DISCUSSION

### COVID-19’s *Gompertzian Force of Mortality*

The exponential relationship between age and risk of death, discovered for **All-Cause *Mortality*** by Gompertz in 1825^2^ and for **COVID-19 *Lethality*** (Deaths/Cases) by Levin and colleagues in 2020,^3^ and confirmed by others,^4-9^ is captured by the ***Gompertz Mortality Equation***, appearing as straight ***Gompertz Lines*** on log graphs (**FIGURE 1**, left). On linear graphs, they appear as exponential curves, whose values from the pre-vaccination era reveal **COVID-19**’s shocking age bias, with individuals stealthily accumulating less than 1% chance of death from infancy to age 60, followed by the disease’s progressively more catastrophic, exponential, increase in **COVID-19 *lethality*** rising to about 3% at age 70, to almost 10% at 80, to about 25% at 90, and to about 80% at 100.

### Each ***Vaccination Event*** decreased **COVID-19 *Lethality*** (Deaths/Cases) by about 1/3^rd^

In **FIGURE 2** are the values for **COVID-19 *Lethality*** (Deaths/Cases), by age, for groups of Israeli patients: “unvaccinated”; or “Only primary vaccination”; or “primary vaccination +1 booster”; or “primary vaccination +2 boosters”.^10^ For each ***Vaccination Event***, the ***Gompertz Line***, and thus ***Gompertzian Height, G***_***H***_, of **COVID-19 *Lethality*** (Deaths/Cases), declined cumulatively (**FIGURE 2, APPENDIX FIGURES** 23-41, **APPENDIX-TABLE** I). Patients who had had only primary vaccination received a ∼36% reduction in **COVID-19 *Lethality***, captured by ***G***_***H***_, relative to the unvaccinated (+/-0.4%). Those with the first booster received yet an additional ∼36% reduction (+/-0.2%). Those with a second booster received one more additional reduction of ∼32%, for a total accumulated reduction in **COVID-19 *Lethality*** (Deaths/Cases), across all ages, of ∼75% (+/-0.6%).

**FIGURE 2.**
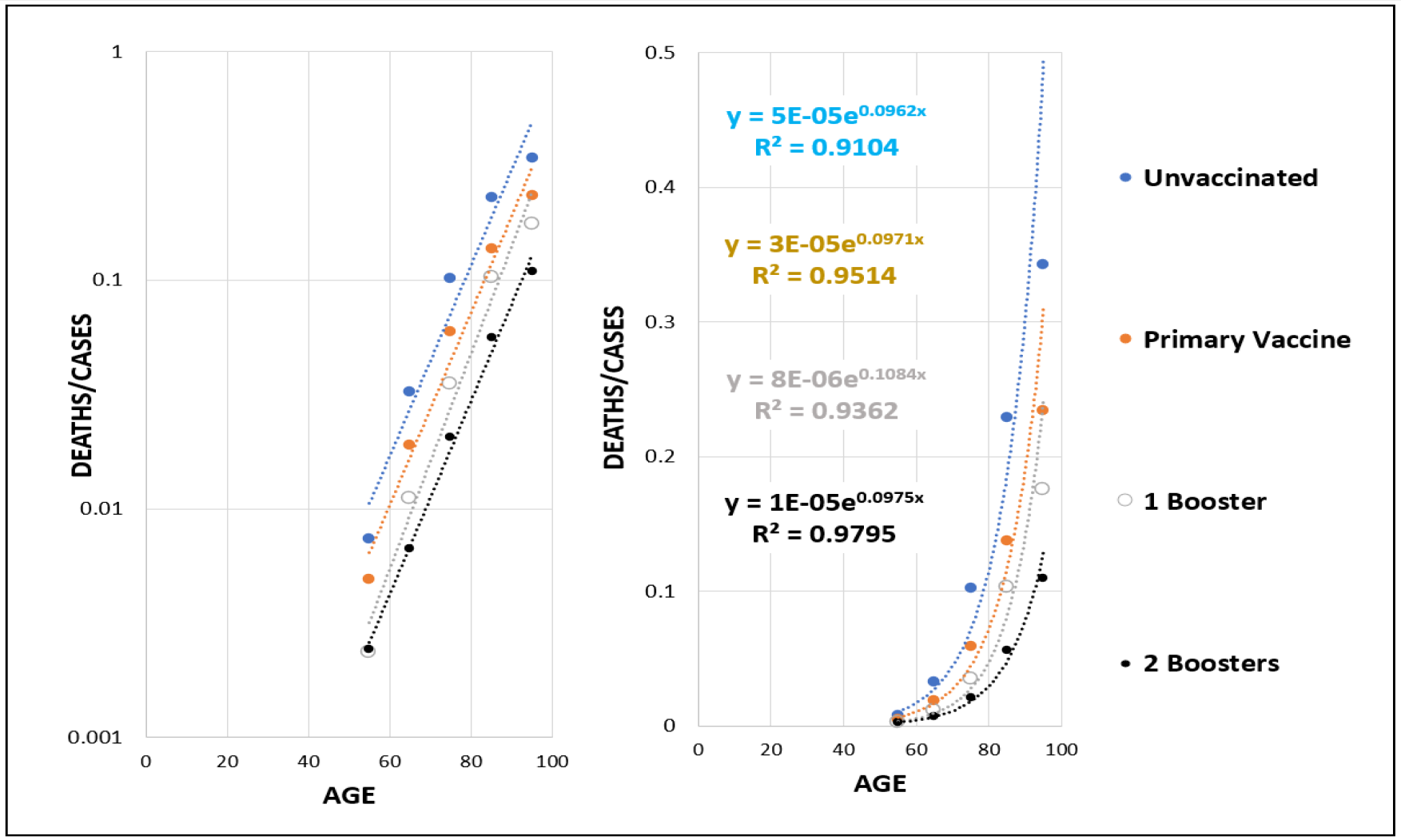
**COVID-19 *Lethality*** (Deaths/Cases), by age, for groups of Israeli patients who have chosen one of four opportunities for vaccination: those who are “unvaccinated”; those who have had “Only primary vaccination”; or “primary vaccination +1 booster”; or “primary vaccination +2 boosters. This dataset excludes patients who have had infections, thus containing the impact of only vaccination on **COVID-19 *Lethality***.

Similar datasets from multiple countries show similar declines in **COVID-19 *Lethality*** (Deaths/Cases) with each ***Vaccination Event*** (See **APPENDIX**). This can be seen in data from Hungary^11^ (**APPENDIX FIGURES** 40-41), and from the USA, for 65 million patients, March 2022-July 2022.^12^ (**APPENDIX FIGURES** 33-39). Recently, data from the USA show a cumulative decline for all 3 boosters, the last of which was the ***Bivalent*** formulation (**APPENDIX-FIGURES-**41 and 42).^13^ Subtle differences between these various datasets are presented and discussed in the **APPENDIX**. Examination of the underlying cellular basis of **COVID-19 *Lethality***, the mathematics for which is outlined in the companion communication to this manuscript,^1^ also suggests that the cellular changes induced by each sequential ***Vaccination Event*** would be expected to lead to a roughly equivalent, and additive, reduction in the ***Gompertzian Height, G***_***H***_, of the ***Gompertz Mortality Equation*** (Michaelson and Huang, unpublished).

### **COVID-19 *Lethality*** and ***Infectivity***: 2021-2022

In **FIGURE 3** are shown values provided by the CDC for approximately 70% of all **COVID-19** cases in the USA for each week from April 2021 to June 2022, for both the vaccinated and unvaccinated, for individuals of age 50-64.^12^ Similar images for the other age groups can be seen in **APPENDIX-FIGURE** 19. This information makes it possible to chart the evolution of **COVID-19 *Infectivity*** (Cases/Population) and **COVID-19 *Lethality*** (Deaths/Cases), the features of which we shall examine in the next two sections.

**FIGURE 3.**
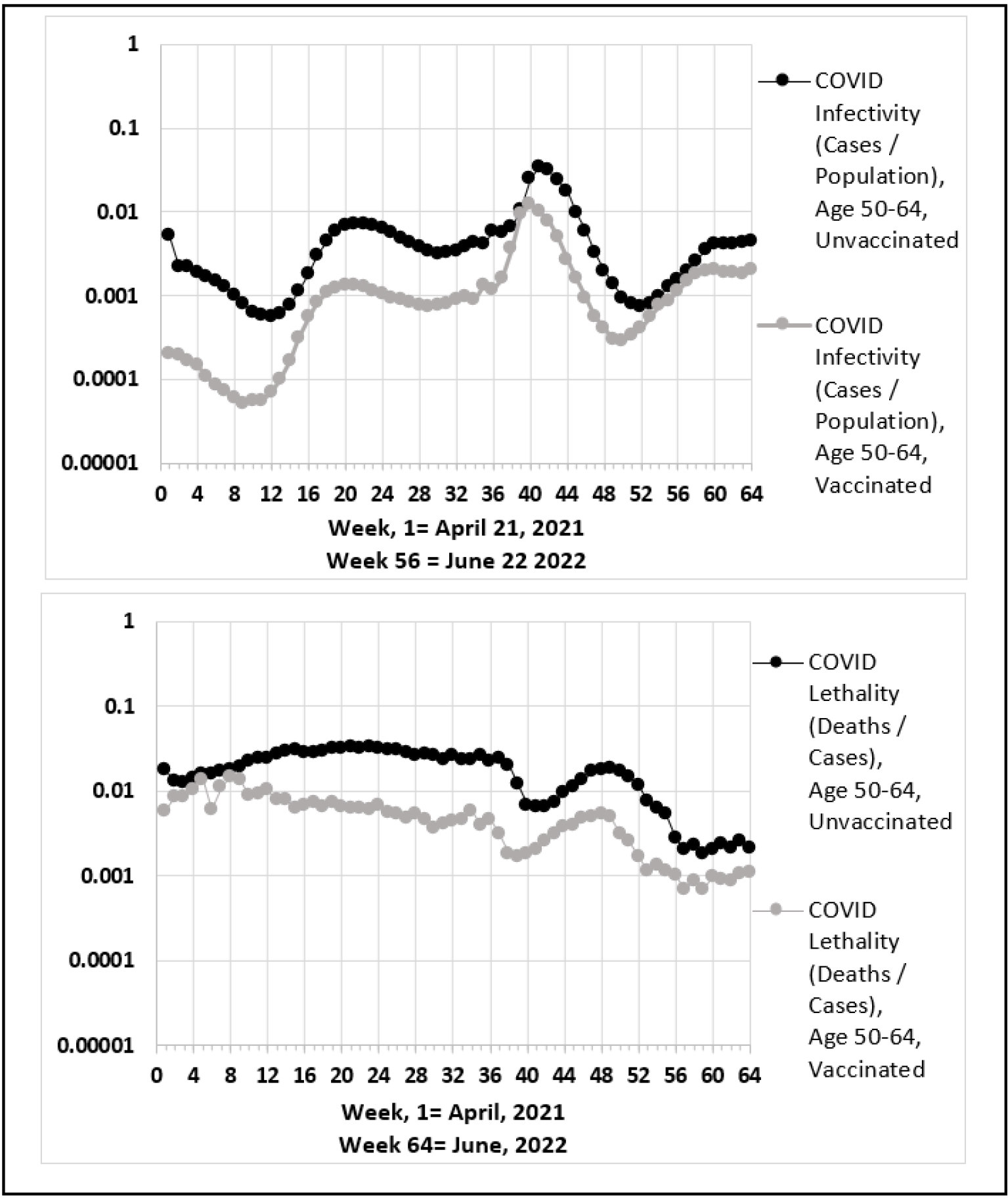
**COVID-19 *Infectivity*** (Cases/Population) and **COVID-19 *Lethality*** (Deaths/Cases), USA April 2021 to June 2022, vaccinated and unvaccinated, age 50-64.^12^

### **COVID-19 *Lethality*** is persistent

As can be seen in **FIGURE 3** (and **APPENDIX-FIGURE** 19), not only was there no sign of fading of vaccine-induced reduction in **COVID-19 *Lethality*** (Deaths/Cases) over the 15-months from April 2021 to June 2022, **COVID-19 *Lethality*** continued to decline, no doubt, by boosters and breakthrough infections. For the unvaccinated, **COVID-19 *Lethality*** (Deaths/Cases) also declined, no doubt, by immunization by infection, at the sad price of lives lost (**APPENDIX-FIGURE** 18, **APPENDIX-FIGURE** 22).

For both the vaccinated and the unvaccinated, **COVID-19 *Lethality*** went up and down mildly over the 15 months, with small waves that varied several-fold in height. However, over the long term, **COVID-19 *Lethality*** declined roughly 10-fold from April 2021 to June 2022. These data provide us with a picture of vaccination’s impact on **COVID-19 *Lethality*** (Deaths/Cases) being persistent and cumulative, with no sign of waning with time.

The persistence of **COVID-19 *Lethality*** (Deaths/Cases), and its cumulative reduction by vaccination, appears again and again in the data. Indeed, I have yet to find a convincing example of fading of **COVID-19 *Lethality***. These many examples can be seen throughout the **APPENDIX**.

### **COVID-19 *Infectivity*** is fleeting

While vaccinated individuals had lower levels of **COVID-19 *Infectivity*** (Cases/Population) than unvaccinated, over the 15-month period, this faded, with vaccinated individuals twice reaching the same level of infection as unvaccinated individuals (**FIGURE 3, APPENDIX-FIGURE** 19). Considerable additional data, from a great variety of populations and datasets, support this image, as can be found in exhaustive detail in the **APPENDIX**.

For both vaccinated and unvaccinated individuals, **COVID-19 *Infectivity*** (Cases/Population) went up and down wildly with time, with the huge waves, varying over a hundred+-fold difference in height, and displaying, over the long term, a roughly a 10-fold increase in the chance of infection from April 2021 to June 2022 (**FIGURE 3, APPENDIX-FIGURE** 19). Vaccinated people had lower rates of **COVID-19 *Infectivity*** (Cases/Population) than unvaccinated people, but the virus kept defeating the vaccines. These data provide us with a picture of vaccination’s impact on **COVID-19 *Infectivity*** as being fleeting, eventually waning with time.

The fleeting nature of **COVID-19 *Infectivity*** (Cases/Population), despite its temporary reduction by vaccination, however welcome, appears again and again in the data. These many examples can be seen throughout the **APPENDIX**.

### What can be expected from sequential vaccinations: **COVID-19 *Lethality*** to negligible levels

What can be expected from sequential ***Vaccination Opportunities*** ? As we have seen above, for each of the first four ***Vaccination Opportunities*** (primary vaccination, 1^st^ booster, 2^nd^ booster, and 3^rd^ booster) there have been cumulative reductions in **COVID-19 *Lethality*** (Deaths/Cases). The ***Gompertz Mortality Equations*** for each of these ***Vaccination Events***, and their observed reductions in **COVID-19 *Lethality*** (Deaths/Cases) are shown in **FIGURES 4** and **5** and **TABLE** 1. These values show, across the ages, that primary vaccination leads to a ∼36% reduction in **COVID-19 *Lethality***, in comparison to the unvaccinated. Primary vaccination plus 1 booster results in a ∼65% reduction, in comparison to the unvaccinated. Primary vaccination plus 2 boosters give an ∼75% reduction, while primary vaccination plus 3 boosters was likely to have led to an ∼85% reduction. (The precise value of this last reduction awaits the release of data on patients sorted by the number of ***Vaccination Events*** up to the bivalent booster: see **APPENDIX** for details).

**FIGURE 4.**
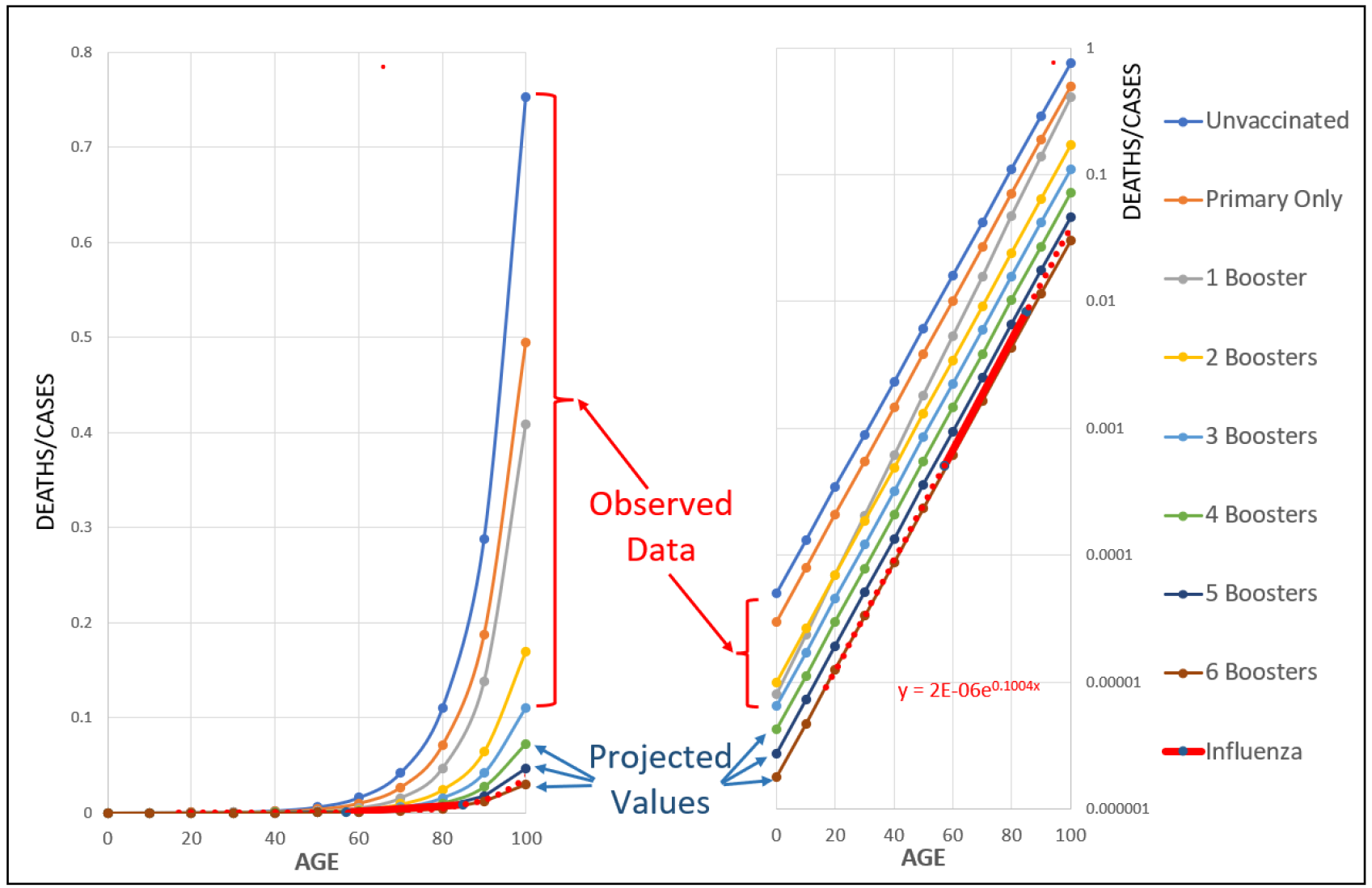
Observed and expected cumulative declines **COVID-19 *Lethality*** (Deaths/Cases) with each ***Vaccination Event***, as captured by the ***Gompertz Mortality Equation***. ∼ 1/3^rd^ reductions for each ***Vaccination Event*** have been observed for the first 3 ***Vaccination Events*** (**FIGURE 2**, and other data: see **APPENDIX**). The precise value of the 4^th^ ***Vaccination Event*** (Bivalent booster) has yet to be measured precisely, due to incomplete data, but appears to approximately this value.^13^ Also shown is the ***Gompertz Line*** for patients with influenza.^16^

**FIGURE 5.**
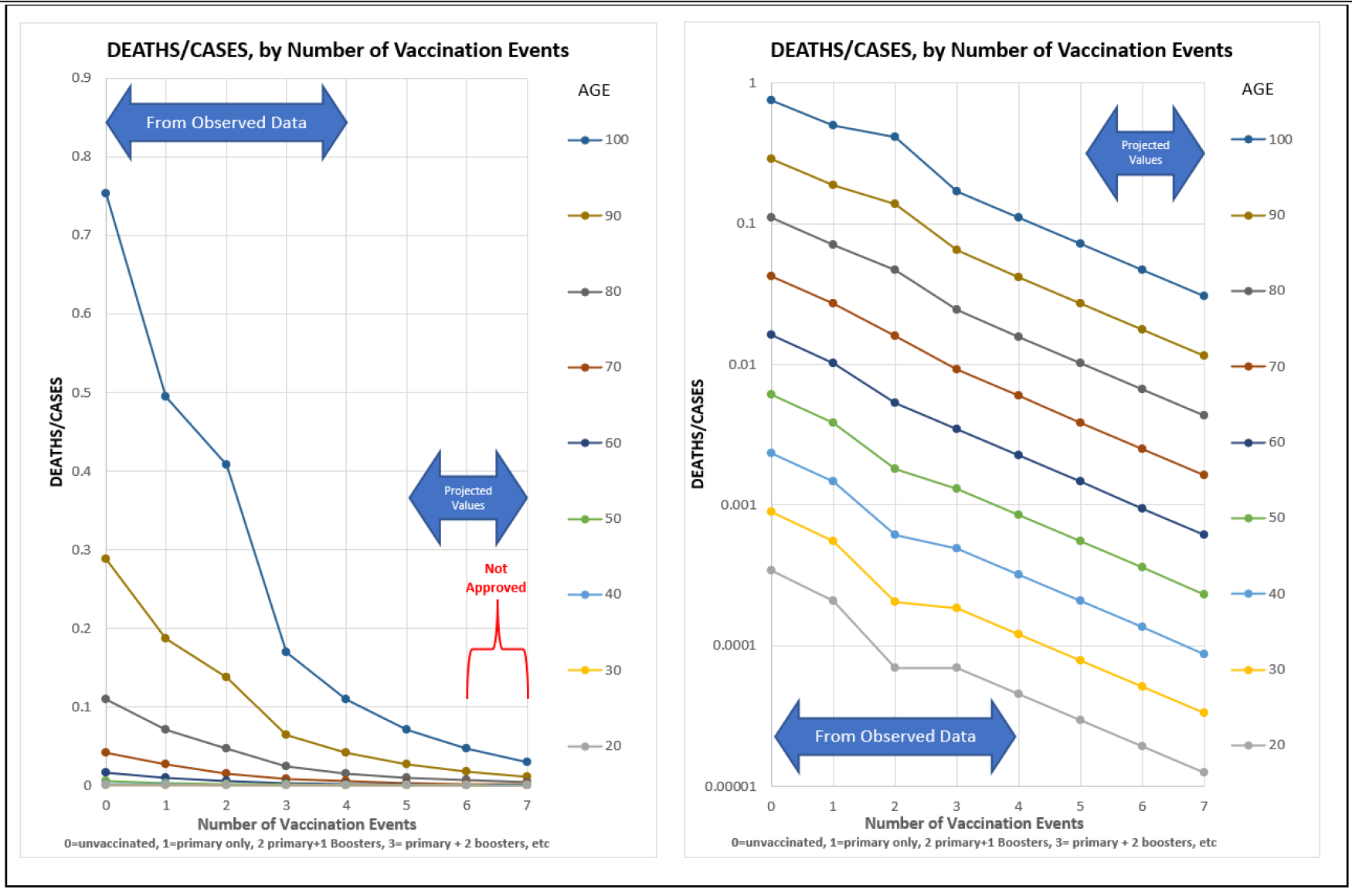
Observed and expected cumulative declines **COVID-19 *Lethality*** (Deaths/Cases), graphed by the number of ***Vaccination Events***. See **FIGURE 4** for details.

### The number of lives that can be saved by higher utilization of vaccination

How do these **COVID-19 *Lethality*** (Deaths/Cases) values (**FIGURES 4** and **5, TABLE 1**) translate into lives saved? In 2022, 13% of Americans had had no **COVID-19** vaccination, 31% were vaccinated without boosters, 39% had one booster, 17% had two boosters,^12^ and there were ∼267,000 **COVID-19** deaths.^14^ The **COVID-19 *Lethality*** values described above, derived by ***Gompertzian Analysis*** from actual data, make it possible to calculate how many deaths would have been prevented, and can be prevented in the future, with greater use of vaccination (**FIGURE 6, APPENDIX-FIGURE-**46, and **APPENDIX TABLE** V). The practical execution of these calculations is straightforward; details can be found in the **APPENDIX** (**APPENDIX-TABLES V** and **VI**).

**FIGURE 6.**
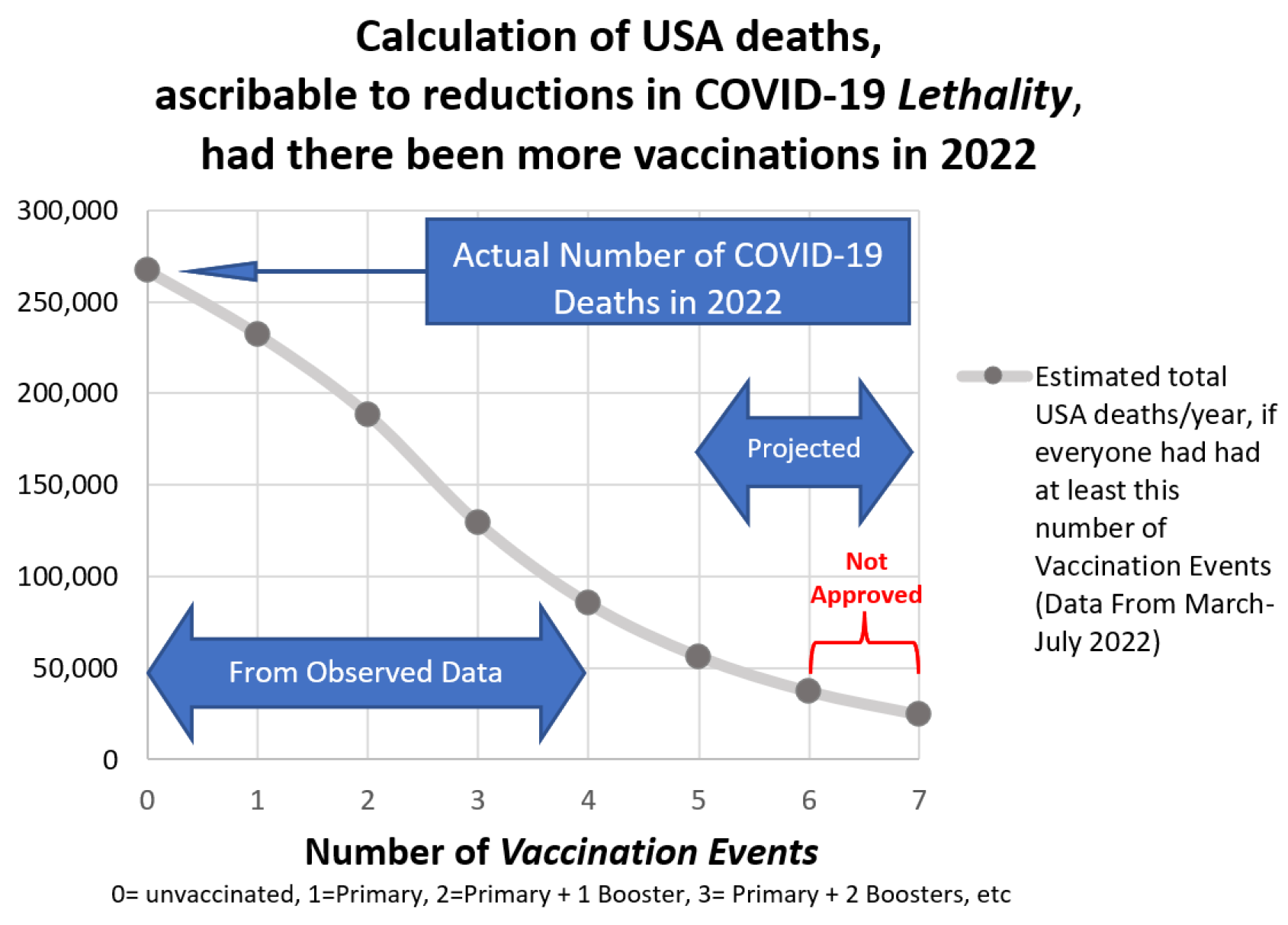
Calculation of USA deaths, ascribable to reductions in **COVID-19** Lethality, had there been more vaccinations in 2022.

**TABLE 1.** Deaths/Million Infections (Values taken from FIGURE 4)

| **TABLE 1. Deaths/Million Infections (Values taken from FIGURE 4)** |  |  |  |  |  |  |  |  |  |  |
| --- | --- | --- | --- | --- | --- | --- | --- | --- | --- | --- |
| Age-> | 10 | 20 | 30 | 40 | 50 | 60 | 70 | 80 | 90 | 100 |
| Unvaccinated | 131 | 342 | 896 | 2,345 | 6,137 | 16,059 | 42,025 | 109,977 | 287,801 | 753,152 |
| Primary Only | 79 | 209 | 552 | 1,459 | 3,851 | 10,170 | 26,855 | 70,912 | 187,250 | 494,448 |
| 1<sup>st</sup> booster | 24 | 70 | 207 | 611 | 1,807 | 5,342 | 15,795 | 46,697 | 138,060 | 408,171 |
| 2<sup>nd</sup> booster | 26 | 70 | 186 | 492 | 1,303 | 3,452 | 9,142 | 24,212 | 64,125 | 169,835 |
| 3<sup>rd</sup> booster | 17 | 46 | 121 | 320 | 847 | 2,244 | 5,942 | 15,738 | 41,681 | 110,393 |

Even without taking into account vaccination’s reduction in infection, that is, reduction in **COVID-19 *Infectivity*** (Cases/Population), had all Americans utilized at least primary vaccination, the reduction in **COVID-19 *Lethality*** (Deaths/Cases) would have resulted in the ∼267,000 **COVID-19** deaths that occurred in 2022 being cut by ∼13% to ∼232,00 deaths (**FIGURE 6**). Had all Americans utilized at least primary vaccination and 1 booster, then **COVID-19** deaths that occurred in 2022 would have been cut by ∼29% to ∼189,00. Had all Americans utilized at least primary vaccination and 2 boosters, then death would have been cut by ∼52% to ∼129,00. Had all Americans utilized primary vaccination and 3 boosters, then lives lost would likely have been cut by ∼68% to ∼85,00 deaths.

### ***Gompertzian Analysis*** gives us rational vaccination guidelines for people of all ages

The findings of ***Gompertzian Analysis*** (**FIGURE 4**) make it possible to create rational, data-driven, age-specific vaccination guidelines for reducing the risk of **COVID-19** death, as Offit has suggested.^15^ For example, most Americans accept the risk of driving: ∼120 automotive deaths/million-Americans/year. Should this risk level define our choices for vaccination, then primary vaccination and 1^st^ booster would be wise if ∼age 20 or older, and the 2^nd^ and 3^rd^ boosters if ∼age 40 or older (**TABLE 2**). Not only would such a guideline allocate vaccines to those who would benefit, it would also save more than 99% of the lives that would be saved had people of every age been vaccinated. For a deeper look at such calculations, see **APPENDIX**.

**TABLE 2.** Deaths Prevented, by Last ***Vaccination Event***/Million Infections; Recommended Vaccinations Highlighted

|  |  |  |  |  |  |  |  |  |  |  |
| --- | --- | --- | --- | --- | --- | --- | --- | --- | --- | --- |
| Age-> | 10 | 20 | 30 | 40 | 50 | 60 | 70 | 80 | 90 | 100 |
| Primary Only | 52 | 133 | 344 | 886 | 2,285 | 5,889 | 15,170 | 39,064 | 100,551 | 258,704 |
| 1<sup>st</sup> booster | 56 | 139 | 346 | 847 | 2,044 | 4,828 | 11,060 | 24,215 | 49,190 | 86,277 |
| 2<sup>nd</sup> booster | 0 | 0 | 21 | 119 | 504 | 1,891 | 6,653 | 22,486 | 73,935 | 238,336 |
| 3<sup>rd</sup> booster | 9 | 25 | 65 | 172 | 456 | 1,208 | 3,200 | 8,474 | 22,444 | 59,442 |

### Future ***Vaccination Opportunities***

An additional ***Vaccination Opportunity*** has been approved by the FDA for the Fall of 2023. One might wonder what impact that this vaccination, and others in the future, could have on **COVID-19** outcome. We can project what might be the result, if each of these leads to the 1/3^rd^ reduction in **COVID-19 *Lethality*** that occurred for the previous ***Vaccination Opportunities***. As noted above, such a possibility is biologically and mathematically the most likely expectation of future impacts of vaccination, and is also soundly based on previous experience, but, as of yet, we lack data to know with certainty what the future might bring. Nonetheless, whether such projected reductions in fact repeat as more ***Vaccination Opportunities*** are made available can be known rapidly by using ***Gompertzian Analysis*** to monitor cases and deaths, and appropriate adjustments in policy can be made as we move forward.

In **FIGURES 4** and **5**, the ***Gompertz Mortality Equations*** for 4^th^, 5^th^ and 6^th^ boosters are shown, based on the possibility of additional 1/3^rd^ reductions in **COVID-19 *Lethality*** for each booster. These projected values suggest that 4 boosters might offer the possibility of a ∼91% reduction in **COVID-19 *Lethality***, in comparison to the unvaccinated, while the calculation for 5 boosters yields a value of a ∼94% reduction, and 6 boosters a ∼96% reduction.

The ***Gompertz Mortality Equation*** for influenza^16^ is also shown in **FIGURES 4** and **5**. Influenza’s ***Lethality*** (Deaths/Cases) lies essentially where the calculated **COVID-19 *Lethality*** (Deaths/Cases) would roughly be after 7 ***Vaccination Events***, that is, after a 6^th^ booster. **COVID-19 *Lethality*** would be slightly lower than Influenza’s ***Lethality*** with just one more booster. This raises the possibility that additional ***Vaccination Opportunities*** might bring the lethal burden of **COVID-19** down to, or even below, the familiar level we see for influenza.

These projected values can also be comprehended in terms of lives that might have been saved if more vaccinations had been utilized. As shown in **FIGURE 6**, these calculations suggest that a total of 6 boosters yields a value of a ∼91% reduction in lives lost, to ∼25,000 deaths, far fewer lives than are lost each year in automobile accidents.

These possibilities appear to merit examination by trial, whose outcomes could be assessed by ***Gompertzian Analysis. Gompertzian Analysis*** could also provide the basis for constructing rational vaccination guidelines by age, as described above, for allocating additional vaccines for optimal effect, as well as for ongoing monitoring of a strategy based on this approach, should it be put into practice.

### Once a finite number of ***Vaccination Events*** are achieved (7?), no more may be required

***Gompertzian Analysis*** suggests that once we avail ourselves of a finite number of ***Vaccination Opportunities***, perhaps 7, we may need no more for the rest of our lives. The reduction in death, potentially down to levels lower than are caused by automobile accidents (**FIGURE 6**), and the reduction in the risk of death once infected, potentially down to a level lower than that for influenza (**FIGURES 4** and **5**), may well be sustained permanently, without ever requiring any more vaccinations.

While vaccination’s reduction in **COVID-19 *Infectivity*** (Cases/Population) has proven to be short lived, it’s reduction in **COVID-19 *Lethality*** (Deaths/Cases) has been persistent, as we have seen in **FIGURE 3** and in many other examples outlined in the **APPENDIX**. **SARS-CoV-2** infection may well continue, but by accumulating enough ***Vaccination Events***, possibly primary vaccination followed by 6 boosters, and then no more, the virus’s ability to cause death may be permanently reduced to levels vastly lower than what we see now.

Of course, continued monitoring will be essential to assess whether we ever do reach such a permanent state of negligible levels of **COVID-19** death. However, the prospect of such a favorable future must be borne in mind as a possibility that may be achievable. In short, ***Gompertzian Analysis*** of the data seem to be telling us that with a few more vaccinations now, and hopefully no more vaccinations in the future, we may well bring **COVID-19’**s loss of life under control.

### The Cellular Mechanisms Behind **COVID-19 *Lethality*** and ***Infectivity***

Why do vaccines reduce the chance of **COVID-19 *Infectivity*** (Cases/Population)? Why do they reduce the chance of **COVID-19 *Lethality*** (Deaths/Cases), pushing down the ***Gompertzian Height, G***_***H***_, of the exponential ***Gompertz Mortality Equation*** ? Behind these macroscopic manifestations of **COVID-19** infection, death, and prevention, lie the microscopic worlds of cells. In the companion manuscript to this communication, ^1^ my colleagues and I have outlined how, by examining growth and development in units of numbers of cells, ***N***,^17-24^ an approach we have called ***Cellular Phylodynamic Analysis***^23^, ***the Gompertz Mortality Equation***, affecting both aging’s impact on mortality generally, and **COVID-19 *Lethality*** specifically, can be traced to the slowing of growth that marks maturity. A likely explanation for this finding is that cell division makes us grow, but it also keeps us well, since biological materials decay with time, a toxic problem that is reversed when cells divide and replace at least half of their components anew.^25,26^ For the purposes of the issues examined in this report, this means that changes in numbers of cells as we age should allow us to trace the origin of **COVID-19 *Lethality*** (Deaths/Cases) that marks old age to the loss of specific cells of the immune system. Counting these cells, in individuals who have had boosters, could estimate vaccination’s impact on the decline in **COVID-19 *Lethality***, without having to rely on death data. Here, I have spared the reader this mathematics, but it can be found in this manuscript’s companions communication,^1^ and in abundance in the **APPENDIX** to that communication.

The same ***Cellular Phylodynamic*** math that could trace age-related **COVID-19 *Lethality*** to the cells in the immune system that fail as we age can also be used to track the cells in the other side of the immune system, which creates the reduction in **COVID-19 *Infectivity*** (Cases/Population) that prevents infection. Such math could be used to examine numbers of immune system cells making antibodies, and thus be put to work optimizing vaccine implementation (dose, schedule, adjuvant, etc.). Again, the methods for generating the math for accomplishing this task can be found in Reference 1.

### Different vaccines have different reductions in **COVID-19 *Infectivity*** and ***lethality***

***Gompertzian Analysis*** of a variety of datasets has found that different vaccines have different reductions in **COVID-19 *Lethality*** (Deaths/Cases) and **COVID-19 *Infectivity*** (Cases/Population) (**APPENDIX TABLES** I & II, **APPENDIX FIGURE** 12). In one population, vaccines with the highest and lowest reductions in **COVID-19 *Infectivity*** (Cases/Population) differed more than ***5-fold***.^27^ This information could be used to select vaccines with the greatest potential for reducing **COVID-19 *Infectivity*** (Cases/Population) and ***Lethality*** (Deaths/Cases).

In the **APPENDIX**, a third measure, ***Malthusian Lethality*** (Deaths/Population), captures the aggregate outcome of **COVID-19 *Lethality*** (Deaths/Cases) and ***Infectivity*** (Cases/Population). This provides a way to optimize vaccine selection and implementation for the greatest possible reduction in death.

Since ***Gompertzian Analysis*** has measured the degree to which different vaccines lead to different reductions in **COVID-19 *Lethality*** and ***Infectivity***, the possibility of examining, in parallel, the changes in cell numbers in the immune system that could be identified by ***Cellular Phylodynamic*** math noted above, is appealing. Such an approach offers the possibility of ***in vitro*** assays for **COVID-19 *Lethality*** and ***Infectivity***, and their reductions by vaccination, and for **COVID-19 *Infectivity***, the status of its persistence or fading with time.

### The separateness of **COVID-19 *Lethality*** and ***Infectivity***

Key to the ***Gompertzian Analysis*** described here has been the examination of **COVID-19 *Lethality*** (Deaths/Cases) and **COVID-19 *Infectivity*** (Cases/Population), separately. This has not only proved essential from a practical standpoint, it is also reasonable biologically, as infection is the result of the interaction of the virus with the surface of the mucosa, while death is the result of the massive proliferation of the virus throughout the body, as has been made clear by recent autopsy and virological studies. ^28,29^ It has been argued, and admirably supported by laboratory study, that vaccine protection against infection is principally accomplished by **B**-cells, and the antibodies that they make, while vaccine protection against death is principally accomplished by **T**-cells, and the cell-mediated responses that they make.^30^ As noted above, ***Cellular Phylodynamic*** math gives us a chance to track **COVID-19 *Lethality*** and **COVID-19 *Infectivity*** to the **T**- and **B**-cells that affect virus impact and count how they change upon immunization.

### Variants

***Gompertzian Analysis*** of two datasets of patients (UK^31^ and USA^32^) infected by different variants has revealed a lower level of **COVID-19 *Lethality*** (Deaths/Cases), in each age group, for ***delta***, and a lower level yet for ***omicron*** (**APPENDIX-FIGURE**S 48 & 49). In contrast, **COVID-19 *Infectivity*** (Cases/Population) increased with each new variant. Viral Darwinism would be expected to select for variants that become more infectious (**COVID-19 *Infectivity***), but not necessarily more deadly (**COVID-19 *Lethality***), and that is what appears to have been the case so far (**APPENDIX-FIGURE**S 48 & 49).

Not only would it seem likely that **COVID-19 *Infectivity*** and **COVID-19 *Lethality***, and their reductions by vaccination, to be separate things, it would also seem likely that the antigens on the spike protein to which **COVID-19 *Infectivity*** and ***Lethality*** react would be separate as well, with **COVID-19 *Infectivity*** B-Cell epitopes being subject to selection, in contrast to **COVID-19 *Lethality*** T-Cell epitopes, for which there is no reason for selection. Viruses may not care whether they kill or not, but Darwinism provides no selective reward for mutations that just increase the chance of death. This provides a motivation for the laboratory search for these suspected ***Infectivity*** and ***Lethality*** epitopes.

### The Evolution of **COVID-19**

Monto and colleagues have shown that the four ordinary coronaviruses cause mild illnesses, early in life, and continue throughout life, with remarkably high numbers of reinfections.^33^ Of 1004 such infections, occurring in the 3418 individuals studied over an eight - year period, of whom 1378 were followed for 3 or more years, 30% were reinfections, ranging from 1 to 13 per individual. It appears likely that nearly all of these infections were reinfections from earlier points in time. One might hypothesize that the reason why these four ordinary coronaviruses are so harmless is that, from our early childhood onward, we have accumulated, at ages when the ***Gompertz Mortality Equation*** renders these infections innocuous, immunological protection against death for the rest of our lives. In contrast, the reason why **SARS-CoV-2** is so harmful, is that it has arrived to us at ages when the ***Gompertz Mortality Equation*** has rendered it monstrously dangerous, without the immunological protection which childhood infection could have built for our present ages. ***Gompertzian Analysis*** also seems to be telling us that we might be able to create, in adult life, this missed childhood acquisition of resistance to **COVID-19** death by sequential **COVID-19** vaccination. These various notions can be tested, and indeed cry out to be tested, with the highest degree of skepticism. Indeed, all of the first findings of ***Gompertzian Analysis*** described here, and all of the possible actions that these findings suggest, can, and should, be tested, and monitored, by this approach, before any actions might be taken.

### The Future of **COVID-19**

The findings of ***Gompertzian Analysis*** suggest that we will gradually evolve towards resistance to **COVID-19** death by sequential immunization, either by infection, at the sad cost of lives lost, or by vaccination. The data suggest that in the developed world, this appears achievable by more of us getting more injections. For the world as a whole, this might be achieved by creating vaccines that yield a greater reduction in **COVID-19 *Lethality*** with each injection, and, one might hope, with a single injection. In this regard, current interest in creating nasal vaccines, and other advancements, are challenged by the need to combine improved suppression of **COVID-19 *Infectivity*** while maintaining, and hopefully also, strengthening, suppression of **COVID-19 *Lethality***. As Morens, Taubenberger and Fauci have noted, “Apart from preventing initial infection, it is also important to consider the role of host immunity in limiting viral spread once infection has been established … preventing severe disease”^34^.

### ***Gompertzian Analysis*** can aid in the formation of **COVID-19** vaccination policy

An examination of **COVID-19** vaccination policy statements, thoughtfully created by a number of national and international authorities, makes clear that a single approach for identifying which individuals, at which ages, should receive which vaccines, has yet to emerge.^35-42^ ***Gompertzian Analysis*** thus gives a method for molding population-wide policy so as to achieve the greatest possible reduction in **COVID-19** deaths, as well as to create methods for tracking individual’s accumulation of ***Vaccination Events***, to monitor the success of such efforts, and to modify guidance if policy does not achieve the goals expected.

### A Strategy for **COVID-19**

Our vaccines have not been created, assessed, or approved for their capacity to reduce death, but for their capacity to reduce infection. The findings of ***Gompertzian Analysis*** suggest that we might be better off focusing on the achievable goal of reducing deaths rather than focusing on the goal, possibly unattainable, of eliminating infection. ***Gompertzian Analysis*** has already revealed that the data tell us that the use of all of the four approved ***Vaccination Opportunities*** (primary + 3 boosters) by more citizens can cut the number of **COVID-19** deaths by 2/3^rds^. ***Gompertzian Analysis*** has further enticed us with the possibility of pushing the risk of **COVID-19** death to a level lower than that of influenza, and the numbers of **COVID-19** deaths to a level lower than automobile deaths. The key to reaching for such a goal may lie not in getting the latest vaccination, or even the best vaccine for reducing the chance of infection, but in acquiring the necessary number of vaccinations needed to build our resistance to **COVID-19** death. ***Gompertzian Analysis*** provides us with a tool for reaching that goal.

Note: this study has not been peer reviewed, and the findings could change.

## Data Availability

The results described were from publicly available datasets, which can be found in the references here

## Acknowledgments

Many thanks for the excellent advice and suggestions by my colleagues, Michael Lynes, Luke Huang, and Samuel Preston.

