## APPENDIX for "A New Vaccination Assessment Method and Strategy for COVID-19"

James Michaelson PhD,

Department of Pathology, Massachusetts General Hospital, USA
Department of Surgery, Massachusetts General Hospital, USA
Department of Pathology, Harvard Medical School, USA
***For correspondence:**
 (JSM)

### TOC

**OVERVIEW**

**THE GOMPERTZIAN FORCE OF MORTALITY AND GOMPERTZ MORTALITY EQUATION**
- Introduction
- COVID-19's Fearful ***Gompertzian Force of Mortality*** Comes into View When Displayed Linearly
- Early life mortality and **COVID-19 *Lethality*** (Deaths/Cases).

**UNDERLYING CELLULAR BASIS of COVID-19 *Lethality* and *Infectivity*: CELLULAR PHYLODYNAMICS**.

***GOMPERTZIAN ANALYSIS*: PRINCIPLES**
***Gompertzian Analysis***: Introduction
**Terminology: *Vaccination Opportunity*** and ***Vaccination Event***.
***Gompertzian Analysis*** treats resistance to **COVID-19 *Infectivity*** and ***Lethality*** as two separate processes.
Vaccines and other things push up and down ***Gompertz Lines***, defining **COVID-19 *Lethality*** by ***Gompertzian Height***, $G_H$,
***Gompertzian Analysis*** of Other Infectious Diseases
***Gompertzian Analysis*** of Non-Infectious Diseases
***Gompertzian Analysis*** gives population-wide values from partial age samples.
***Gompertzian Analysis*** of Hospitalization.
***Gompertzian Analysis*** with full age information

**THE PROBLEM WE FACE.**

**GOMPERTZIAN ANALYSIS: COVID-19**

***Gompertzian Analysis*** gives 3 new measures: ***Pasteurian Infectivity***, ***Gompertzian Lethality***, ***Malthusian Lethality***.

**Terminology: *Gompertzian Lethality*, COVID-19 *Lethality*, *Pasteurian Infectivity*, COVID-19 *Infectivity*.**

***Gompertzian Analysis*** of **COVID-19 *Infectivity*** and ***Lethality*** in USA 2021-2022, analyzed in aggregate.
- Basic Features, on Graphs.
- Basic Features, in Numbers:3 new measures are each captured by a single number: the ***Gompertzian Height, $G_H$***. ***Gompertzian Lethality*** and ***Case Fatality***.

***Reductions***: 3 additional new measures of ***Gompertzian Analysis*** for measuring impact of vaccination and other forces.
- ***Pasteurian Infectivity Reduction***: The Effect of Vaccines on the Risk of Infection.
- ***Malthusian Lethality Reduction***: The Effect of Vaccines on the Aggregate Risk of Infection and Death Combined
- ***Gompertzian Lethality Reduction***: The Effect of Vaccines on Risk of Death Once Infected

***Gompertzian Analysis*** of the impact of **COVID-19** Vaccination in USA 2021-2022, analyzed in aggregate.
- ***Reductions***: Basic Features, on Graphs.
- ***Reductions***: Basic Features, in Numbers.
  - ***Reductions as Percentages***
  - ***Reductions as Fold Changes***
  - ***Reduction*** in Contrast to ***Vaccine Effectiveness***

***Gompertzian Analysis*** of **COVID-19** Primary Vaccination: VA(USA), Vaccine Impact
***Gompertzian Analysis*** of **COVID-19** Primary Vaccination: VA(USA), Primary Vaccine Comparisons

***Gompertzian Analysis*** of **COVID-19** Primary Vaccination: HUNGARY & ARGENTINA, Vaccine Impact & Comparisons

***Gompertzian Analysis*** of **COVID-19** Primary Vaccination: ISRAEL, Vaccine Impact

***Gompertzian Analysis*** of **COVID-19** Vaccination: Population Comparisons

***Gompertzian Analysis*** of **COVID-19** Vaccination: USA 2021-2022, impact over time.
- ***Gompertzian Analysis*** of **COVID-19** Vaccination, over time, by age.
  - ***Pasteurian Infectivity*** went up and down wildly in 2021-2022, ending 10-fold higher than at the beginning.
  - ***Gompertzian Lethality*** declined progressively in 2021-2022, ending 10-fold lower than at the beginning.
- The 2021-2022 decline in ***Gompertzian Lethality*** of the vaccinated declined in ***Gompertzian Height***, not ***Slope***.
- The 2021-2022 decline in ***Gompertzian Lethality*** of the unvaccinated declined in ***Gompertzian Height***, not ***Slope***.
- **COVID-19** Infection, like vaccination, pushes down the ***Gompertzian Height*** of ***Gompertzian Lethality***.
- **COVID-19** Infection, and its impact on ***Gompertzian Lethality***, give us a chance to learn how to schedule boosters.

***Gompertzian Analysis*** of the impact of the number of **COVID-19** ***Vaccination Events*** (Primary & Boosters)

I. ISRAEL, Boosters

Each ***Vaccination Event*** decreased **COVID-19** ***Lethality*** (Deaths/Cases) by about 1/3rd.
**COVID-19** vaccine impact: On ***Lethality***, persistent & cumulative; On ***Infectivity***, fleeting.
2 more boosters could push **COVID-19** ***Lethality*** down to the familiar level of **Influenza** ***Lethality***.

II. USA(CDC): March 2022 to July 2022, and October 2021 to Dec 2022, Boosters.

1. Boosters: March 2022 to July 2022: ***Gompertzian Lethality*** (Deaths/Cases)
2. Boosters: March 2022 to July 2022: Persistence of ***Gompertzian Lethality*** (Deaths/Cases)
3. Boosters: March 2022 to July 2022: Lack of Persistence of ***Pasteurian Infectivity*** (Cases/Population)
4. Boosters: ***Gompertzian Lethality***, ***Pasteurian Infectivity***, ***Malthusian Lethality*** compared
5. Boosters, October 2021 to Dec 2022: 3rd Booster (***Bivalent***) decreases **COVID-19** ***Lethality*** further.
6. Boosters: The value of sorting patients by number of ***Vaccination Events***, infections, and age.
7. Boosters: July 2021-June 2022: **COVID-19** ***Lethality*** shows no waning for at least 11 months.

III. HUNGARY, Boosters
IV. Comparison of USA and Israel Booster Studies
V. More recent 2nd Booster Studies

***Gompertzian Analysis*** provides useful values for the benefit, and marginal benefit, of vaccination, by age.
***Gompertzian Analysis*** gives us rational age-based vaccination guidelines.

Who's dying now?

Benefits of higher vaccination compliance and additional ***Vaccination Opportunities***

***Gompertzian Analysis*** of the Impact of Non-Immunological Factors On **COVID-19** Outcome
Vaccines and other things change **COVID-19** ***Lethality*** by pushing up or down ***Gompertz Lines***.
The impact of Sex on All-Cause Mortality
The impact of Sex on **COVID-19** ***Infectivity*** and ***Lethality***.
The impact of Diabetes on **COVID-19** ***Infectivity*** and ***Lethality***.

The impact of VARIANTS on **COVID-19** ***Infectivity*** and ***Lethality***.

***Gompertzian Analysis*** of LONG COVID

***Gompertzian Analysis*** can aid in the formation of **COVID-19** vaccination policy.

#### OVERVIEW

How might **COVID-19** be brought under control? Here, I provide the details noted in the main text of this communication, concerning how data on the numbers of **COVID-19** cases and deaths can yield up hidden suggestions for scheduling, and configuring, vaccines, so as to save many more lives. In the main text of this communication, I have outlined how this new method, ***Gompertzian Analysis***, counting cases and deaths, by age, and displaying them on logarithmic graphs, can identify the magnitude of **COVID-19 *Gompertzian Lethality*** (Deaths/Cases), and **COVID-19 *Pasteurian Infectivity*** (Cases/Population), and the impact which vaccination and other forces can have on these components of **COVID-19** outcome. The lessons from this analysis also give us general insights into infectious disease, and ageing. All of the data examined here were extracted from publicly available datasets, each identified by a reference.

#### THE GOMPERTZIAN FORCE OF MORTALITY AND GOMPERTZ MORTALITY EQUATION

##### Introduction

The key to accomplishing the goal described above lies in examining the numbers of **COVID-19** cases and deaths in light of the ~10,000-fold exponential increase in the risk of **COVID-19 *Lethality*** (Deaths/Cases) that occurs from infancy to old age. This exponential quality for **All-Cause *Mortality*** was discovered by Gompertz in 1825[1] and for **COVID-19 *Lethality*** (Deaths/Cases) by Levin in 2020,[2] and others[3,4,5,6,7,8] (APPENDIX-FIGURE 1). Levin *et al*'s initial report of the log-linear, ***Gompertzian***, appearance of the age association of **COVID-19 *Lethality***[9] (APPENDIX-FIGURE 1a) was subsequently confirmed by a number of additional studies summarizing many countries (APPENDIX-FIGURES 1b, and 1c, values kindly provided by Dr. Brazeau[10,11,12,13]). Levin followed up with a comparative analysis of the age distribution of **COVID-19 *Lethality*** in the developed and undeveloped world[14], revealing a somewhat higher occurrence of death by age, again characterizing the ~10,000-fold exponential difference in **COVID-19 *Lethality*** from infancy to old age (APPENDIX-FIGURE 1).

This exponential increase in lethality with age is known as the ***Gompertzian Force of Mortality***, and is captured by the ***Gompertz Mortality Equation,*** from Gompertz, who reported in 1835[1] that the chance of death for humans, ***D***, increases exponentially with age, ***t***. This ***Gompertzian Force of Mortality*** describes the risk of death generally, and the risk of many diseases, including **COVID-19**[16] (APPENDIX-FIGURE 1):

$$D = G_H \cdot e^{G_S \cdot t} \tag{1}$$

which is equivalent to:

$$\log(D) = (G_S \cdot t) + \log(G_H) \tag{2}$$

Let us call $G_s$, the ***Gompertzian Slope***, and $G_H$, the ***Gompertzian Height***.

##### COVID-19's Fearful *Gompertzian Force of Mortality* Comes into View When Displayed Linearly

As we shall see throughout this report, ***Gompertz Plots***, that is, the log plots of age-sorted data, and their mathematical equivalent, the exponential equation, give us powerful insights into **COVID-19 *Infectivity*** and ***Lethality***, but they do not provide easy visualizations of the things we care about most: how many of us will be affected, what are our individual chances of being affected, and what can be done about it. These more personal and practical understandings come into view by transforming the log ***Gompertz Plots*** into linear graphs (APPENDIX-FIGURE 1, right). The first thing that strikes us by such a linear revisualization is the shocking, progressively more lethal, outcome of **COVID-19** that occurs as we age. For example, on such linear graphs as in APPENDIX-FIGURE 1 (right), we can hardly detect the line of the **COVID-19 *Gompertz Mortality Equation*** (Deaths/Cases) in the first five decades of life, when its ***Gompertzian Force of Mortality*** stealthily accumulates less than 1% chance of death, but from age 60 on, we see the progressively more catastrophic, exponential, increase in **COVID-19 *Lethality*** with age, with the risk of death rising from about 1% at age 60, to about 3% at age 70, to almost 10% at age 80, to about 25% at age 90, and almost 80% at age 100.

##### Early life mortality and **COVID-19 *Lethality*** (Deaths/Cases)

As can be seen in APPENDIX-FIGURE 1b, not only does the risk of **COVID-19** death after infection increase with age, as does the risk of death generally, but there also is a smaller "J-shaped" increase in both types of lethalities early in life, as described by Makeham in 1860 for all-cause mortality,[15], and by the COVID-19 Forecasting Team in 2022 for **COVID-19 *Lethality*** (Deaths/Cases).[16] I won't have anything more to add to this topic in this communication, except to note that this early-life increase in **COVID-19 *Lethality*** (Deaths/Cases), and all-cause infant mortality generally, are active areas of analysis by my colleagues and I, about which we shall have more to say at a later date.

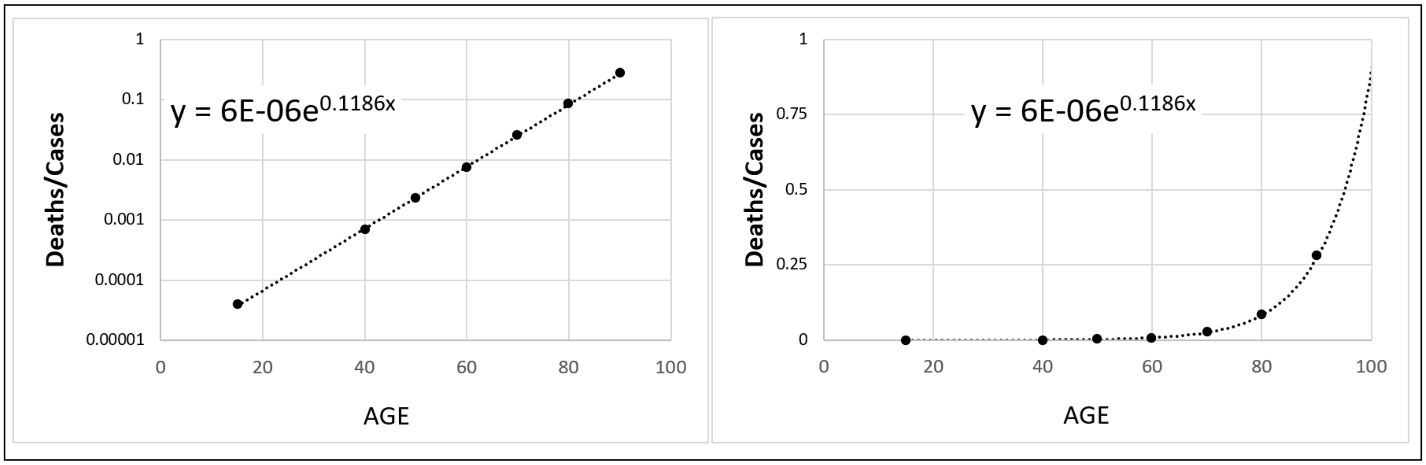

APPENDIX-FIGURE 1a Levin and colleagues [7]

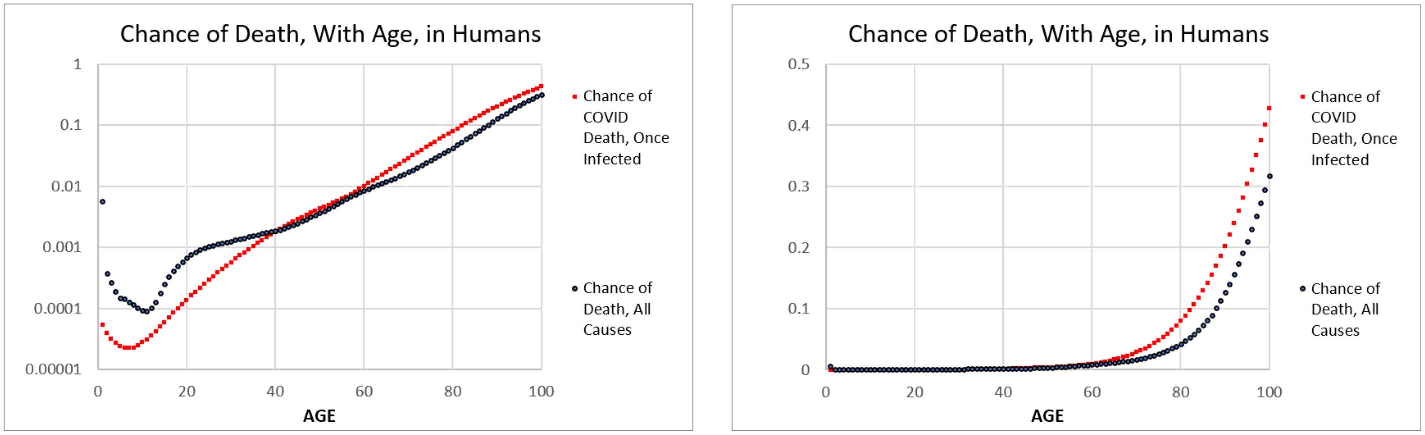

APPENDIX-FIGURE 1b **COVID-19** Forecasting Team [16]

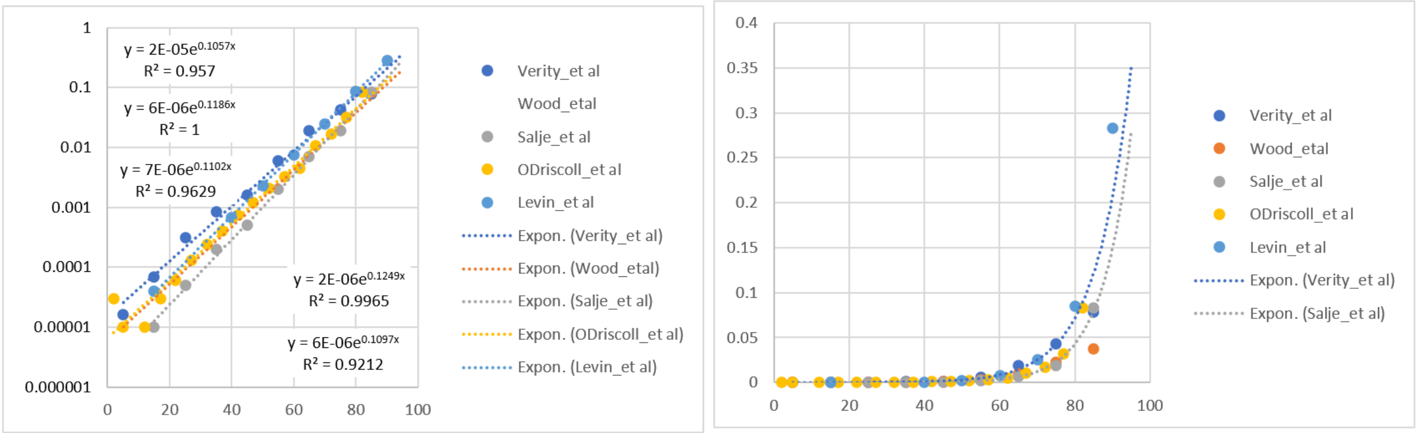

**APPENDIX-FIGURE 1. COVID-19 *Lethality*** (Deaths/Cases) in the Pre-vaccination era, various reports.

Visualization by the ***Gompertz Mortality Equation***:

$$log(D) = (G_S \cdot t) + \log(G_H)$$, which is equivalent to $$D = G_H \cdot e^{G_s \cdot t}$$,

for the relationship between **COVID-19 *Lethality***, ***D***, the risk of death if infected, that is, (Deaths/Cases), vs age, ***t***, captured by the parameters of the ***Gompertzian Slope***, $G_s$, the rate of increase with age, and the ***Gompertzian Height***, $G_H$, the location of the ***Gompertz Line*** on the log graph, up or down.

**UNDERLYING CELLULAR BASIS of COVID-19 *Lethality* and *Infectivity*: CELLULAR PHYLODYNAMICS.**

In this communication, we shall see, again and again, that counting cases and deaths reveals that the risk of **COVID-19 *Gompertzian Lethality*** (Deaths/Cases), and indeed many other causes of death, increases exponentially with age, the ***Gompertzian Force of Mortality***, captured by the ***Gompertz Mortality Equation***. In the companion manuscript to this communication,[17] we have seen how counting our cells as we grow, a method we have called ***Cellular Phylodynamic Analysis***,[18] reveals that this ***Gompertzian Force of Mortality*** is correlated with the decline in the fraction of our cells dividing, the ***Mitotic Fraction***, which occurs as we settle into our final size and years. Indeed, by counting our cells, it has been possible to put into quantitative terms, how, and why, and when, we grow, from single fertilized cells into adults of full size and shape, how the control of cell division harnesses our unruly cells into well behaved multicellular creatures, and how this process ultimately makes us old and ill.[19,20,21] This math reveals that our lifespan is correlated with an age when fewer than one-in-a-thousand cells are dividing, quantifying the long-appreciated mechanism of aging, the failure of cells to be rejuvenated by dilution with new materials made, and DNA repaired, at mitosis.[22,23,24] This ***Cellular Phylodynamic Math*** thus links **COVID-19 *Lethality*** (Deaths/Cases) to our cells. ***Cellular Phylodynamic Math*** also gives us a way to find which cells account for ***Pasteurian Infectivity*** (Cases/Population), and which cells account for ***Gompertzian Lethality*** (Deaths/Cases), as well as to how to make the calculations needed to optimize **COVID-19** vaccination. Details may be found in Reference 17.

#### ***GOMPERTZIAN ANALYSIS*: PRINCIPLES**

***Gompertzian Analysis***: Introduction

The exponential increase in **COVID-19 *Lethality*** (Deaths/Cases) from infancy to old age can be captured mathematically by the ***Gompertz Mortality Equation*** (Equations #1 and #2), and visualized graphically by sorting patients by age and examining deaths and cases on log graphs, called here "***Gompertz Plots***", yielding straight rows of datapoints, called here "***Gompertz Lines***" (APPENDIX-FIGURE 1). I have called this method "***Gompertzian Analysis***". Values for the two parameters of the ***Gompertz Mortality Equation***, the "***Gompertzian Slope***", $G_s$, and the "***Gompertzian Height***", $G_H$, emerge by fitting the data to the exponential equation.

**Terminology: *Vaccination Opportunity*** and ***Vaccination Event***.

The term ***Vaccination Opportunity*** will be used to describe an approved opportunity for patients to receive a primary vaccination or booster. The term ***Vaccination Event*** will be used to describe the actual participation in such a ***Vaccination Opportunity***.

***Gompertzian Analysis*** treats resistance to **COVID-19 *Infectivity*** and ***Lethality*** as two separate processes.

Key to gaining useful insight into vaccine impact is to approach resistance to **COVID-19 *Infectivity*** (Cases/Population) and ***Lethality*** (Deaths/Cases) as separate processes. As we shall see, many findings of ***Gompertzian Analysis*** provide an empirical support for such an approach, as well as the practical utility of such double approach. Indeed, as we shall see, by examining **COVID-19 *Lethality***, and **COVID-19 *Infectivity***, separately, a straightforward strategy for reducing the lethal burden of **COVID-19** emerges. For a closer look at this dual approach, see: "***Gompertzian Analysis*** finds that resistance to **COVID-19 *Infectivity*** and ***Lethality*** are two separate processes" in the text.

Vaccines and other things push up and down ***Gompertz Lines***, defining **COVID-19 *Lethality*** by ***Gompertzian Height***, $G_H$,

For reasons that are biologically and mathematically subtle, but needn't concern us here, as we shall see below, things that lead to a greater risk of **COVID-19** death (diabetes, male sex) appear on a log plot as a rise in the ***Gompertz Line***, reflecting an increase in the ***Gompertzian Height***, $G_H$, while things that lead to a lower risk of **COVID-19** death (vaccination, survived **COVID-19** infection) appear as a decline in the ***Gompertz Line***, reflecting a decrease in the ***Gompertzian Height***, $G_H$, without a marked change in the ***Gompertzian Slope***, $G_s$. This gives us a simple single number, $G_H$, the ***Gompertzian Height***, which can allow us to examine, predict, and test, the combined impacts of morbidity factors, ***Vaccination Events***, infections, and age, into a single measure of **COVID-19 *Lethality*** (Deaths/Cases).

*__Gompertzian Analysis__* <u>of Other Infectious Diseases</u>

A number of other infectious disease, but by no means all, display the log-linear ***Gompertzian*** case fatality relationship noted above for **COVID-19**.[25] ***Gompertz Plots*** of **COVID-19**, Hepatitis, and Influenza can be seen in APPENDIX-FIGURE 2.

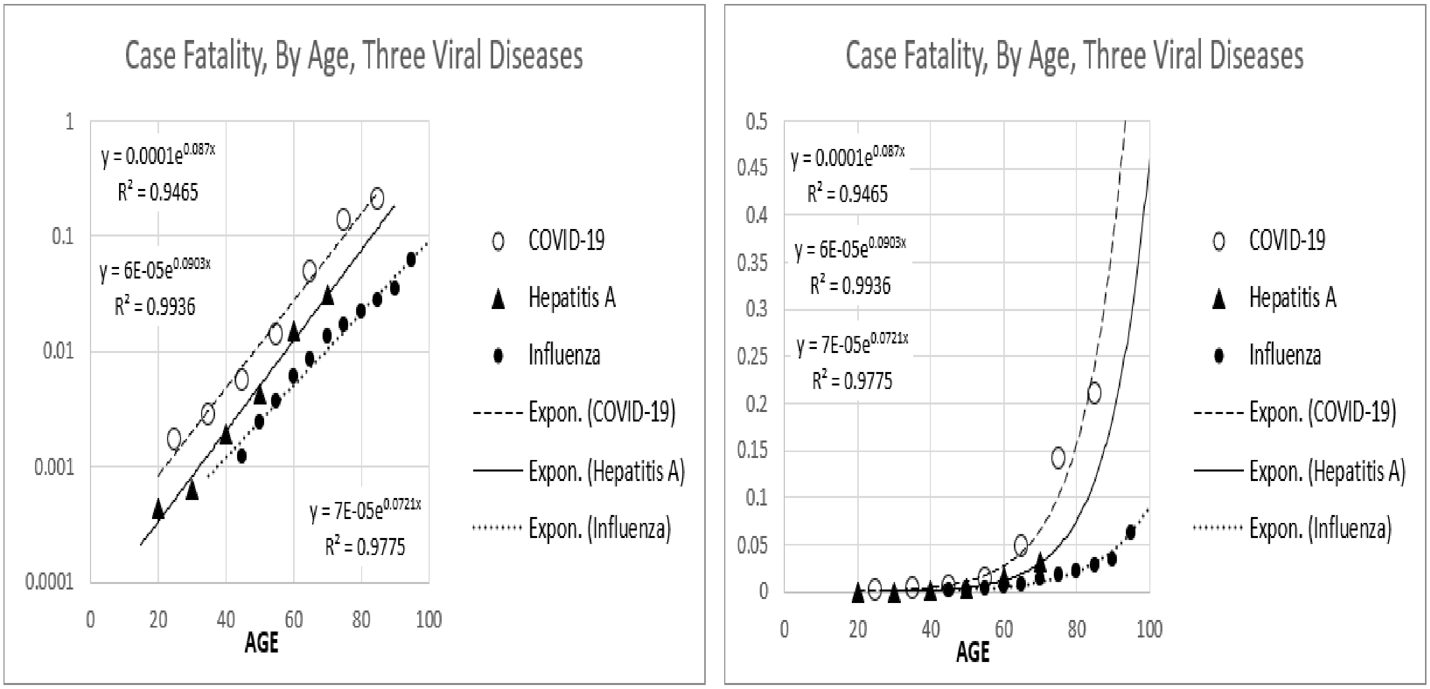

<u>**APPENDIX-FIGURE 2**</u>[25]

***Gompertzian Analysis*** of Non-Infectious Diseases

Many individual causes of death, not just infectious disease, have also been found to form ***Gompertz Lines***, as can be seen in APPENDIX-FIGURE 3 of data assembled by Jones[26]. Thus, the log-linear ***Gompertzian*** appearance of **COVID-19** death is not a specific phenomenon of that disease, but a general phenomenon of aging, and most of the maladies of longevity. This wide spectrum of illnesses, all displaying the ***Gompertzian Force of Mortality***, may well reflect the body-wide decline in functional cells caused by the slowing of growth, and thus the decline in the fraction of cells dividing that marks adult life, as noted above, and shown in the companion manuscript to this communication.[17]

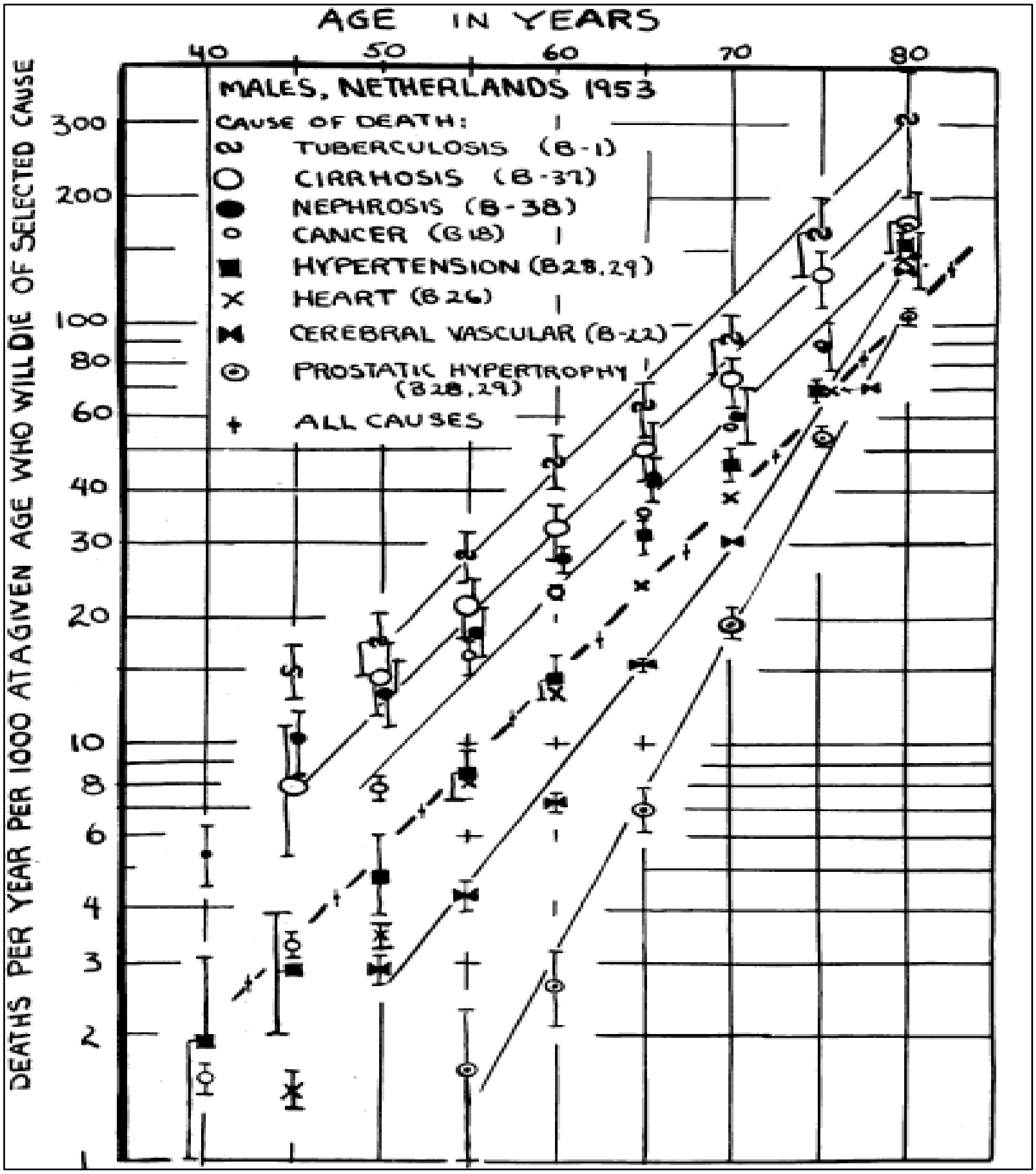

**APPENDIX-FIGURE 3**[26]

***Gompertzian Analysis*** gives population-wide values from partial age samples.

Seldom do we have data from people of all ages. However, the exponential quality of the ***Gompertz Line*** allows us to make estimates of lethality over all ages from data on patients of just a few ages, whether young or old. An example of this can be seen in data from the USA[27] shown in APPENDIX-FIGURE 4, where the exponential fits made from patients of age 21-60, and 61-89, and all ages from 21-89, yields similar ***Gompertz Lines***, with similar ***Gompertzian Slopes***, $G_S$, and ***Gompertzian Heights***, $G_H$ (APPENDIX-FIGURE 4). Note in APPENDIX-FIGURE 4b, how using data points provided by the **COVID-19** Forecasting Team[16] (APPENDIX-FIGURE 1), values from age 40 to 70, or from 80 to 100, yields quite similar ***Gompertz Lines***, over the full life range of life from age 40 to 100. This means that we needn't have a full age sample of a population to make a reasonable picture of **COVID-19** ***Lethality*** over the full adult age range of a population.

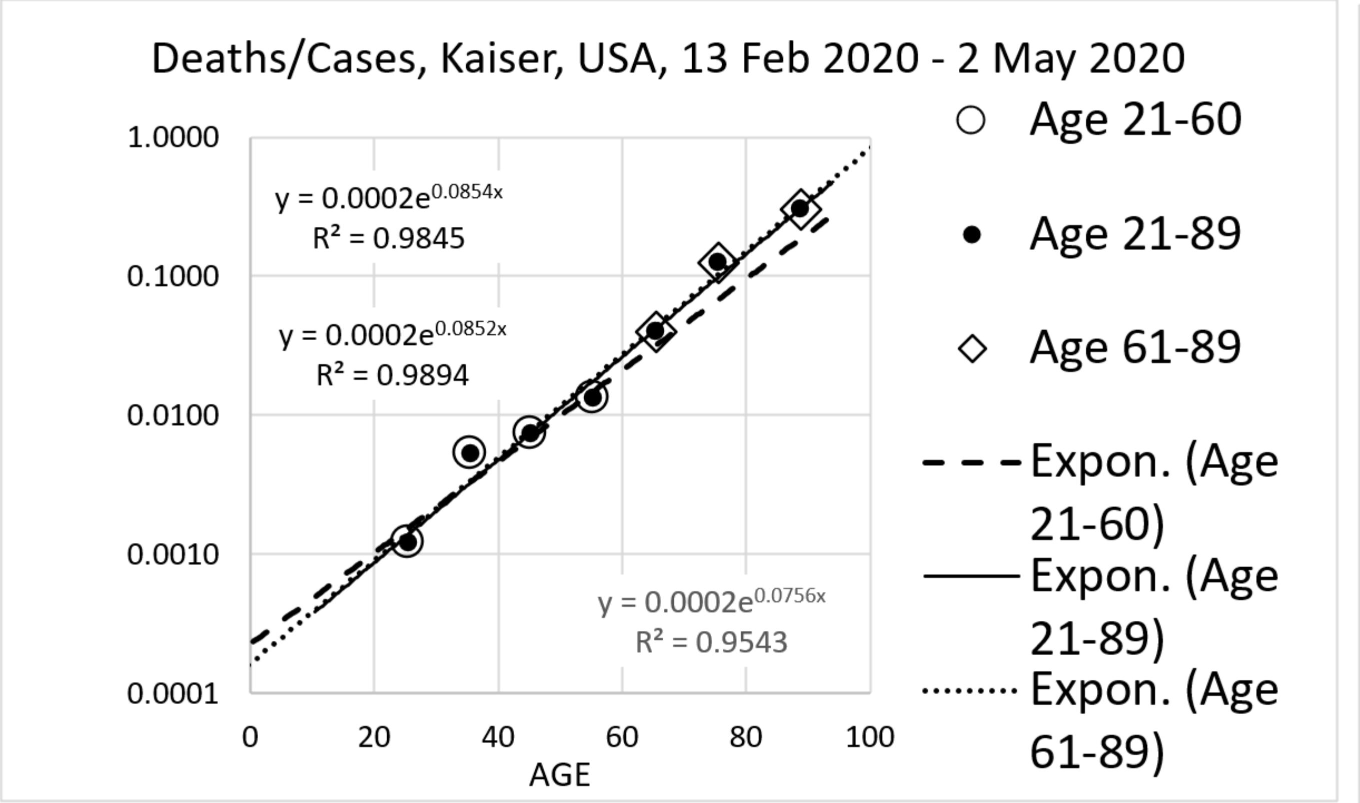

APPENDIX-FIGURE 4a

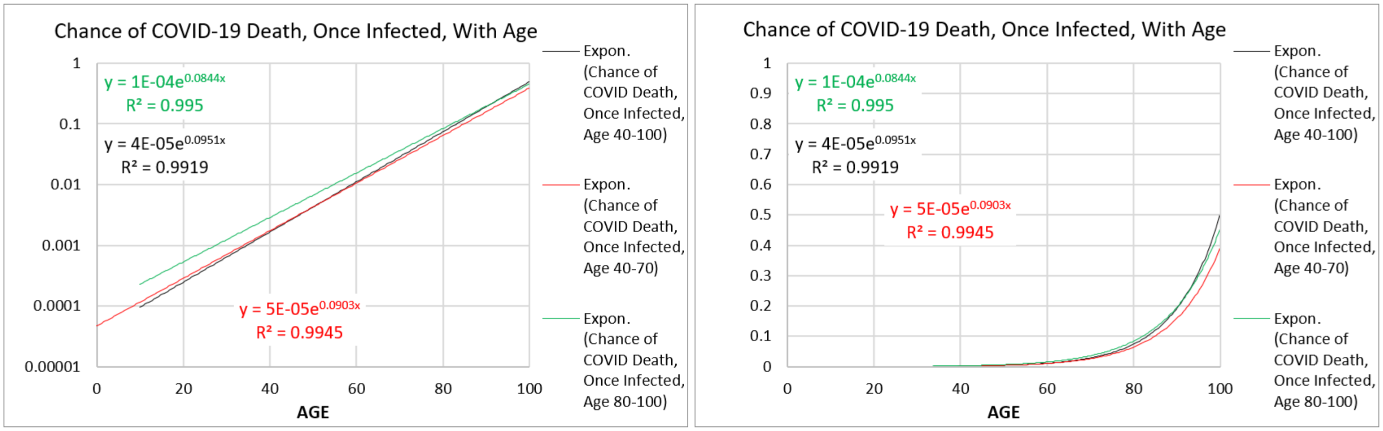

APPENDIX-FIGURE 4b.[16]

***Gompertzian Analysis*** of Hospitalization.

Hospitalization is often used for a metric of **COVID-19** vaccination outcome. However, when examined in terms of its relationship to age, an obvious correspondence to death was not seen, and thus I didn't use it any further (APPENDIX-FIGURE 5).[28]

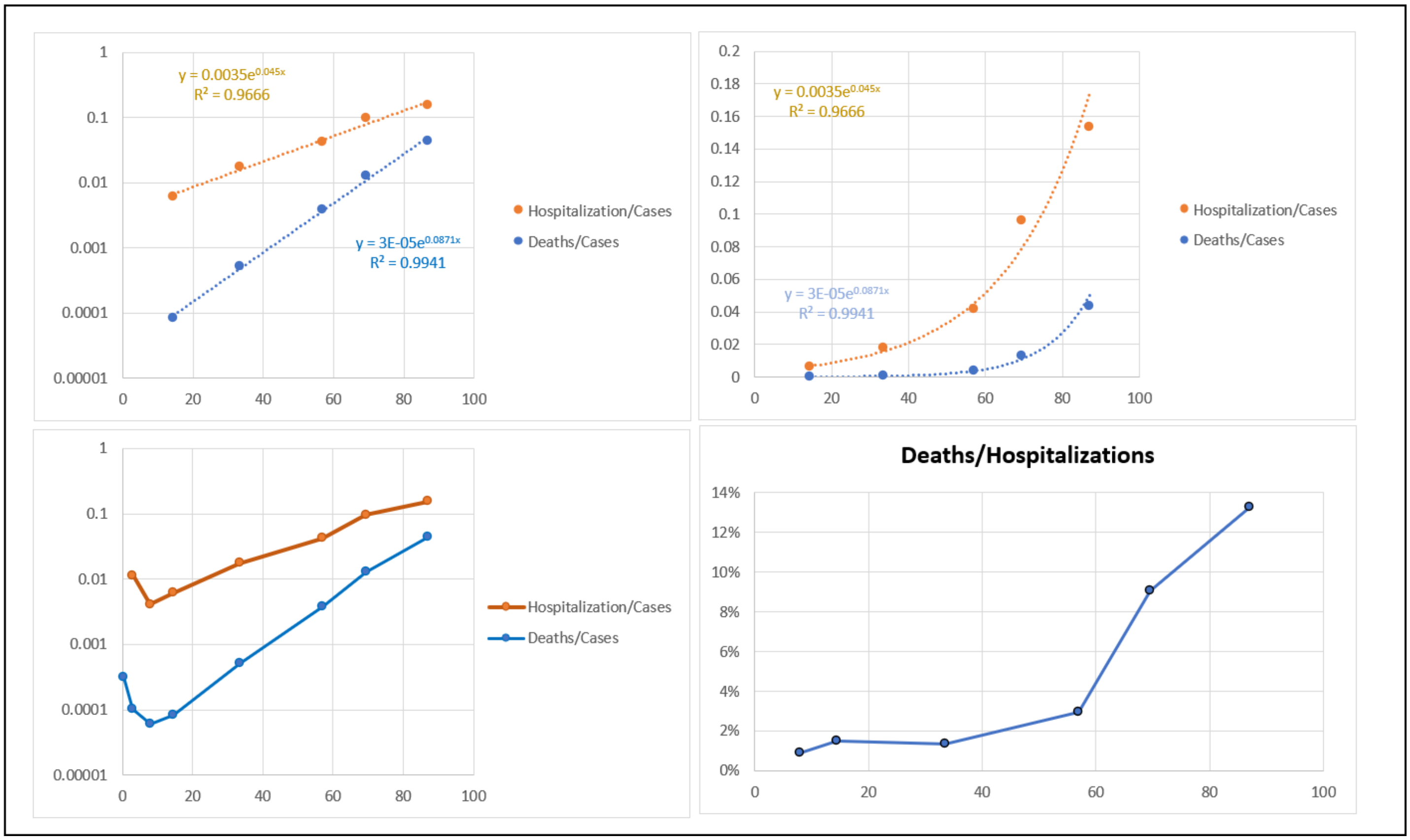

APPENDIX-FIGURE 5[28]

***Gompertzian Analysis*** with Full Age Information

The public data used here are provided in age-group categories, usually open-ended for the youngest and oldest groups, which reduces the potential precision of ***Gompertzian Analysis*** to characterize the parameters of ***Gompertzian Lethality***, ***Pasteurian Infectivity***, ***Malthusian Lethality***, and their ***Reductions*** by vaccination and other forces. Fortunately, more accurate measures of the parameters of the ***Gompertz Mortality Equation*** can be made when the age of each patient is known, rather than placed in an age-group, as has been described by Meuller et al[29] and others. Those with direct access to larger datasets, and thus with knowledge of the age of each case and death, should benefit from this opportunity. Analysis with the knowledge of every patient's age will become especially useful when sequential vaccinations have pushed the **COVID-19** ***Lethality*** down to very low levels.

#### THE PROBLEM WE FACE.

In APPENDIX-FIGURE 6, are shown data from a CDC dataset of approximately 70% of all **COVID-19** cases in the USA from April 2021 to June 2022, containing data on the disease among ~180,000,000 Americans.[30]. Using these data, we have shown **COVID-19 *Infectivity*** (Cases/Population) and **COVID-19 *Lethality*** (Deaths/Cases) for the vaccinated (APPENDIX-FIGURE 6 left and center) and unvaccinated (APPENDIX-FIGURE 6 right), in the 65-79 age group. Data from other age groups show essentially the same outcomes.

In APPENDIX-FIGURE 6, we can see that vaccination reduces **COVID-19 *Lethality*** (Deaths/Cases), which continues to decline for the vaccinated, no doubt from boosters and breakthrough infections. We can also see that the unvaccinated also show a decline in **COVID-19 *Lethality*** (Deaths/Cases), no doubt from infections, at the cost of lives lost (APPENDIX-FIGURE 6 bottom).

In APPENDIX-FIGURE 6, we can also see that vaccination also reduces **COVID-19 *Infectivity*** (Cases/Population). However, for both vaccinated and unvaccinated, numbers of infections increase 100-fold, ending the 14-month period 10-fold higher than when it began.

These data show how much progress has occurred in reducing **COVID-19 *Lethality***, and how challenging it has been to find a way to persistently reduce **COVID-19 *Infectivity***. In the ***Gompertzian Analysis*** to follow, we shall see how the information we extract from this, and other, datasets can tell us how we make the most out of the tools we have and save many more lives with vaccination.

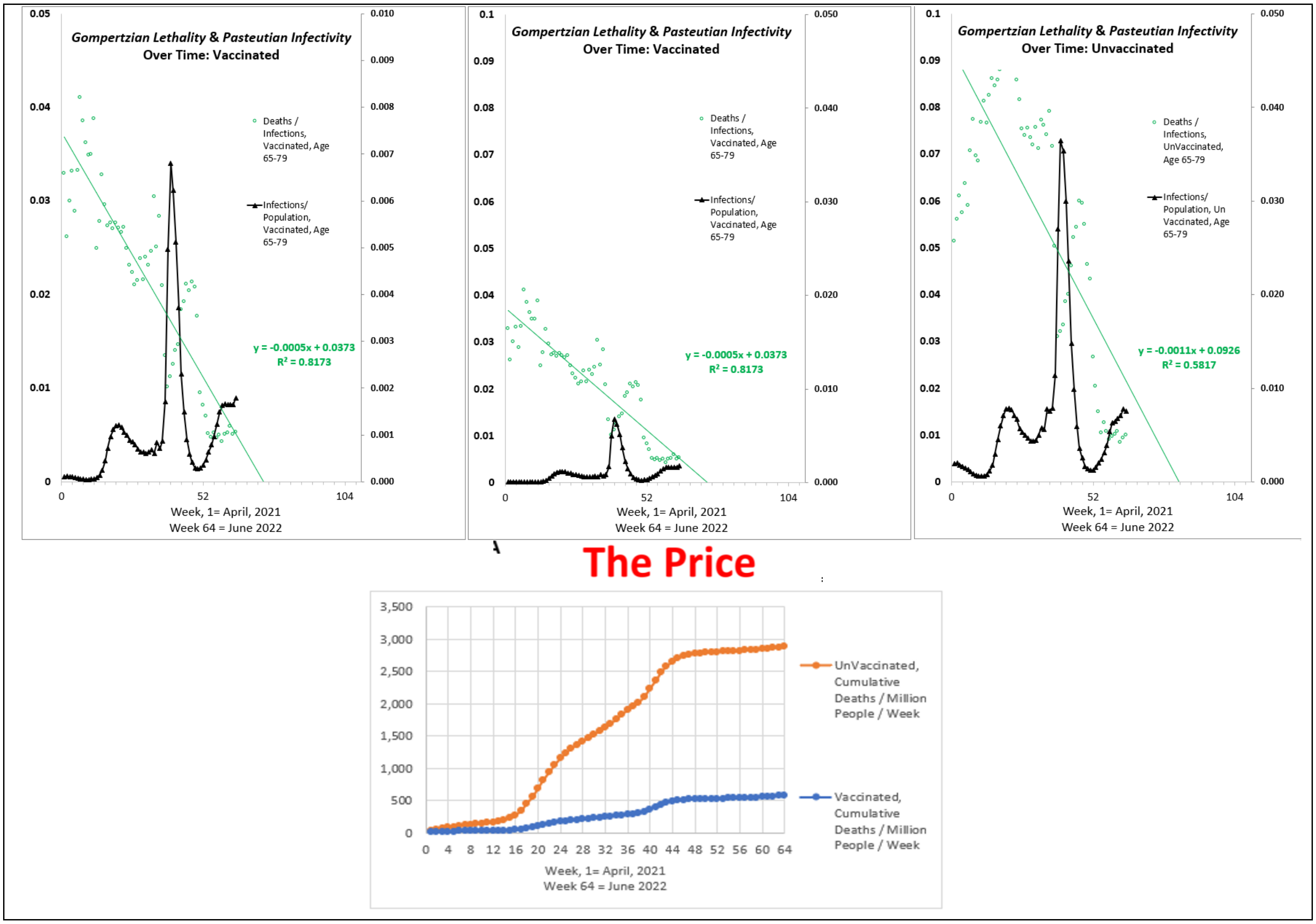

APPENDIX-FIGURE 6: See next page for larger version

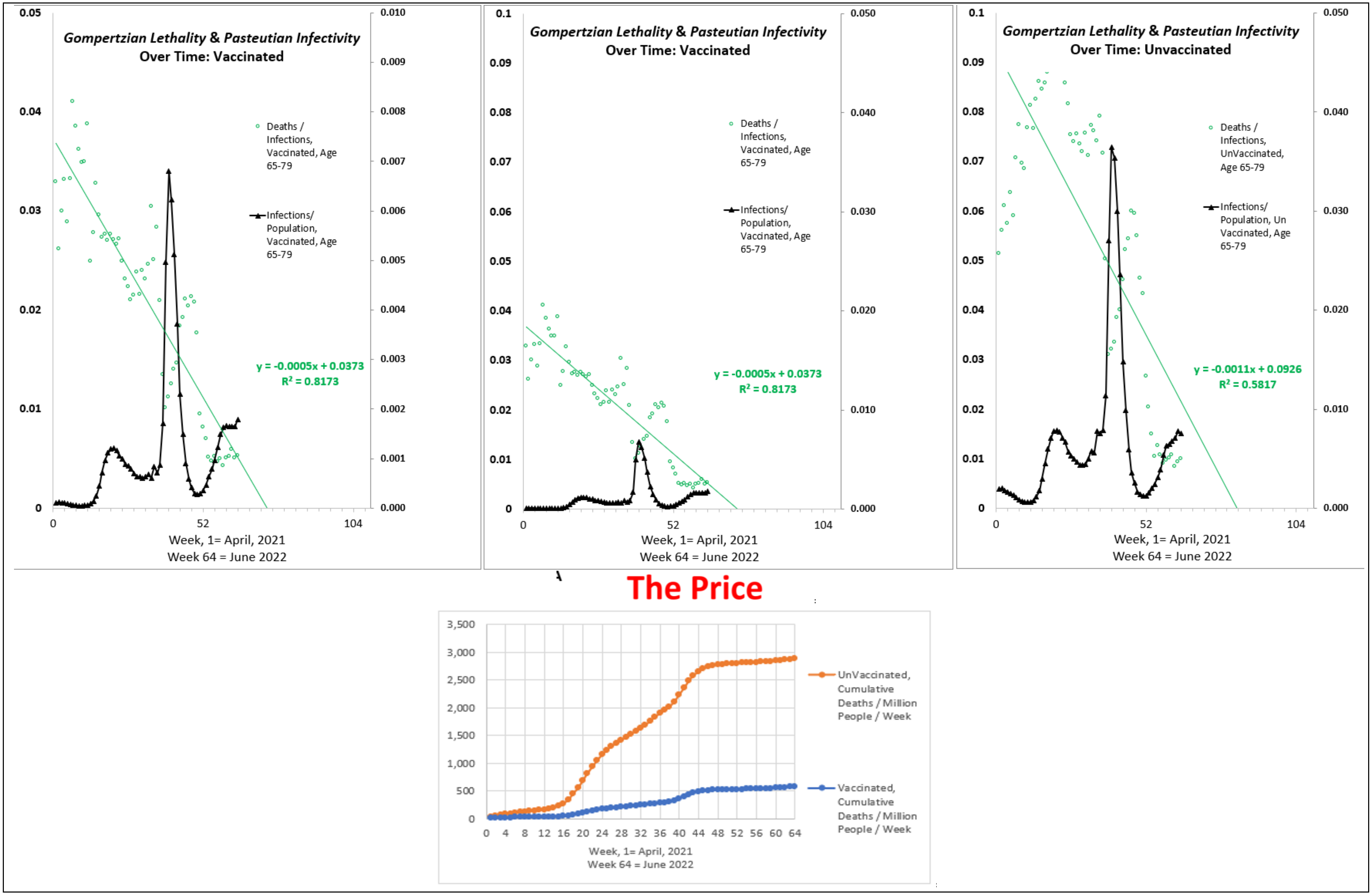

APPENDIX-FIGURE 6 (Cont.)

**GOMPERTZIAN ANALYSIS: COVID-19**

***Gompertzian Analysis* gives 3 new measures: *Pasteurian Infectivity*, *Gompertzian Lethality*, *Malthusian Lethality*.**

***Gompertzian Analysis***, that is, sorting patients by age, counting cases and deaths, and displaying these numbers on logarithmic graphs, has made it possible to generate three new practical measures of **COVID-19** impact:

- ***Pasteurian Infectivity***: Cases/Population.
- ***Gompertzian Lethality***: Deaths/Cases.
- ***Malthusian Lethality***: Deaths/Population.

All three measures, when compared against age, appear as ***Gompertz Lines*** on logarithmic ***Gompertz Plots***, shown in APPENDIX-FIGURE 7, with data from a rather extraordinary CDC dataset of approximately 70% of all **COVID-19** cases in the USA from April 2021 to June 2022.[31] Similar examples of ***Gompertzian Analysis*** can be seen in many other figures throughout the main text of this communication and in this APPENDIX. Note in APPENDIX-FIGURE 7, how ***Gompertzian Analysis*** distills down each of these three measures -***Pasteurian Infectivity***, ***Gompertzian Lethality***, ***Malthusian Lethality***- to two numbers -***Gompertzian Height***, $G_H$, and ***Gompertzian Slope***, $G_S$- of the ***Gompertz Mortality Equation*** (#1 and #2, see above). For ***Gompertzian Lethality*** and ***Malthusian Lethality***, this exercise captures the ~10,000-fold rise in lethality with age.

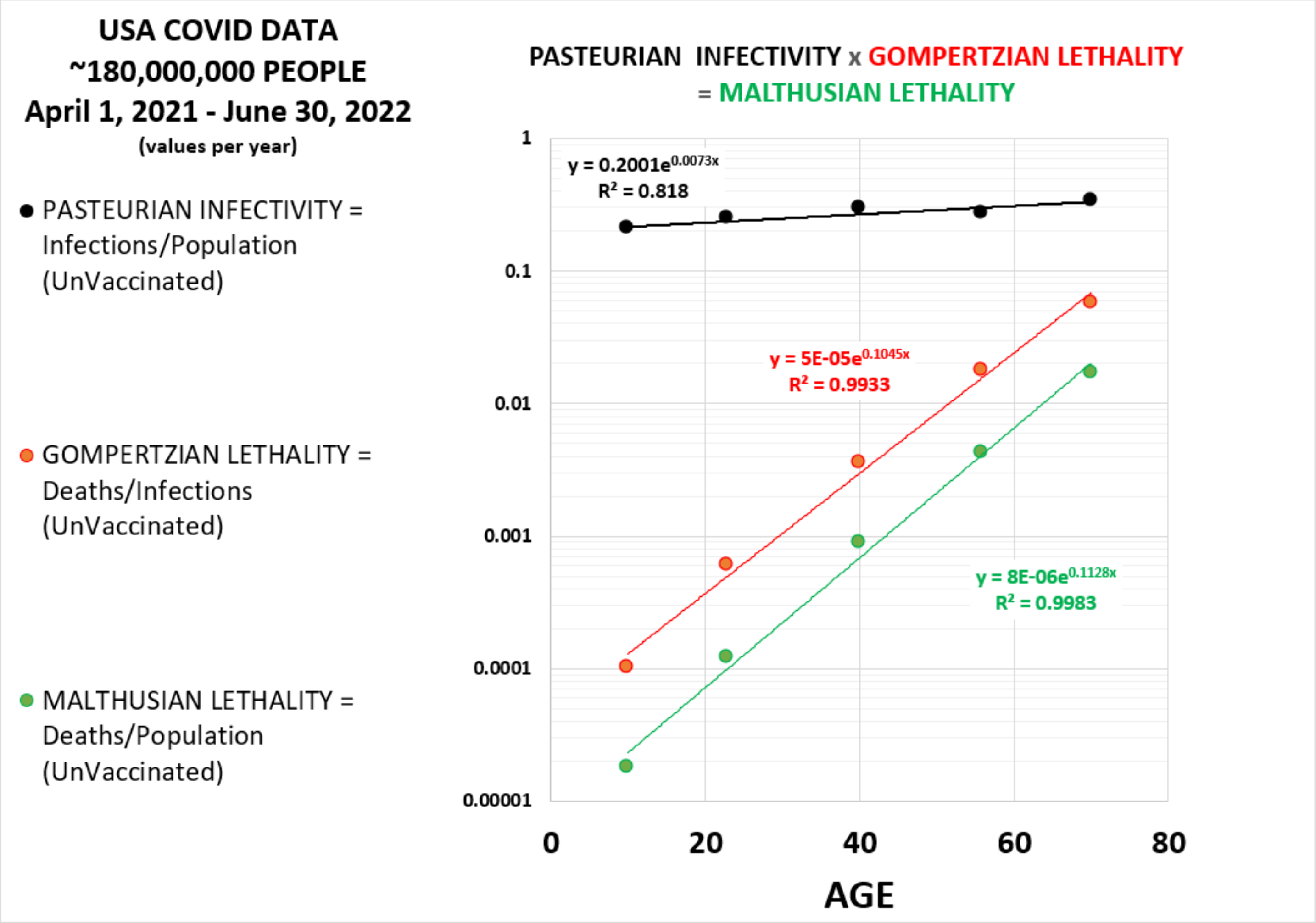

APPENDIX-FIGURE 7

**Terminology: *Gompertzian Lethality*, COVID-19 *Lethality*, *Pasteurian Infectivity*, COVID-19 *Infectivity*.**

To ease communication of the basic findings of ***Gompertzian Analysis*** in the main body of this communication, to which this is the APPENDIX, we shall refer to **COVID-19 *Gompertzian Lethality*** (Deaths/Cases) simply as **COVID-19 *Lethality*** (Deaths/Cases), and **COVID-19 *Pasteurian Infectivity*** (Cases/Population) simply as **COVID-19 *Infectivity*** (Cases/Population). The longer terms will be most relevant for examining infectious disease generally.

***Gompertzian Analysis*** of **COVID-19 *Infectivity*** and ***Lethality*** in USA 2021-2022, analyzed in aggregate.

Basic Features, on Graphs.

We can see these essential qualities of **COVID-19 *Infectivity*** and ***Lethality***, by sorting data by age, and examining the information on logarithmic ***Gompertz Plots***, as we have done in APPENDIX-FIGURE 7 for the CDC dataset. Note, that both ***Gompertzian Lethality*** (Deaths/Cases), and ***Malthusian Lethality*** (Deaths/Population), display the alarming log-linear distribution in chance of death, in agreement with Levin's data, which revealed the ~10,000-fold, log-linear difference in **COVID-19 *Lethality*** from infancy to old age (APPENDIX-FIGURE 7). On the other hand, the CDC data displays only a small, probably irrelevant, 2-fold, difference between the young and the old in ***Pasteurian Infectivity***, the fraction of people in the population who have been infected (Cases/Population).

Thus, these data reveal that age has little, if any, practical impact in the chance of infection, but an enormous impact on the chance of death if infected. Old people aren't noticeably more likely to get infected by **SARS-CoV-2**, but they are catastrophically more likely to die of **COVID-19** if infected.

Basic Features, in Numbers: 3 new measures are each captured by a single number: the ***Gompertzian Height, $G_H$***.

The associations of ***Pasteurian Infectivity***, ***Gompertzian Lethality***, and ***Malthusian Lethality*** with age could be quantified by fitting each set of points to the exponential equation.

For ***Gompertzian Lethality*** (Deaths/Cases) and ***Malthusian Lethality*** (Deaths/Population), these exponential regressions distilled each of these qualities down to just two numbers: the ***Gompertzian Height***, $G_H$, and ***Gompertzian Slope***, $G_S$, and the ***Malthusian Height***, $M_H$, and ***Malthusian Slope***, $M_S$.

For ***Pasteurian Infectivity***, these exponential regressions capture the small, probably irrelevant, 2-fold, difference between the youngest and oldest reflected in the ***Pasteurian Slope***, $G_S$, being close to zero. Since any number to the zero power equals 1, as a useful approximation, the ***Pasteurian Slope***, $G_S$, can be ignored, with the measure of ***Pasteurian Infectivity*** reflected simply in the ***Pasteurian Height***, $P_H$. The small difference in ***Pasteurian Infectivity*** from age to age can be comprehended by this math, a topic that would be most worthy of analysis, but as a first approximation, it won't be taken into account here.

For ***Gompertzian*** and ***Malthusian Lethality***, the ***Slopes***, $M_S$ and $G_S$, are quite similar, and we shall see that this remains the case as these features are modified by many forces, including vaccination and virus genotype. This mathematically subtle point is a topic of continuing analysis by my colleagues and I, but, for now, we need only appreciate that this simplifies our analysis of **COVID-19 *Lethality***.

Thus, ***Pasteurian Infectivity*** and ***Gompertzian Lethality***, and ***Malthusian Lethality*** can each be captured by the single parameter of ***Height***: $G_H$, $P_H$, and $M_H$. For all three qualities, higher is bad, lower is good.

***Gompertzian Lethality*** and ***Case Fatality***.

Both ***Gompertzian Lethality*** and ***Case Fatality*** measure (Deaths/Cases). However, because (Deaths/Cases) displays an ~10,000-fold exponential rise in lethality with age, even slight differences in ages of the individuals in a group will skew the measure of ***Case Fatality***. In contrast, this does not affect ***Gompertzian Lethality***, which provides a number, $G_H$, the ***Gompertzian Height*** of ***Gompertzian Lethality*** of **COVID-19 *Lethality,*** which is independent of patient age. Such a calculation was not available before.

***Reductions***: 3 additional new measures of ***Gompertzian Analysis*** for measuring impact of vaccination and other forces.

***Gompertzian Analysis*** gives 3 additional new measures for measuring impact of vaccination and other forces on **COVID-19** called ***Reductions***. ***Pasteurian Infectivity Reduction***, ***Gompertzian Lethality Reduction***, and ***Malthusian Lethality Reduction***) calculated for each age group as "1-(vaccinated/unvaccinated)", and all of the age groups are then averaged (APPENDIX-FIGURE 8). These three new measures add additional information not available before from other measures, such as ***Vaccine Effectiveness***.

***Pasteurian Infectivity Reduction***: The Effect of Vaccines on the Risk of Infection.

***Pasteurian Infectivity Reduction*** and ***Vaccine Effectiveness Against Infection*** are both calculated in terms of 1-vaccinated/unvaccinated (Cases/Population). Because ***Pasteurian Infectivity*** occurs at similar rates across ages, the two measures are roughly equivalent.

***Malthusian Lethality Reduction***: The Effect of Vaccines on the Aggregate Risk of Infection and Death Combined.

***Malthusian Lethality Reduction*** and ***Vaccine Effectiveness Against Death*** are both calculated in terms of 1-vaccinated/unvaccinated (Deaths/Population). However, because (Deaths/Population) displays an ~10,000-fold exponential rise in lethality with age, even slight differences in ages of the patients in the vaccinated and unvaccinated groups may lead to imprecision in the measure of ***Vaccine Effectiveness Against Death***. In contrast, this does not affect ***Malthusian Lethality Reduction***, which provides a measure of vaccine impact that is independent of patient age.

***Gompertzian Lethality Reduction***: The Effect of Vaccines on Risk of Death Once Infected

***Gompertzian Lethality Reduction*** provides the measure for how vaccines, and other forces, affect just **COVID-19 *Lethality***. Such a calculation was not available before. As we shall see below, this new measure of vaccine impact is key for identifying how to utilize vaccination so as to achieve the greatest possible reduction in **COVID-19** death.

***Gompertzian Analysis*** of the impact of **COVID-19** vaccination in USA 2021-2022, analyzed in aggregate.

***Reductions***: Basic Features, on Graphs.

In APPENDIX-FIGURE 8, data have been added to that seen on APPENDIX-FIGURE 7 on the people that had been vaccinated. Note how vaccination has led to a ***Reduction*** in ***Pasteurian Infectivity***, the fraction of people in a population who have been infected (Cases/Population), and to a ***Reduction*** in ***Gompertzian Lethality***, the fraction of infected people who have died (Deaths/Cases). The aggregate effect is seen in a ***Reduction*** of ***Malthusian Lethality***, the fraction of people in a population who have died (Deaths/Population).

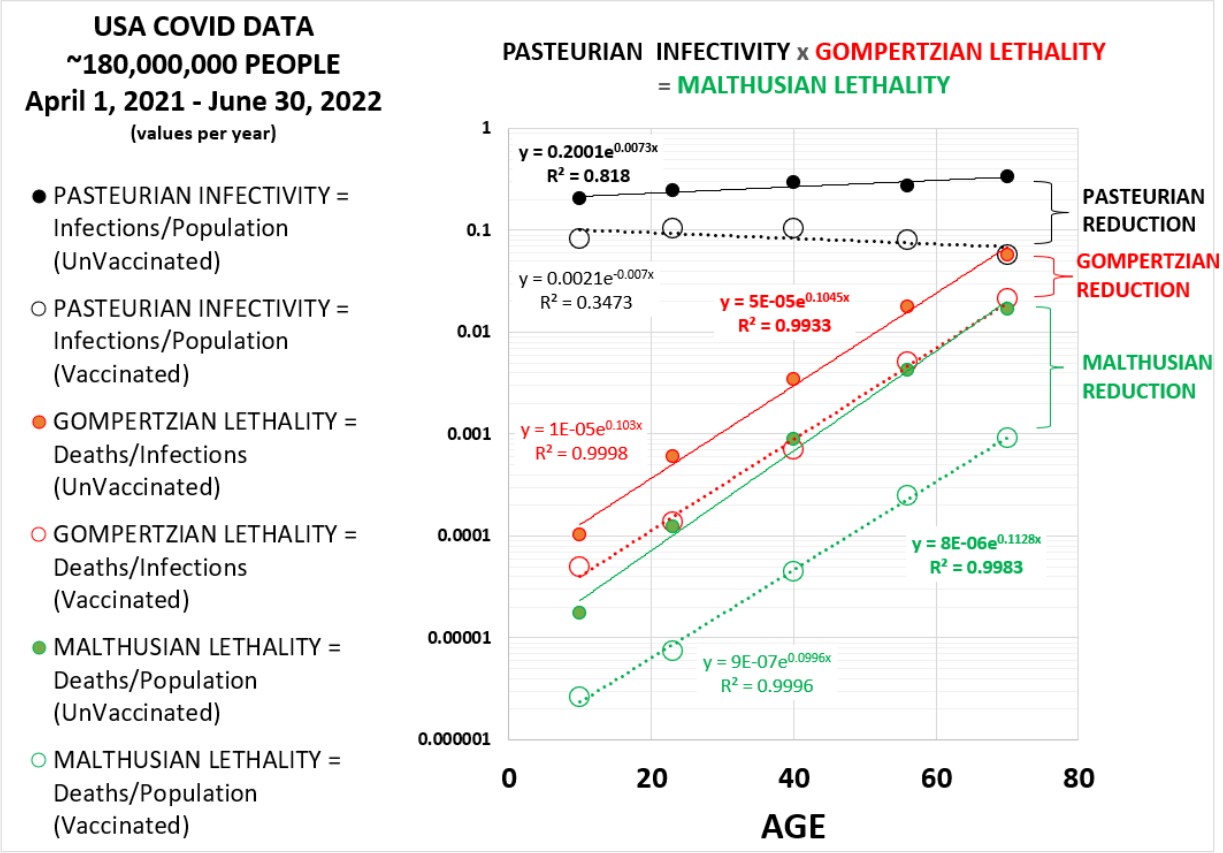

APPENDIX-FIGURE 8

***Reductions***: Basic Features, in Numbers

***Reductions*** may be expressed as "***percentages***" or "***folds***", based on the measure of vaccine action that is most useful for communication. Let us now examine these numbers for the patients in the CDC dataset.

***Reductions as Percentages***

Recall that vaccine activity can be measured by its "***Reduction***," "1-(vaccinated/unvaccinated)", expressed as a percentage.

Calculations with the data shown in APPENDIX-FIGURE 9 show that the ***Gompertzian Reduction***, that is the reduction in ***Gompertzian Lethality*** (Deaths/Cases) of vaccinated compared to unvaccinated, averaged from all of the age groups, was 73%.

The ***Pasteurian Reduction***, that is the reduction in ***Pasteurian Infectivity*** (Cases/Population) of vaccinated compared to unvaccinated, averaged from all of the age groups, was 83%.

The ***Malthusian Reduction***, that is the reduction in ***Malthusian Lethality*** (Deaths/Population) of vaccinated compared to unvaccinated, averaged from all of the age groups, was 94%.

***Reductions as Fold Changes***

Sometimes it can be useful to frame a vaccine's impact in terms of "***how many-fold***", as calculated simply by: (unvaccinated/vaccinated).

As shown in APPENDIX-FIGURE 9 the ***Gompertzian Reduction*** of 73% caused by vaccination in the USA population reflects a **4-*fold*** reduction in in ***Gompertzian Lethality*** (Deaths/Cases).

Similarly, the ***Pasteurian Reduction*** was 83% reflects a **6-*fold*** reduction in ***Pasteurian Infectivity*** (Cases/Population).

The ***Malthusian Reduction*** 94% in this dataset reflects a **17-*fold*** reduction in ***Malthusian Lethality*** (Deaths/Population).

***Reduction*** in Contrast to ***Vaccine Effectiveness***

As noted above, ***Pasteurian Infectivity Reduction*** and ***Vaccine Effectiveness Against Infection*** are roughly equivalent, as ***Pasteurian Infectivity*** occurs at similar rates across ages.

***Malthusian Lethality Reduction*** and ***Vaccine Effectiveness Against Death*** both measure the combined impact of infection and lethality. However, ***Vaccine Effectiveness Against Death*** is highly susceptible to distortion by age, which ***Malthusian Lethality Reduction*** is not.

The ***Gompertzian Lethality Reduction*** provides a measure of the impact of vaccination on ***just Gompertzian Lethality*** (Deaths/Cases), a measure that was not available before. Again, as we shall see below, this affords new, key, insights, into how we can schedule primary vaccination and boosters, so as to bring **COVID-19** death under control.

***Gompertzian Analysis*** of **COVID-19** Primary Vaccination: VA(USA), Vaccine Impact

Two studies carried out at the US Veterans Affairs healthcare system, examined the impact of **COVID-19** primary vaccination. In the first study, outcome for vaccinated individuals were compared with unvaccinated controls, December 11, 2020, to March 25, 2021, that is, about 3½ months, ending a few days before the CDC dataset reviewed above begins, which itself continued for the next 15 months, up until June 30, 2022.[32] The VA group then examined a follow-up to June 30, 2021.[33] Note in APPENDIX-FIGURE 9 how for this VA population, as we saw earlier for the CDC dataset, that vaccination led to a reduction in ***Pasteurian Infectivity***, the fraction of people in a population who have been infected (Cases/Population), and in ***Gompertzian Lethality***, the fraction of infected people who have died (Deaths/Cases). The multiple, the aggregate reduction in the ***Malthusian Lethality***, the fraction of people in a population who have died (Deaths/Population), also displayed its reduction, as in the CDC population dataset.

The ***Gompertzian Reduction*** (Deaths/Cases) of vaccinated compared to unvaccinated was 53%.

The ***Pasteurian Reduction*** (Cases/Population) of vaccinated compared to unvaccinated was 69%.

The ***Malthusian Reduction*** (Deaths/Population) of vaccinated compared to unvaccinated was 87%.

Note again how the ***Gompertz Lines*** of the vaccinated and unvaccinated are roughly parallel, indicating roughly equivalent reductions in all age groups.

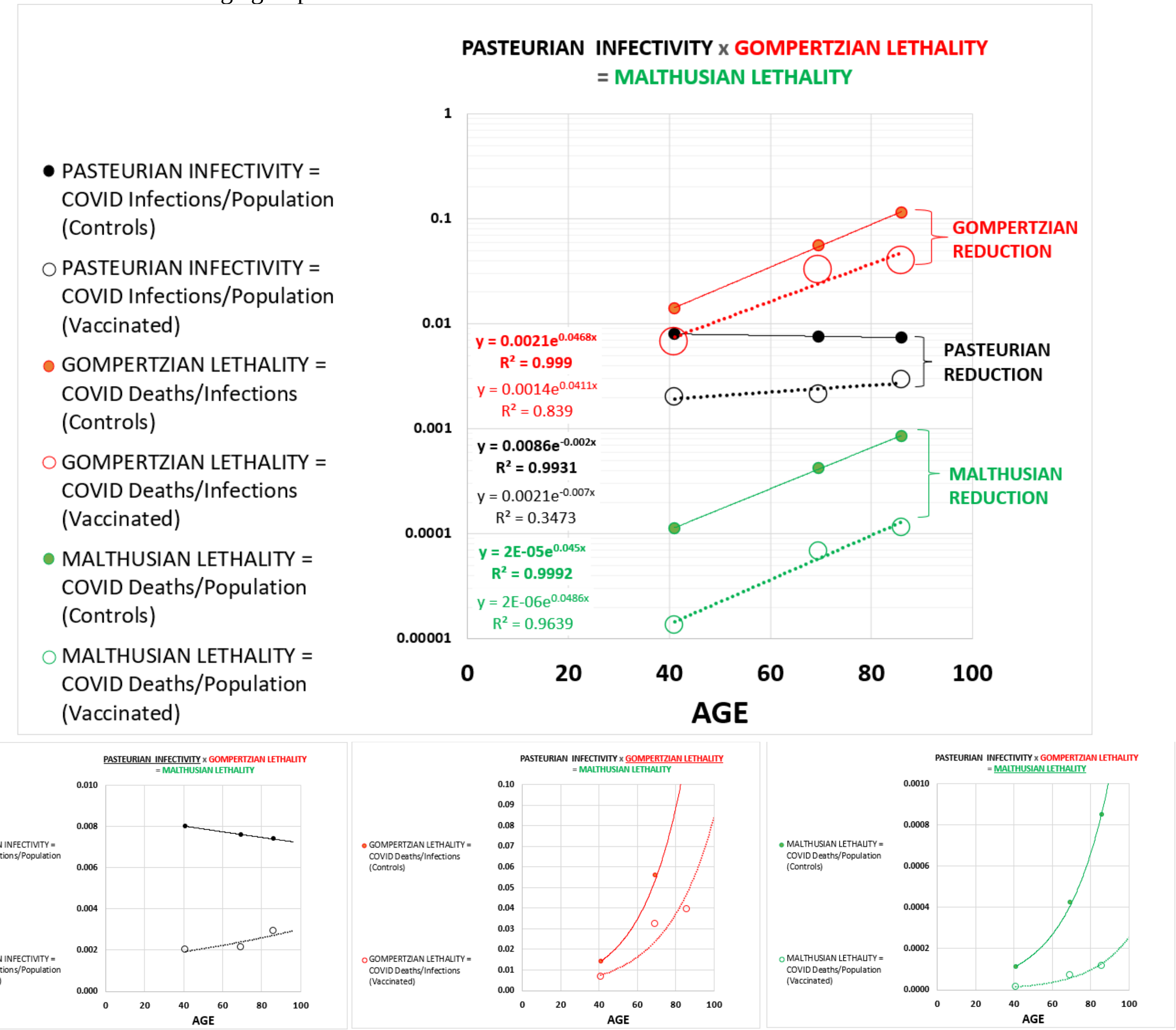

APPENDIX-FIGURE 9

<u>***Gompertzian Analysis*** of **COVID-19** Vaccination: VA(USA), Primary Vaccine Comparisons</u>

In the second study of this population by this research group, during the same period, one primary vaccine was compared against another (Moderna Vs Pfizer) (APPENDIX-FIGURE 10). Both primary vaccines yielded quite similar ***Gompertz Lines*** of ***Gompertzian Lethality*** (Deaths/Cases), while the data on ***Pasteurian Infectivity***, the fraction of people in a population who have been infected (Cases/Population), appears to show a slightly better performance for the Moderna primary vaccine.

The ***Gompertzian Reduction*** (Deaths/Cases) of Moderna compared to Pfizer primary vaccines was 7%, essentially the same.

The ***Pasteurian Reduction*** (Cases/Population) of Moderna compared to Pfizer primary vaccines was 21%.

The ***Malthusian Reduction*** (Deaths/Population) of Moderna compared to Pfizer primary vaccines was 12%.

Thus, the two primary vaccines examined by this study yielded almost the same ***Gompertzian Lethality*** (Deaths/Cases), but slightly different ***Pasteurian Infectivity***. This observation gives us our first hint, which we shall see appearing again and again below, that ***Gompertzian Lethality*** and ***Pasteurian Infectivity*** may well be separate, unlinked, outcomes of vaccination.

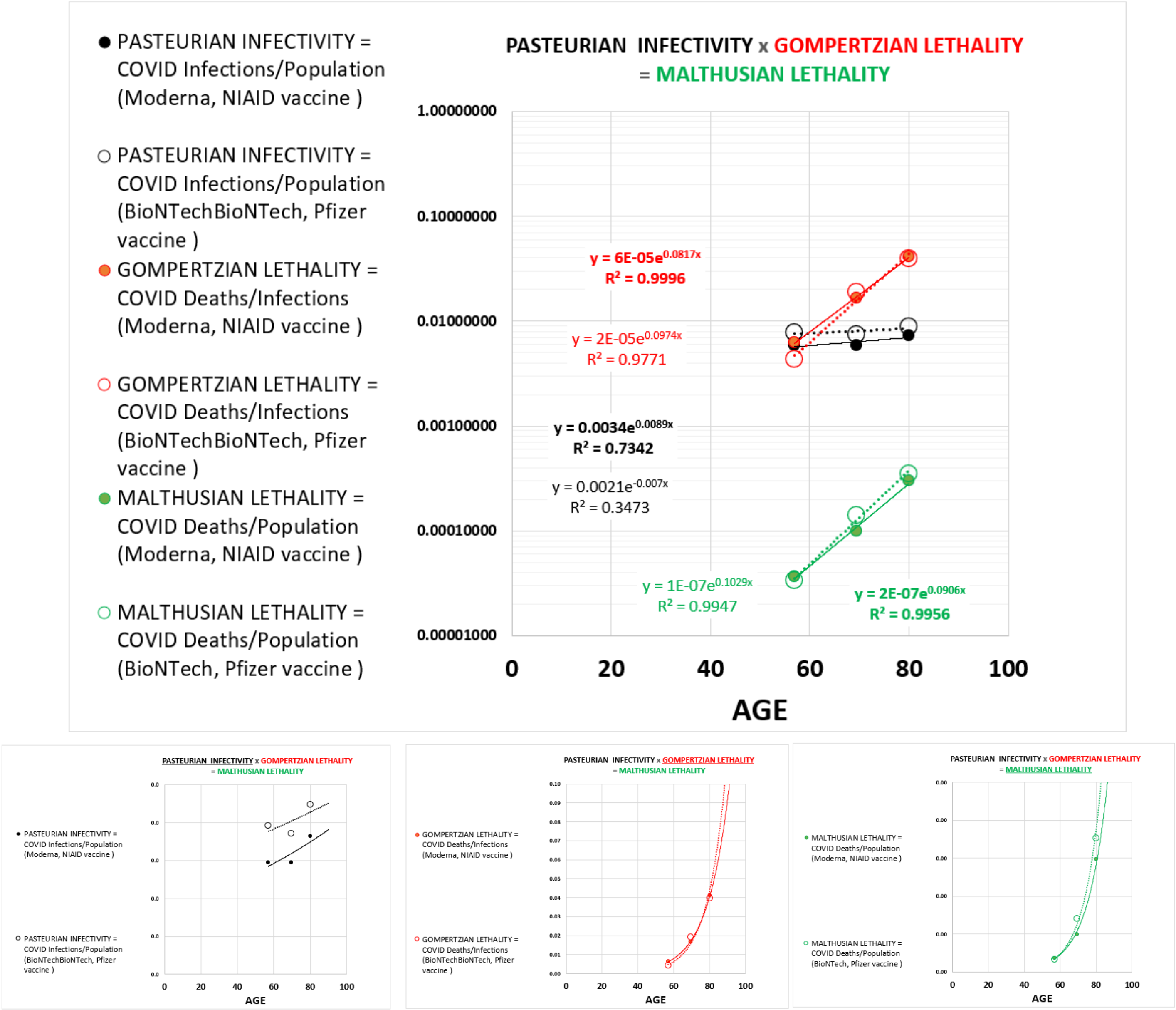

APPENDIX-FIGURE 10

**_Gompertzian Analysis_ of COVID-19** Primary Vaccination: HUNGARY & ARGENTINA, Vaccine Impact & Comparisons

The Hungarian national primary vaccination campaign, with 3.7 million vaccinated individuals, examined the impact of five primary vaccines (Pfizer, Moderna, Sputnik-V, AstraZeneca, Sinopharm), from January 22, 2021, to June 10, 2021, with data reported for finely grained age groups, for both the primary vaccine, and the two boosters, all described in three reports.[34,35,36] Also included in this section is data from a second study, from Argentina, as it also measured the impact of the Sputnik-V primary vaccine.[37] The Sputnik-V primary vaccine has had a troubled history, with rightly criticized analysis, but the results from the Hungary and Argentina studies were so surprising that it seemed wise to lay them out, despite uneasiness with putting too much hope in the certainty of what the data might be telling us.

From the Hungary data, the ***Gompertzian Reduction*** (Deaths/Cases) was 56%.

The ***Pasteurian Reduction*** (Cases/Population) was 87%.

The ***Malthusian Reduction*** (Deaths/Population) was 93%.

Note again how the ***Gompertz Lines*** of the vaccinated and unvaccinated are roughly parallel, indicating roughly equivalent reductions in all age groups (APPENDIX-FIGURES 11 and 12).

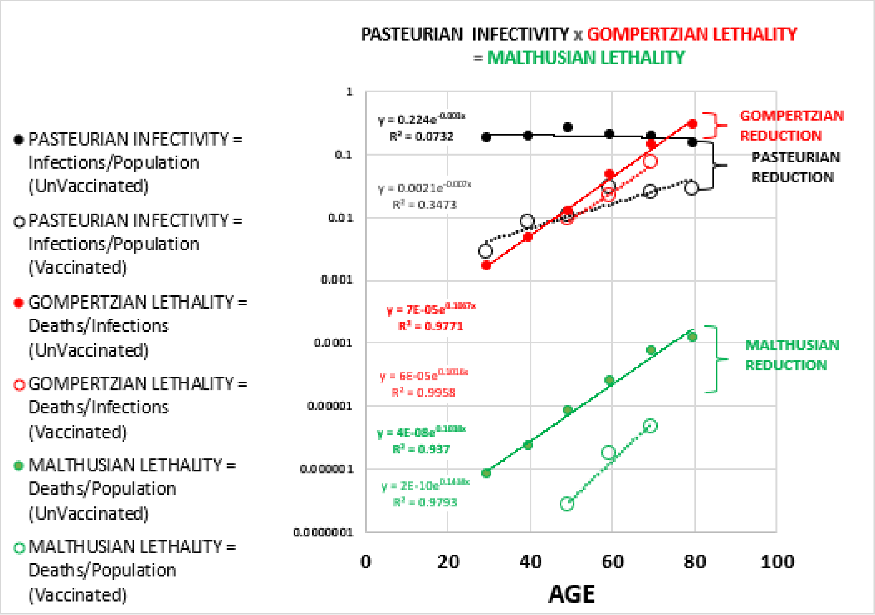

APPENDIX-FIGURE 11

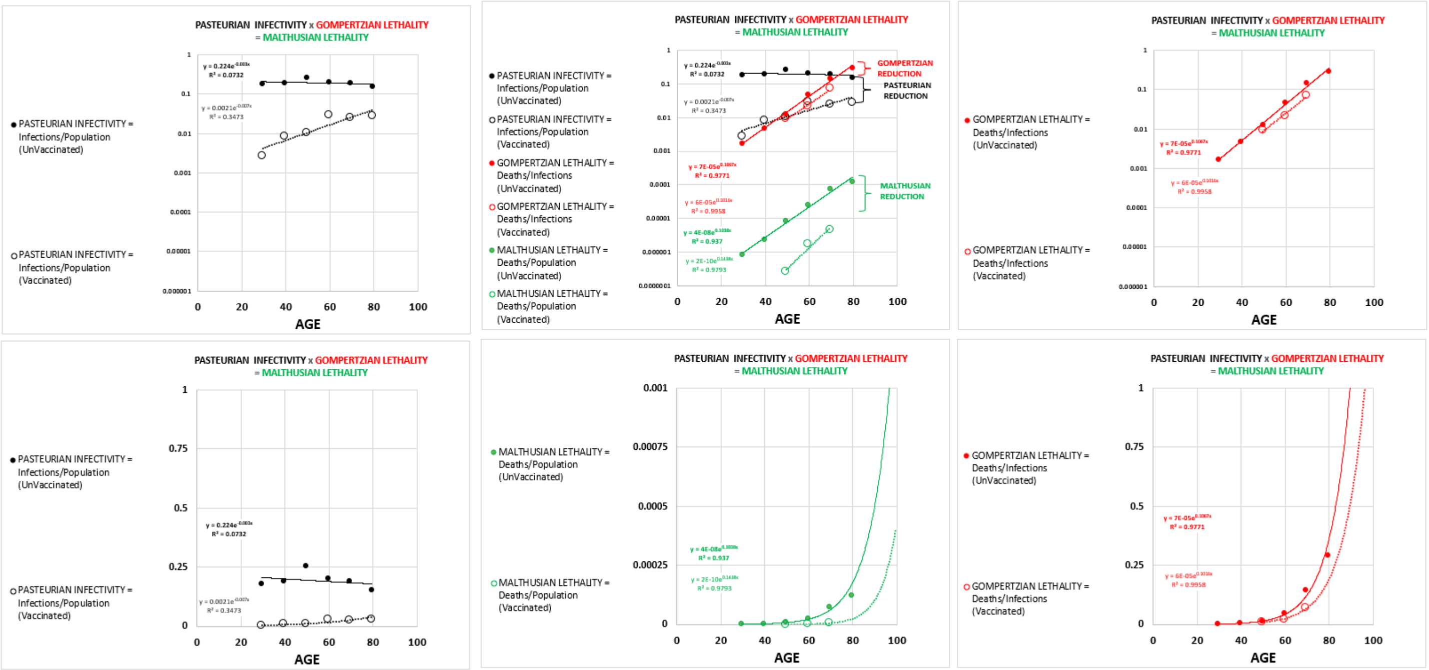

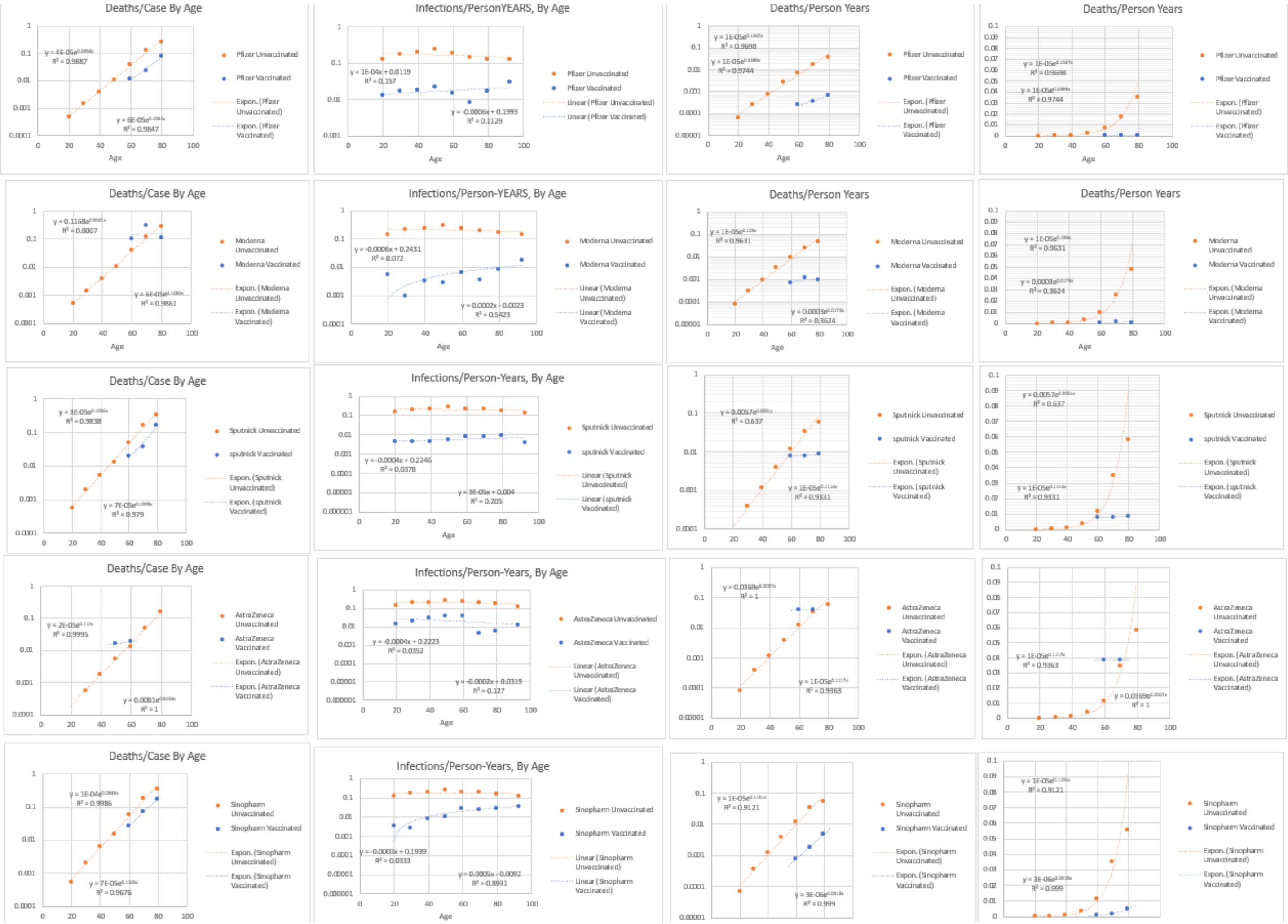

APPENDIX-FIGURE 12

***Gompertz Plots***, revealing log-linear ***Gompertz Lines***, of ***Gompertzian Lethality*** (Deaths/Cases) ***Pasteurian Infectivity*** (Cases/Population) and ***Malthusian Lethality*** (Deaths/Population) for the unvaccinated and vaccinated for each of the primary vaccines, can be seen in the figures above. Again, the ***Gompertz Lines*** of the vaccinated and unvaccinated are roughly parallel, indicating roughly equivalent reductions in all age groups.

Some of the age groups for individual primary vaccines had no deaths, which can't be visualized graphically, since the log of zero has no meaning, and thus has no place to go on a log graph. However, we can use the exponential equation, fit to the data from unvaccinated, to calculate how many deaths would have occurred in each age group IF they had not been vaccinated. We can then add the numbers up, and see, over all ages, how much the reductions were. These calculations can be seen in the APPENDIX-TABLES I and II below.

Perhaps the most striking finding of these calculations was the remarkable benefits of the Sputnik primary vaccine, revealed by these data, especially on ***Gompertzian Lethality***:

- While the most widely used primary vaccine in Hungary, the Pfizer, displayed a ***Gompertzian Reduction***, of 45%, a 1.81-fold reduction in the lethality, the Sputnik primary vaccine displayed a ***Gompertzian Reduction***, of 62%, a 2.63-fold reduction in the lethality.
- The Pfizer vaccine displayed a ***Pasteurian Reduction***, of 90%, a 10.25-fold reduction in ***Infectivity***, while the Sputnik primary vaccine displayed a ***Pasteurian Reduction***, of 99%, a 34.12-fold reduction in the ***Infectivity***.
- In total, this added up to Pfizer primary vaccine having caused a ***Malthusian Reduction***, of 95%, a 10.59-fold reduction in the deaths, while the Sputnik primary vaccine displayed a ***Malthusian Reduction***, of 99%, an 89.67-fold reduction in deaths.

| **Gompertzian Reduction** | Pfizer- | Moderna | Sputnik-V | AstraZeneca | Sinopharm | ALL |
|---|---|---|---|---|---|---|
| Observed CFR/Expected CFR-> | 0.55107 | 0.75426 | 0.3805 | 0.5857 | 0.56567 | 0.55573 |
| +/- 95% -> | 0.0185 | 0.0905 | 0.0423 | 0.0368 | 0.0205 | 0.0123 |
| Reduction, as a % (1-v/un) | 45% | 25% | 62% | 41% | 43% | 56% |
| **#-Fold Reduction in Death, once infected ->** | **1.81** | **1.33** | **2.63** | **1.71** | **1.77** | **1.80** |
| **Pasteurian Reduction** | | | | | | |
| Incidence Vacinated / Incidence UnVacinated -> | 0.0976 | 0.0308 | 0.02931 | 0.15854 | 0.13065 | 0.13065 |
| Reduction, as a % (1-v/un) | 90% | 97% | 97% | 84% | 87% | 87% |
| **#-Fold Reduction in Infection ->** | **10.25** | **32.46** | **34.12** | **6.31** | **7.65** | **7.65** |
| **Malthusian Reduction** (ie Combined) | 0.05378 | 0.02323 | 0.01115 | 0.09286 | 0.0739 | 0.0726 |
| Reduction, as a % (1-v/un) | 95% | 98% | 99% | 91% | 93% | 93% |
| **#-Fold Reduction in Death, Overall ->** | **18.59** | **43.04** | **89.67** | **10.77** | **13.53** | **13.77** |
| **Pasteurian Reduction / Gompertzian Reduction** | **2.01** | **3.94** | **1.57** | **2.03** | **2.00** | **1.56** |

**APPENDIX-TABLE I**

The Hungary data revealed these striking differences between various primary vaccines, in terms of their impact on both ***Gompertzian Lethality*** (Deaths/Cases) and ***Pasteurian Infectivity*** (Cases/Population). This can be seen in the ratio of ***Pasteurian Reduction***/***Gompertzian Reduction***, in the comparison of one primary vaccine to another (**APPENDIX-TABLE I**). Note that some vaccines are twice as powerful as others. Indeed, these two qualities of ***Pasteurian Reduction*** and ***Gompertzian Reduction***, appear to be independent manifestations of **COVID-19** outcome, and the impact which vaccination has on that outcome. Recall that we saw this in the VA(USA) primary vaccine comparison study outlined above. We will see this again in other instances below.

The Hungary data revealing a remarkably superior outcome for the Sputnik primary vaccine, was also seen in a second study of this Sputnik primary vaccine, carried out in Argentina.[37] With data from this Argentinian study, the value of ***Gompertzian Lethality***, that is, Deaths/Cases, by age, could again calculated. These calculations yielded a ***Gompertzian Reduction***, of 62%, a 2.64-fold reduction in the lethality, after the first ***Primary Vaccination Event***, and a ***Gompertzian Reduction***, of 94%, a 15.41-fold reduction in the lethality, after the second dose.

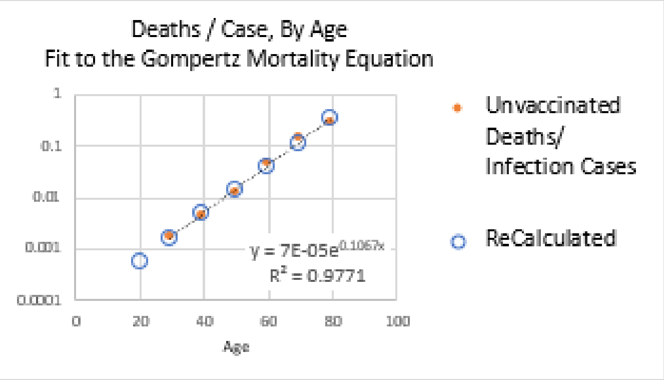

$$y = 7E\text{-}05e^{0.1067x}$$

| | Age | Age | | Pfizer- | Moderna | Sputnik-V | AstraZenec | Sinopharm | ALL |
|---|---|---|---|---|---|---|---|---|---|
| Cases, Among Vaccinated -> | 16–24 | 20 | Cases-> | 89 | 3 | 13 | 6 | 5 | 116 |
| Cases, Among Vaccinated -> | 25–34 | 29.5 | | 253 | 1 | 29 | 16 | 6 | 305 |
| Cases, Among Vaccinated -> | 35–44 | 39.5 | | 391 | 6 | 59 | 79 | 24 | 559 |
| Cases, Among Vaccinated -> | 45–54 | 49.5 | | 610 | 7 | 95 | 310 | 29 | 1,051 |
| Cases, Among Vaccinated -> | 55–64 | 59.5 | | 432 | 19 | 155 | 270 | 456 | 1,332 |
| Cases, Among Vaccinated -> | 65–74 | 69.5 | | 335 | 16 | 131 | 5 | 1,124 | 1,611 |
| Cases, Among Vaccinated -> | 75–84 | 79.5 | | 666 | 35 | 25 | 3 | 593 | 1,322 |
| Total Cases, Among Vaccinated -> | | | | 2,776 | 87 | 507 | 689 | 2,237 | 6,296 |
| | Age | Age | | Pfizer- | Moderna | Sputnik-V | AstraZenec | Sinopharm | ALL |
| Deaths, Among Vaccinated -> | 16–24 | 20 | Deaths -> | 0 | 0 | 0 | 0 | 0 | 0 |
| Deaths, Among Vaccinated -> | 25–34 | 29.5 | | 0 | 0 | 0 | 0 | 0 | 0 |
| Deaths, Among Vaccinated -> | 35–44 | 39.5 | | 0 | 0 | 0 | 0 | 0 | 0 |
| Deaths, Among Vaccinated -> | 45–54 | 49.5 | | 7 | 0 | 0 | 5 | 0 | 12 |
| Deaths, Among Vaccinated -> | 55–64 | 59.5 | | 10 | 2 | 3 | 5 | 12 | 32 |
| Deaths, Among Vaccinated -> | 65–74 | 69.5 | | 26 | 5 | 5 | 0 | 82 | 118 |
| Deaths, Among Vaccinated -> | 75–84 | 79.5 | | 118 | 4 | 4 | 0 | 104 | 230 |
| Total Deaths, Among Vaccinated -> | | | | 161 | 11 | 12 | 10 | 198 | 392 |
| | Age | Age | | Pfizer- | Moderna | Sputnik-V | AstraZenec | Sinopharm | ALL |
| | | | | Expected Cases, in matched indiduals, unvaccinated | | | | | |
| Incidence Vacinated / Incidence UnVacinated -> | | | | 10% | 3% | 3% | 16% | 13% | 13% |
| Expected Cases, if unvaccinated -> | 16–24 | 20 | | 912 | 97 | 444 | 38 | 38 | 1,529 |
| Expected Cases, if unvaccinated -> | 25–34 | 29.5 | | 2,592 | 32 | 990 | 101 | 46 | 3,761 |
| Expected Cases, if unvaccinated -> | 35–44 | 39.5 | | 4,006 | 195 | 2,013 | 498 | 184 | 6,896 |
| Expected Cases, if unvaccinated -> | 45–54 | 49.5 | | 6,250 | 227 | 3,242 | 1,955 | 222 | 11,896 |
| Expected Cases, if unvaccinated -> | 55–64 | 59.5 | | 4,426 | 617 | 5,289 | 1,703 | 3,490 | 15,525 |
| Expected Cases, if unvaccinated -> | 65–74 | 69.5 | | 3,433 | 519 | 4,470 | 32 | 8,603 | 17,057 |
| Expected Cases, if unvaccinated -> | 75–84 | 79.5 | | 6,824 | 1,136 | 853 | 19 | 4,539 | 13,371 |
| Total Expected Cases, if unvaccinated -> | | | | 28,444 | 2,824 | 17,300 | 4,346 | 17,122 | 70,036 |
| | Age | Age | | Pfizer- | Moderna | Sputnik-V | AstraZenec | Sinopharm | ALL |
| | | | 7.00E-05 | Expected Deaths, in matched indiduals, unvaccinated | | | | | |
| | | | IFR | Pfizer- | Moderna | Sputnik-V | AstraZenec | Sinopharm | |
| Expected Deaths, if unvaccinated -> | 16–24 | 20 | 0.0006 | 0 | 0 | 0 | 0 | 0 | 0 |
| Expected Deaths, if unvaccinated -> | 25–34 | 29.5 | 0.0016 | 0 | 0 | 0 | 0 | 0 | 0 |
| Expected Deaths, if unvaccinated -> | 35–44 | 39.5 | 0.0047 | 2 | 0 | 0 | 0 | 0 | 3 |
| Expected Deaths, if unvaccinated -> | 45–54 | 49.5 | 0.0138 | 8 | 0 | 1 | 4 | 0 | 14 |
| Expected Deaths, if unvaccinated -> | 55–64 | 59.5 | 0.0400 | 17 | 1 | 6 | 11 | 18 | 53 |
| Expected Deaths, if unvaccinated -> | 65–74 | 69.5 | 0.1163 | 39 | 2 | 15 | 1 | 131 | 187 |
| Expected Deaths, if unvaccinated -> | 75–84 | 79.5 | 0.3381 | 225 | 12 | 8 | 1 | 201 | 447 |
| Total Expected Deaths, if unvaccinated -> | | | | 292 | 15 | 32 | 17 | 350 | 705 |
| **Gompertzian Reduction** | | | | Pfizer- | Moderna | Sputnik-V | AstraZenec | Sinopharm | ALL |
| Observed CFR/Expected CFR-> | | | | 0.55107 | 0.75426 | 0.3805 | 0.5857 | 0.56567 | 0.55573 |
| | | | +/- 95% -> | 0.0185 | 0.0905 | 0.0423 | 0.0368 | 0.0205 | 0.0123 |
| Reduction, as a % (1-v/un) | | | | 45% | 25% | 62% | 41% | 43% | 56% |
| **#-Fold Reduction in Death, once infected ->** | | | | **1.81** | **1.33** | **2.63** | **1.71** | **1.77** | **1.80** |
| **Pasteurian Reduction** | | | | | | | | | |
| Incidence Vacinated / Incidence UnVacinated -> | | | | 0.0976 | 0.0308 | 0.02931 | 0.15854 | 0.13065 | 0.13065 |
| Reduction, as a % (1-v/un) | | | | 90% | 97% | 97% | 84% | 87% | 87% |
| **#-Fold Reduction in Infection -** | | | | **10.25** | **32.46** | **34.12** | **6.31** | **7.65** | **7.65** |
| **Malthusian Reduction** (ie Combined) | | | | 0.05378 | 0.02323 | 0.01115 | 0.09286 | 0.0739 | 0.0726 |
| Reduction, as a % (1-v/un) | | | | 95% | 98% | 99% | 91% | 93% | 93% |
| **#-Fold Reduction in Death, Overall ->** | | | | **18.59** | **43.04** | **89.67** | **10.77** | **13.53** | **13.77** |

APPENDIX-TABLE II

***Gompertzian Analysis*** of **COVID-19** Primary Vaccination: ISRAEL, Vaccine Impact

Israel engaged in a nationwide primary vaccination program (Pfizer) in December 2020. A number of excellent studies, quite comparable, examined the impact of the primary vaccine in the first months of 2021. For our purposes here, Goldberg et al[38] is more relevant, with data from December 20, 2020, to March 20, 2021 (APPENDIX-FIGURE 13). Note yet again how the ***Gompertz Lines*** of the vaccinated and unvaccinated are roughly parallel, indicating roughly equivalent reductions in all age groups.

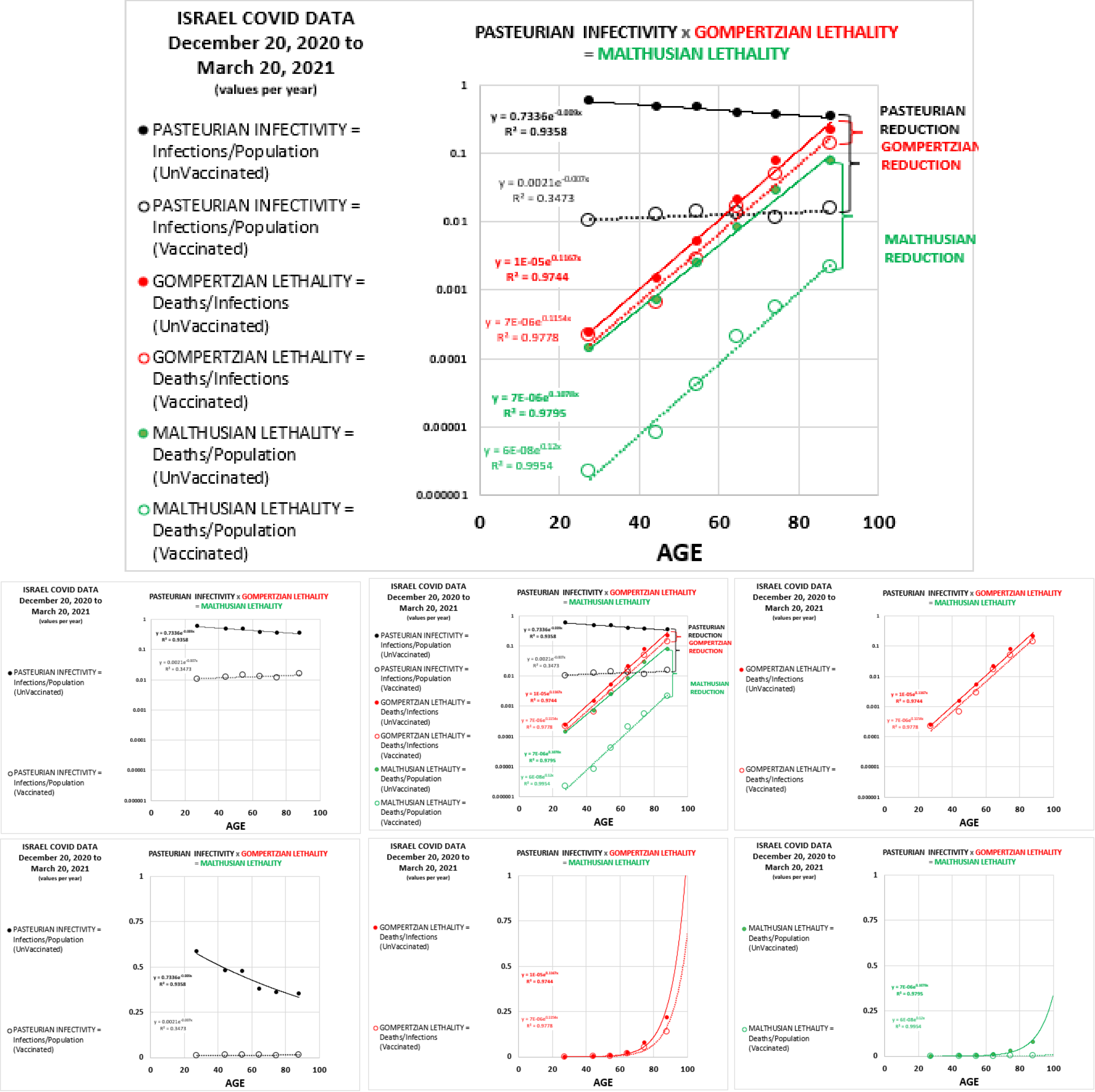

APPENDIX-FIGURE 13

The ***Gompertzian Reduction*** (Deaths/Cases) of vaccinated compared to unvaccinated was 41%.
The ***Pasteurian Reduction*** (Cases/Population) of vaccinated compared to unvaccinated was 97%.
The ***Malthusian Reduction*** of vaccinated compared to unvaccinated was 98%.

Goldberg et al went further to examine cohorts of patients, differing by the amount of time since the first and second primary doses, and 384 individuals with **COVID-19** infection who had been previous infected, among which there was only one death. These data from these cohorts also reveal that ***Gompertzian Reduction*** (Deaths/Cases) improved for first to second doses, and with time. Remarkably, the previously infected individuals had ***Gompertzian*** and ***Pasteurian Reductions*** from the infection that endowed them with ***Gompertzian*** and ***Pasteurian Reductions*** in future infections that were as good as, and perhaps even better than, the benefit of primary vaccination seen in individuals who had never been infected (APPENDIX-FIGURE 14).

| | Cohort 0: unvaccinated and not previously infected. | Cohort 1A: followed from day 1 to day 14 after the first vaccine dose. | Cohort 1B: followed from 15 days after the first dose to 13 days after the second dose. | Cohort 2: followed from 14 days after the second dose onwards. | Recovered cohort: previously infected individuals. |
|---|---|---|---|---|---|
| **Gompertzian Reduction**: 1- (Actual # of Deaths/Expected # of Deaths [for Age-Matched, Unvaccinated, Individuals]) | 4% | 36% | 39% | 50% | 83% |
| **X-Fold** Reduction in Lethality by Vaccination or Previous Disease | 1.04 | 1.55 | 1.64 | 2.01 | 5.99 |

**<u>APPENDIX-TABLE III</u>**

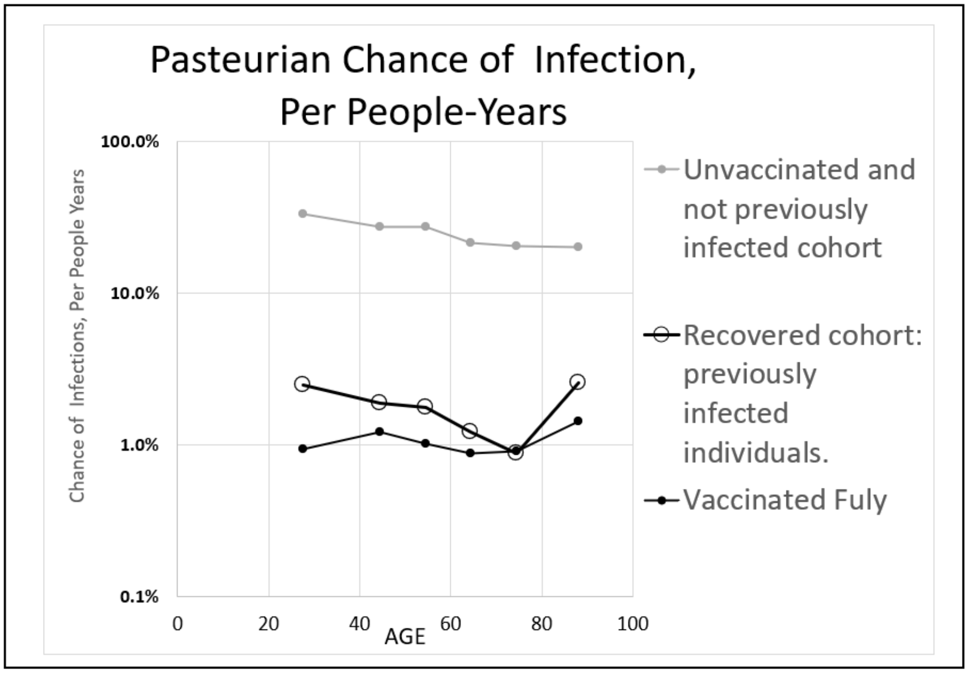

**<u>APPENDIX-FIGURE 14</u>**

***Gompertzian Analysis*** of **COVID-19** Vaccination: Population Comparisons

A comparison of the results of the ***Gompertzian Analysis*** of data on **COVID-19** vaccination from each of the four studies described above (Israel, 12/20/21-3/20/21; Hungary, 1/22/21-6/10/21; USA(CDC), 4/1/21-6/20/22; VA(USA), 12/11/20-6/30/21) gives us our first look at how ***Gompertzian Lethality*** (Deaths/Cases) ***Pasteurian Infectivity*** (Cases/Population) and ***Malthusian Lethality***, (Deaths/Population) may vary independently, and change over time. (APPENDIX-FIGURE 15, APPENDIX-TABLE IV).

In the Israel population, the reduction in **COVID-19** ***Infectivity***, measured by ***Pasteurian Reduction*** (Cases/Population) was much stronger than the reduction in **COVID-19** ***Lethality***, measured by ***Gompertzian Reduction*** (Deaths/Cases).

In contrast, in the USA(CDC), population, the reduction in **COVID-19** ***Lethality***, the ***Gompertzian Reduction*** (Deaths/Cases) was much closer to the reduction in **COVID-19** ***Infectivity***, the ***Pasteurian Reduction*** (Cases/Population).

The Hungary and VA(USA) populations fell in between.

These values suggest, again, that change in **COVID-19** ***Gompertzian Lethality*** measured by the ***Gompertzian Reduction*** of Deaths/Cases, can be independent of change in **COVID-19** ***Pasteurian Infectivity*** measured by the ***Pasteurian Reduction*** of Cases/Population, even though vaccination is at work on both effects.

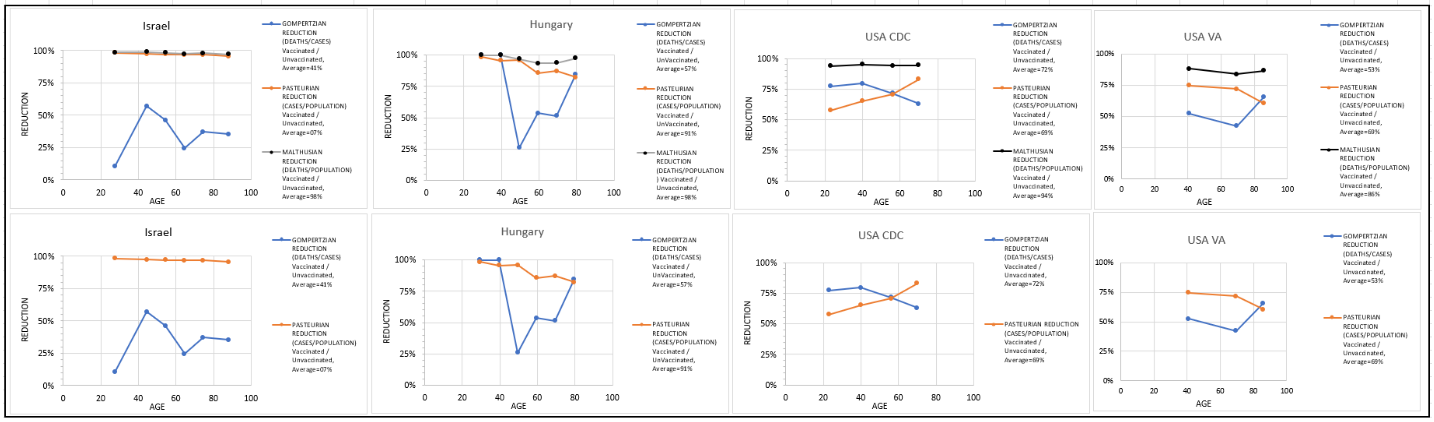

APPENDIX-FIGURE 15

| STUDY | GOMPERTZIAN REDUCTION (CASES / DEATHS) “1-(vaccinated / unvaccinated)” | PASTEURIAN REDUCTION (CASES / POPULATION) “1-(vaccinated / unvaccinated)” | MALTHUSIAN REDUCTION (DEATHS / POPULATION) “1-(vaccinated / unvaccinated)” | GOMPERTZIAN / PASTEURIAN |
|---|---|---|---|---|
| Israel, 12/20/21-3/20/21 | 41% | 97% | 98% | 0.43 |
| Hungary, 1/22/21-6/10/21 | 56% | 87% | 93% | 0.64 |
| USA(CDC), 4/1/21-6/20/22 | 73% | 83% | 94% | 0.88 |
| VA(USA), 12/11/20-6/30/21 | 53% | 69% | 87% | 0.78 |

APPENDIX-TABLE IV

***Gompertzian***, ***Pasteurian***, and ***Malthusian Reductions***, of four studies, each an average of the age groups.

***Gompertzian Analysis*** of **COVID-19** Vaccination: USA 2021-2022, impact over time.

Let us now move on from examining **COVID-19** datasets as single chunks of data, to examining data assembled to capture how **COVID-19**'s impact has changed over time. We begin by returning to the rather extraordinary CDC data on approximately 70% of all **COVID-19** cases in the USA. The first dataset, from April 2021 to June 2022, contains data on patients sorted by vaccination status, but no booster information. Thus, we have data over the period from three months after the first primary vaccines became available in late December 2021, beginning when the alpha variant was predominant, and then followed by delta in ~August 2021, and then omicron in ~December 2021+. During this period, the first boosters were approved in late September 2021, then the second boosters at the end of March 2022, and then the third boosters in September 2022. This dataset didn't contain booster data, but a second dataset does, and its detailed analysis will be discussed below (See: ***Gompertzian Analysis*** of the impact of the number of **COVID-19 *Vaccination Events*** (Primary & Boosters).

At the beginning of the first CDC dataset, April 2021, about 25% of Americans were vaccinated, and thus about 75% unvaccinated. By the end of this dataset, June 2022, the reverse was the case, with about 75% of Americans being vaccinated and about 25% unvaccinated (APPENDIX-FIGURE 16).

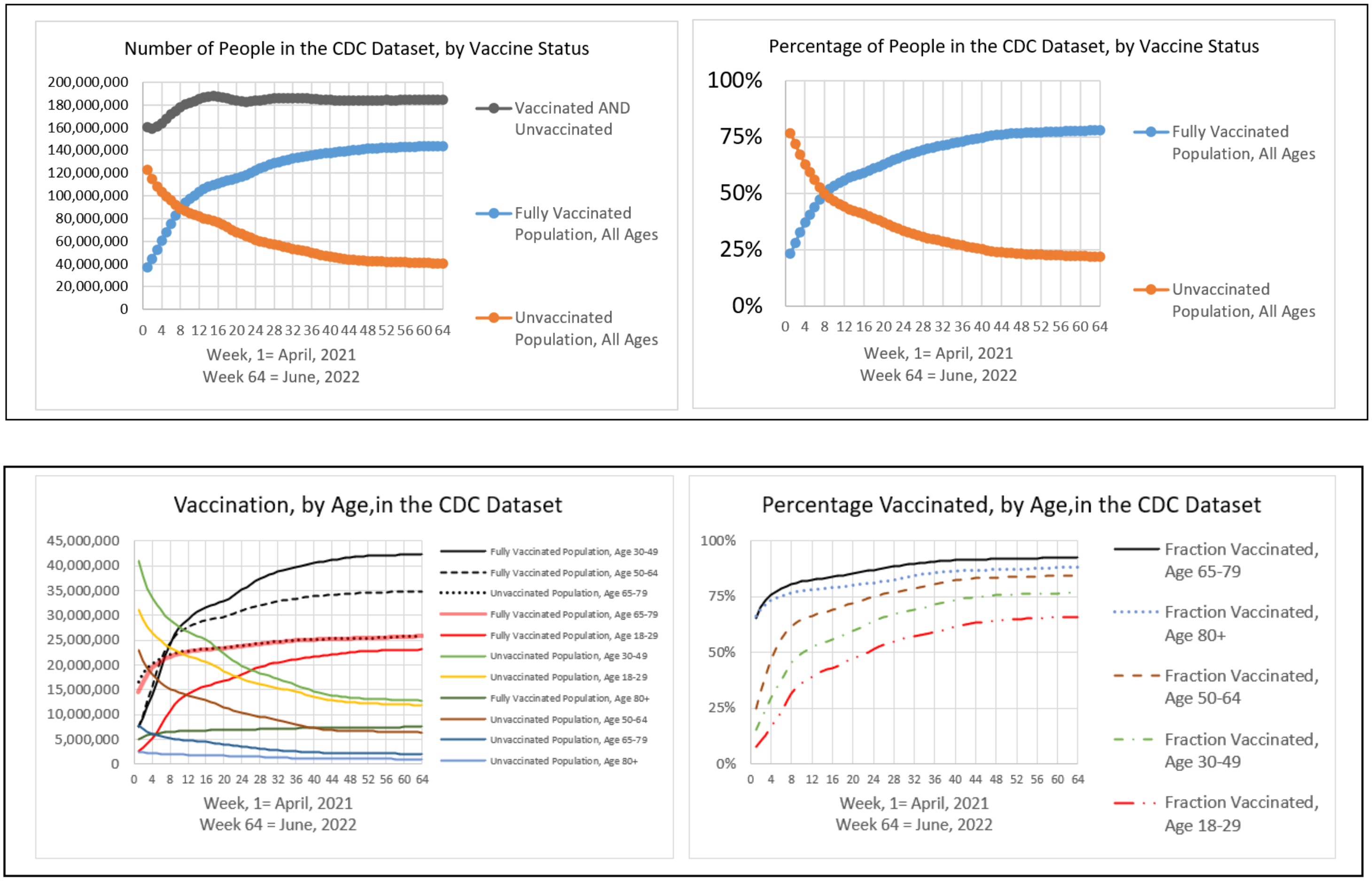

APPENDIX-FIGURE 16

In parallel, in the first dataset, from April 2021 the percentage of **COVID-19** cases occurring among the vaccinated rose from about 25% of all cases to about 75% by June 2022 (APPENDIX-FIGURE 17).

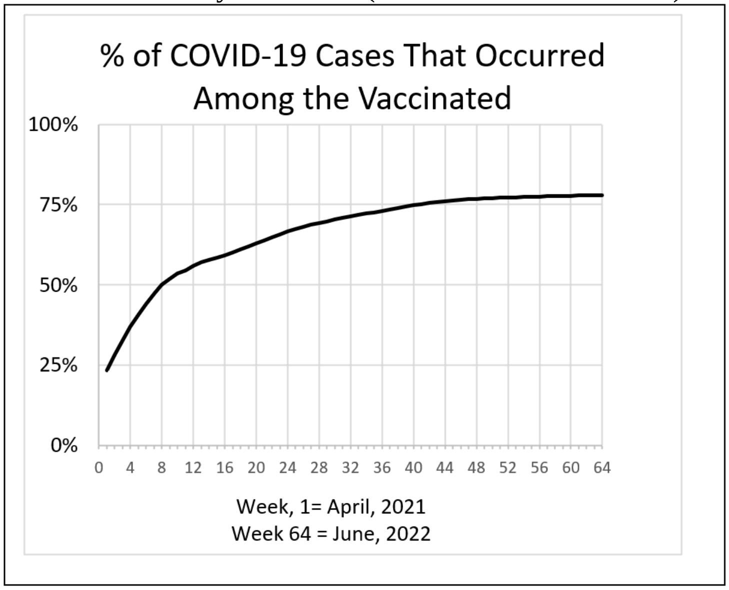

APPENDIX-FIGURE 17

Vaccinated patients had a markedly lower rate of death, with about 70,000 lives lost among the vaccinated from April 2021 to June 2022, and about 180,00 lives lost among unvaccinated during this time period (APPENDIX-FIGURE 18). In terms of the populations of the vaccinated and unvaccinated individuals, the unvaccinated had about six times the frequency of death.

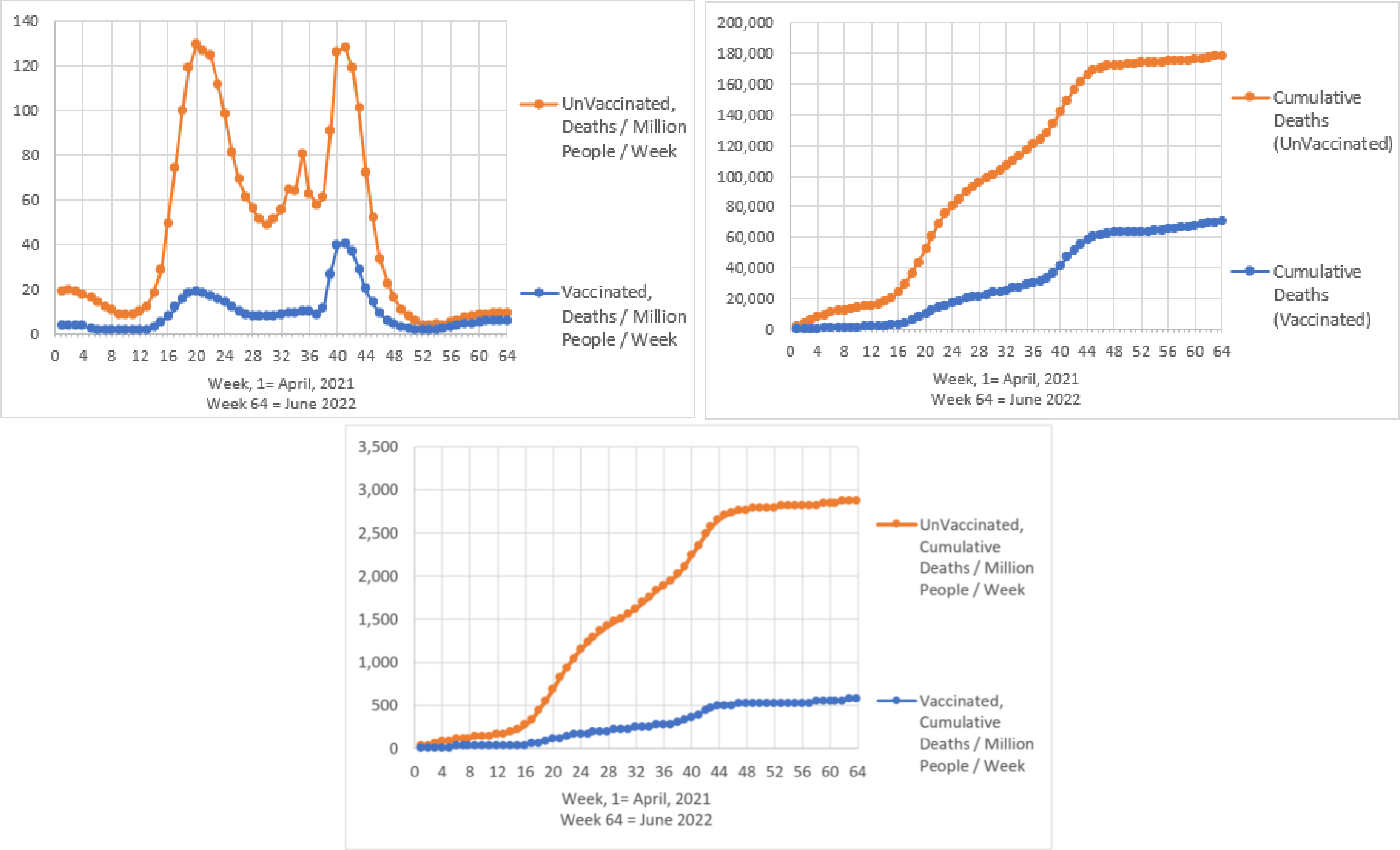

APPENDIX-FIGURE 18

***Gompertzian Analysis*** of **COVID-19** vaccination, over time, by age.

In APPENDIX-FIGURE 19 are shown values for ***Pasteurian Infectivity*** (Cases/Population) and ***Gompertzian Lethality*** (Deaths/Cases), for the vaccinated and unvaccinated, for each week, from April 2021 to June 2022. The Y-Axis for these figures is on a logarithmic scale, in units of 10, so a dot that is twice as high as another dot captures a 10-fold higher value, and a dot that's three times as high captures a 100-fold higher value, etc. The series of dots form wave-like images, from April 2021 on the left of each graph, to June 2022 on the right, visualizing how ***Pasteurian Infectivity*** (Cases/Population) and ***Gompertzian Lethality*** (Deaths/Cases) changed over time, for the vaccinated, and the unvaccinated. There is a great deal of information in these figures, which we shall examine, item by item:

***Pasteurian Infectivity*** went up and down wildly in 2021-2022, ending 10-fold higher than at the beginning.

For both vaccinated and unvaccinated, ***Pasteurian Infectivity*** (Cases/Population) went up and down wildly with time, with the huge waves, varying over a hundred+-fold difference in height, and displaying, over the long term, a roughly a 10-fold increase in the chance of infection from April 2021 to June 2022 (APPENDIX-FIGURE 19). Vaccinated people had lower rates of ***Pasteurian Infectivity*** (Cases/Population) than unvaccinated people, but the virus kept defeating the vaccines.

***Gompertzian Lethality*** declined progressively in 2021-2022, ending 10-fold lower than at the beginning.

For both vaccinated (APPENDIX-FIGURE 20, right) and unvaccinated (APPENDIX-FIGURE 20, left), ***Gompertzian Lethality*** (Deaths/Cases) went up and down mildly with time, with small waves that varied several-fold in height. However, over the long term, ***Gompertzian Lethality*** declined roughly 10-fold from April 2021 to June 2022.

Graphing ***Gompertzian Lethality*** (Deaths/Cases), sorted by age, among the vaccinated, over time, on a linear scale, reveals all age groups pointing down to minimal values (APPENDIX-FIGURE 20).

Precisely why ***Gompertzian Lethality*** (Deaths/Cases) among the vaccinated declined over this 15-month period is not contained in these data. Boosters, infections, new variants, or other forces could be behind these changes. However, as we shall see below, using a second dataset with booster information, it was found that boosters are likely to be a major force in lessening ***Gompertzian Lethality*** (Deaths/Cases).

Among the unvaccinated, the decline in ***Gompertzian Lethality*** (Deaths/Cases) was most likely driven by immunization by infections, at the cost of lives lost. We shall return to this sad process below.

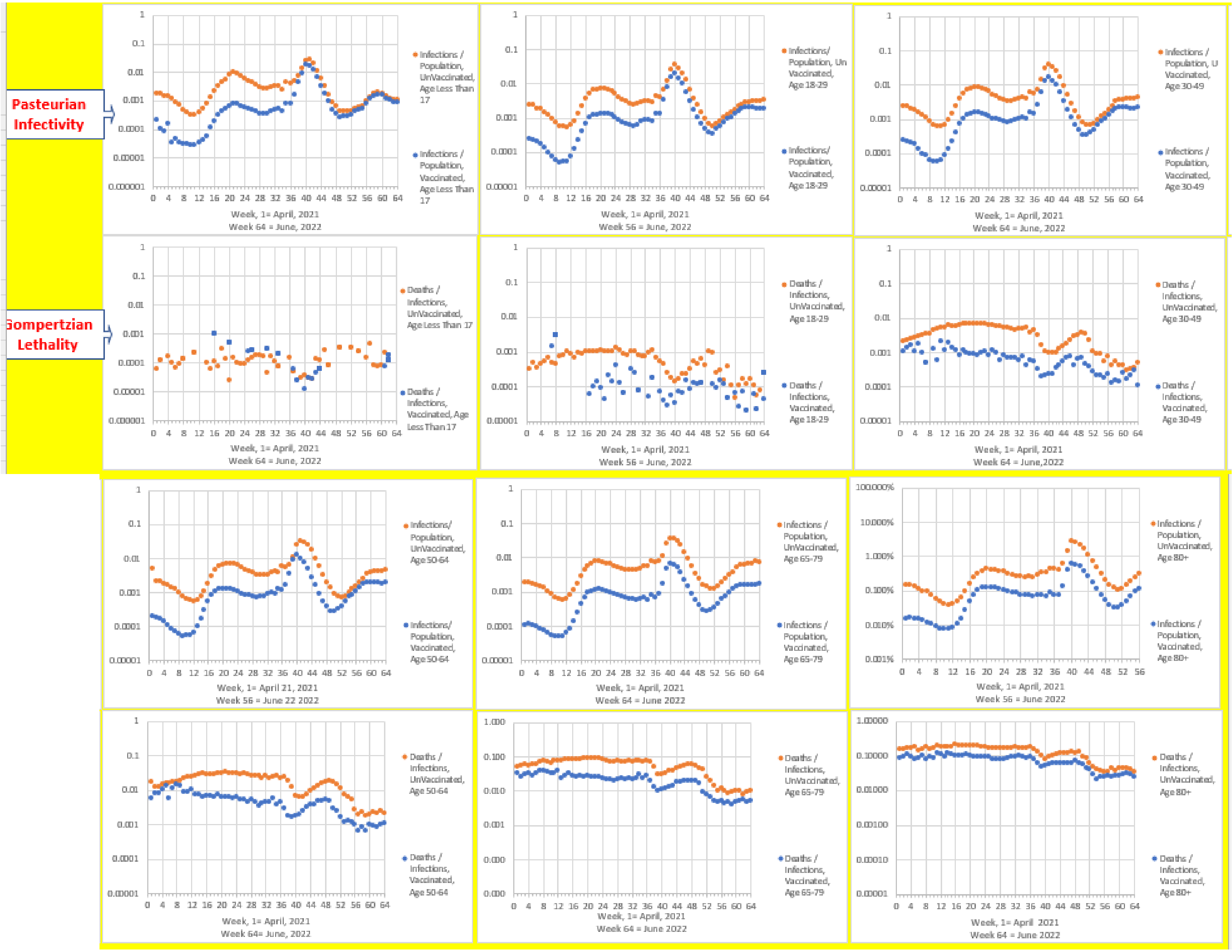

APPENDIX-FIGURE 19

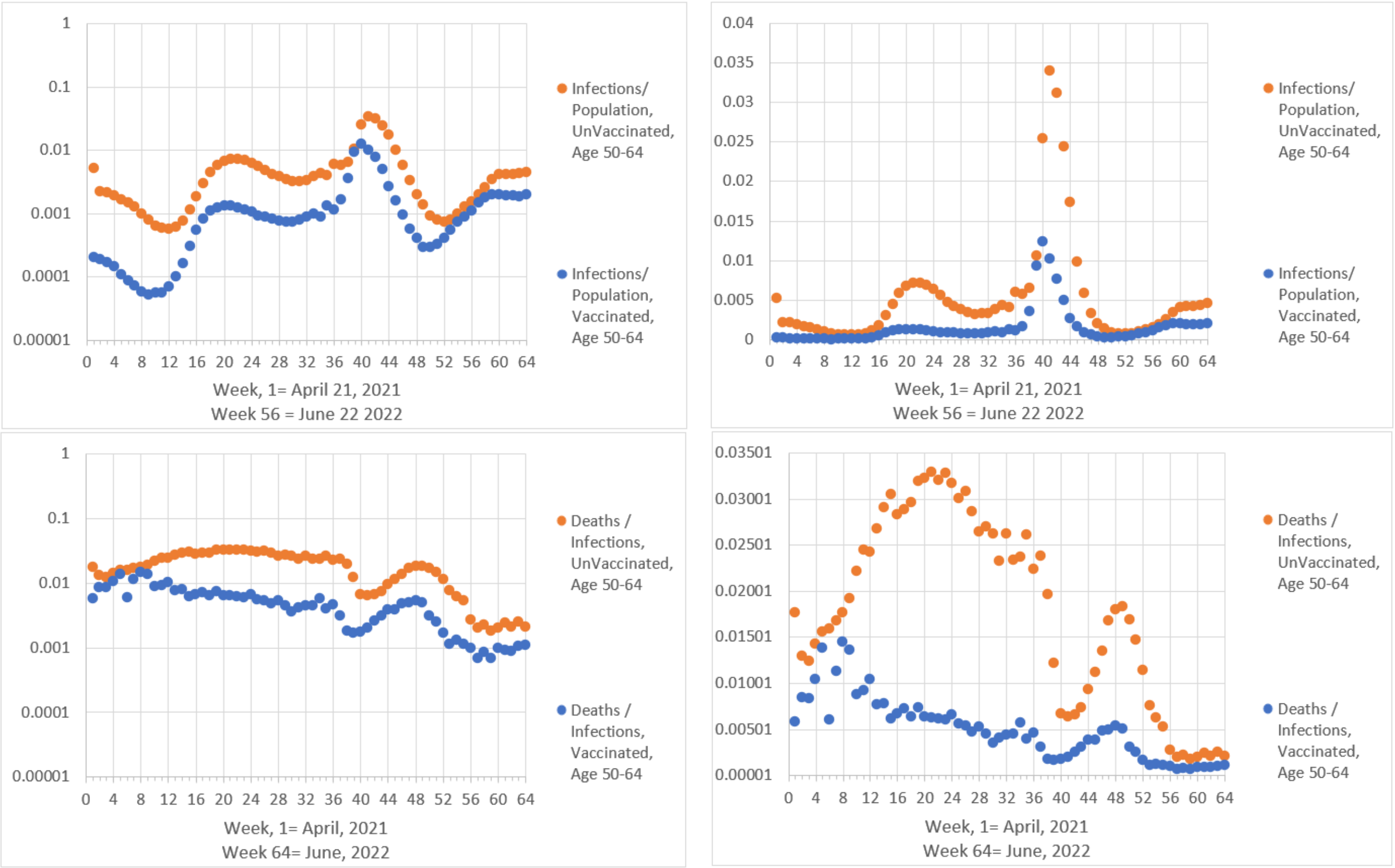

APPENDIX-FIGURE 19b

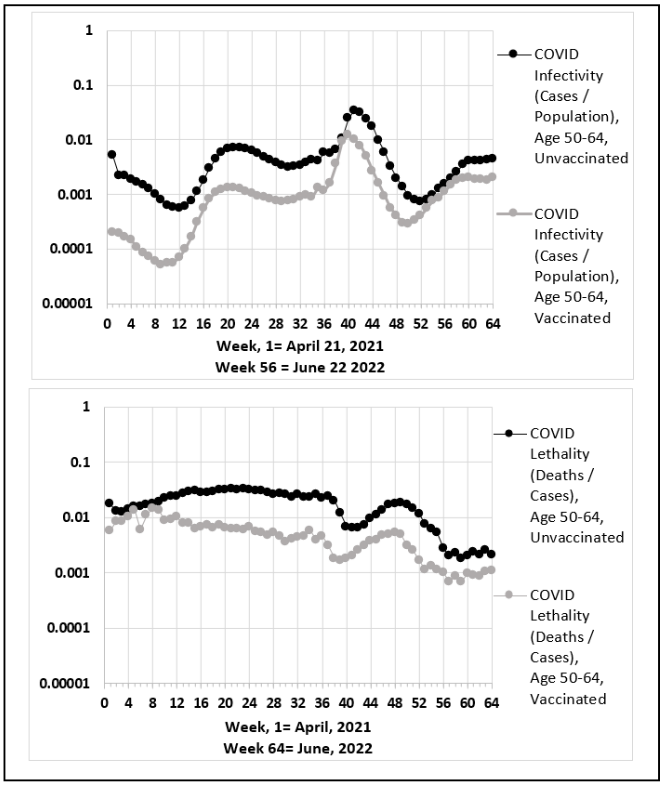

APPENDIX-FIGURE 19c. **(FIGURE 4)**

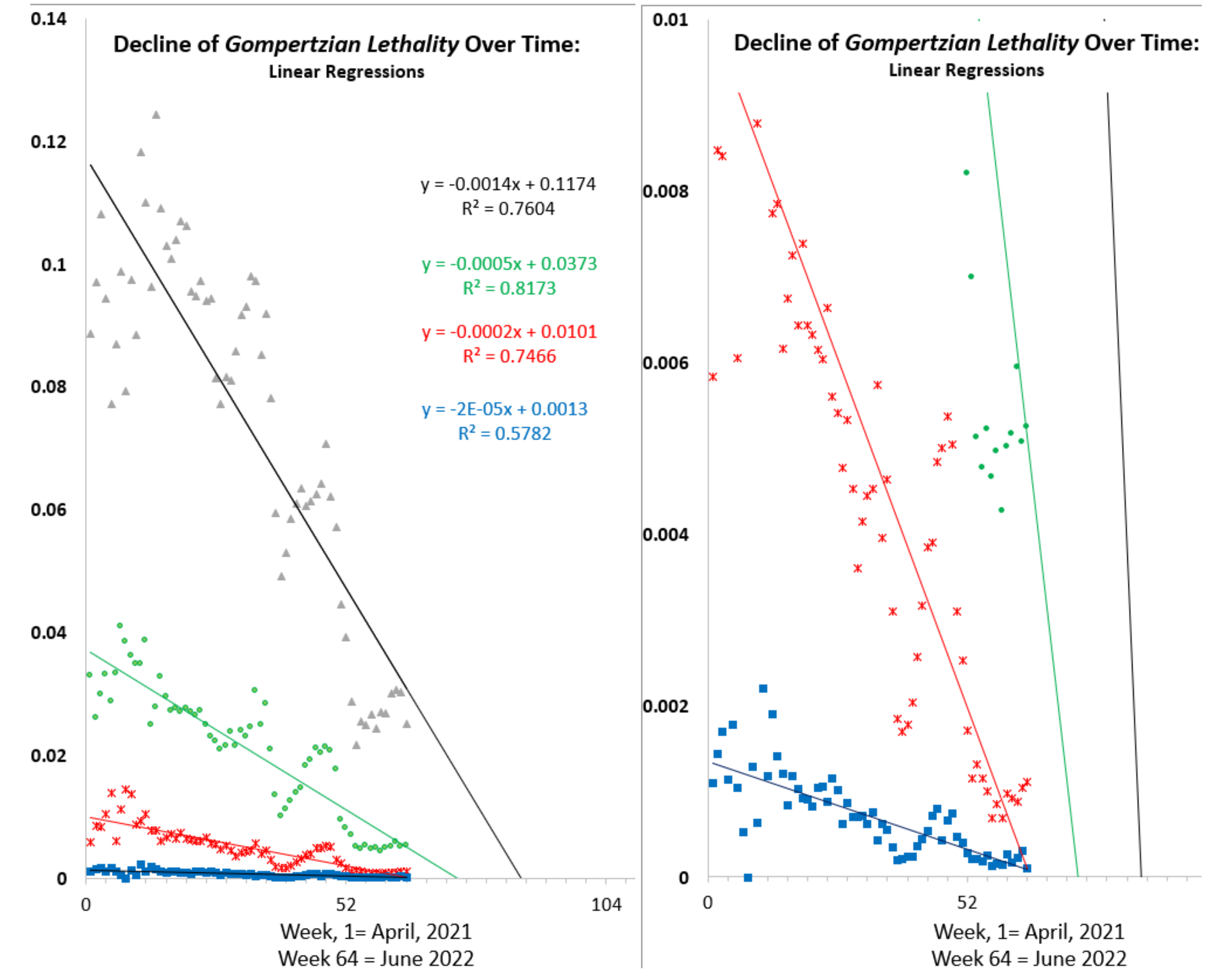

APPENDIX-FIGURE 20. ***Gompertzian Lethality*** (Deaths/Cases), over time.
Unvaccinated, left; Vaccinated, right. Grey age 80+, Green age 65-70, red age 50-64, blue age 30-49

<u>The 2021-2022 decline in ***Gompertzian Lethality*** among the vaccinated declined in ***Gompertzian Height***, not ***Slope***.</u>

In APPENDIX-FIGURE 21, are shown **COVID-19** ***Infectivity*** (Cases/Population) and **COVID-19** ***Lethality*** (Deaths/Cases) for the vaccinated, in the 65-79 age group. (Similar results were found for all of the other age groups.) Graphs of the log of ***Gompertzian Lethality*** (Deaths/Cases) vs (age), for monthly groups of vaccinated **COVID-19** patients revealed ***Gompertz Lines*** of ***Gompertzian Lethality***, declining progressively in ***Gompertzian Height***, with little evident change in the ***Gompertzian Slope*** (APPENDIX-FIGURE 21). That is to say, ***Gompertzian Lethality*** declines by roughly equivalent improvement across all ages.

Again, boosters and breakthrough infections are likely to be the forces behind this 10-fold decline in ***Gompertzian Lethality*** (Deaths/Cases).

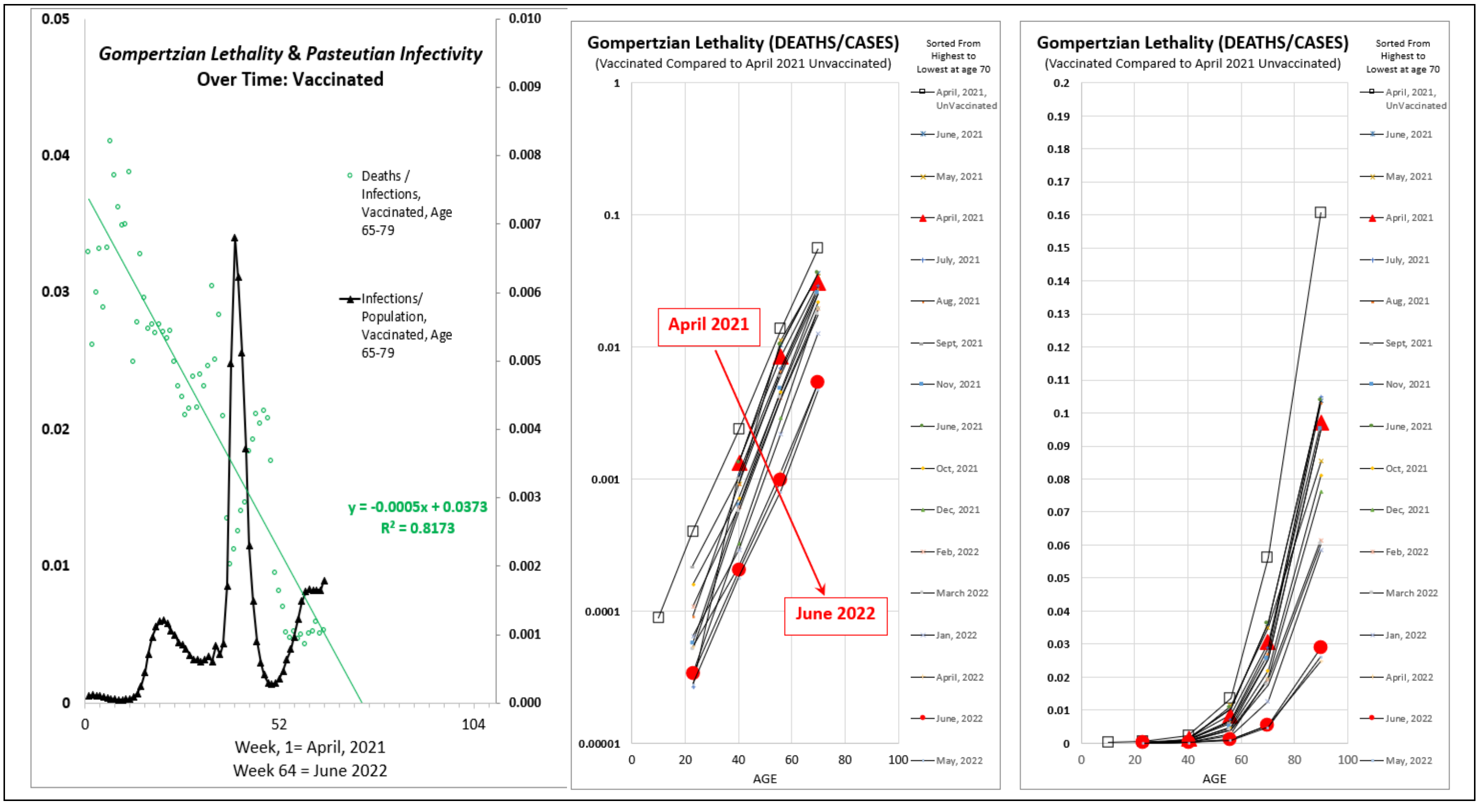

APPENDIX-FIGURE 21

<u>The 2021-2022 decline in ***Gompertzian Lethality*** of the unvaccinated declined in ***Gompertzian Height***, not ***Slope***.</u>

In APPENDIX-FIGURE 22, are shown **COVID-19** ***Infectivity*** (Cases/Population) and **COVID-19** ***Lethality*** (Deaths/Cases) for the unvaccinated, in the 65-79 age group. (Similar results were found for all of the other age groups.) Graphs of the log of ***Gompertzian Lethality*** (Deaths/Cases) vs (age), for monthly groups of unvaccinated **COVID-19** patients revealed ***Gompertz Lines*** of ***Gompertzian Lethality***, declining progressively in ***Gompertzian Height***, with little evident change in the ***Gompertzian Slope*** (APPENDIX-FIGURE 22). That is to say, ***Gompertzian Lethality*** declines by roughly equivalent improvement across all ages.

Thus, the unvaccinated also underwent a similar, but smaller, decline in ***Gompertzian Lethality*** as that seen among the vaccinated (APPENDIX-FIGURE 21), but they never caught up with the vaccinated (APPENDIX-FIGURE 22). Perhaps this decline ***Gompertzian Lethality*** among the unvaccinated was created by **COVID-19** infection, as we saw above from the Israel cohort of recovered **COVID-19** individuals (APPENDIX-TABLE III). If so, the sad price for the reduction in the ***Gompertzian Lethality*** among the unvaccinated was the loss of life that occurred after some of these infections; by June 2022, unvaccinated Americans were ¼th of the population in this dataset (APPENDIX-FIGURE 16) but had 2½ times more deaths than the vaccinated (APPENDIX-FIGURE 22 right)

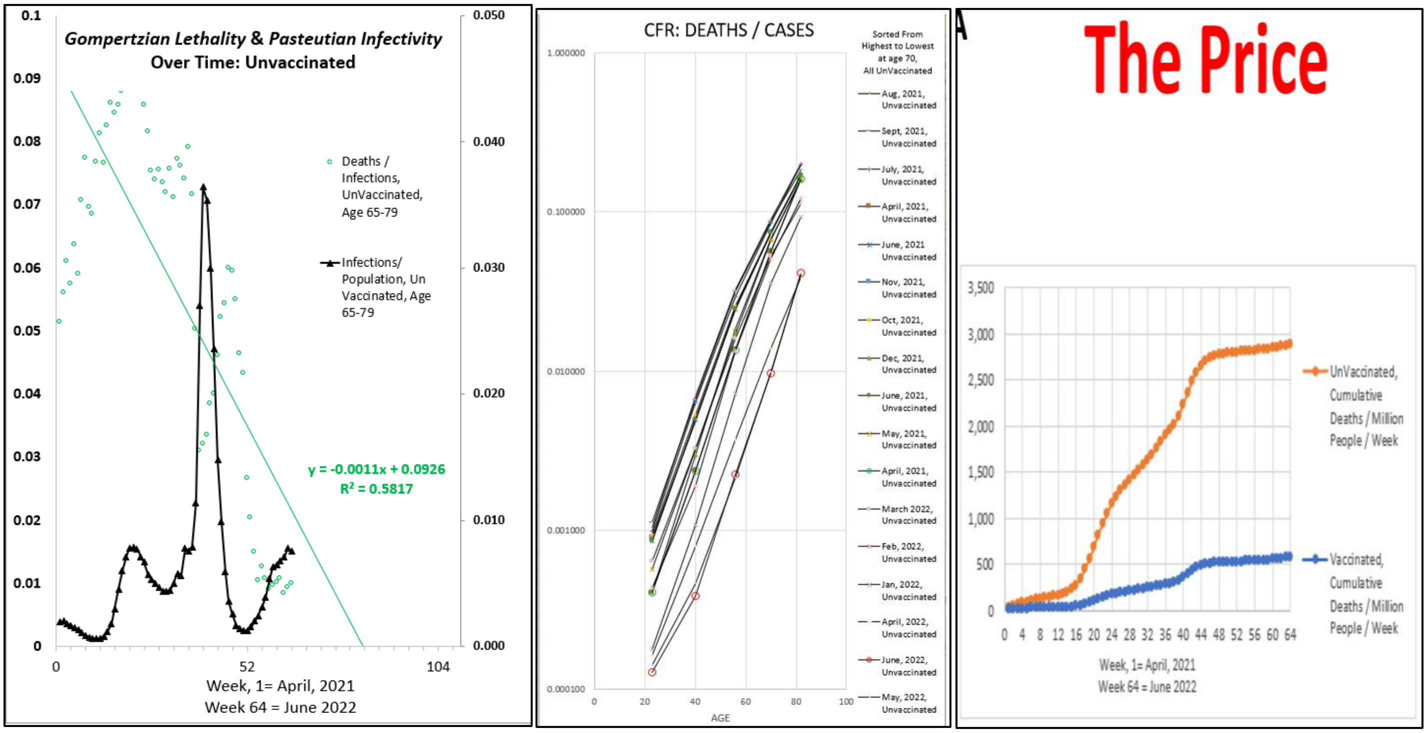

**<u>APPENDIX-FIGURE 22</u>**

**COVID-19** Infection, like vaccination, pushes down the ***Gompertzian Height*** of ***Gompertzian Lethality***.

A direct measure of the impact of primary vaccination, and infection, on the reduction in **COVID-19** ***Lethality*** (Deaths/Cases) emerged from a study of Israel's nationwide primary vaccination program by Goldberg et al[39], as described above, who examined data from December 20, 2020, to March 20, 2021 (APPENDIX-TABLE III, APPENDIX-FIGURE 14). From these data, calculations of the average reduction in **COVID-19** ***Lethality*** (Deaths/Cases) of vaccinated compared to unvaccinated, averaged over all of the age groups, was 41%. Goldberg et al went further to examine cohorts of patients, differing by the amount of time since the first and second dose, and 384 individuals with **COVID-19** infection who had been previous infected, among which there was only one death. These data from these cohorts also reveal that ***Gompertzian Reduction*** of **COVID-19** ***Lethality*** (Deaths/Cases) improved for first to second doses, and with time. Remarkably, the group of previously infected individuals had **COVID-19** ***Lethality Reductions*** from the infection that endowed them with **COVID-19** ***Lethality*** levels in future infections that were as good as, and perhaps even better than, the benefit of primary vaccination seen in those who had never been infected (APPENDIX-TABLE III, APPENDIX-FIGURE 14).

**COVID-19** Infection, and its impact on ***Gompertzian Lethality***, give us a chance to learn how to schedule boosters.

Data on infection's impact on **COVID-19** ***Lethality*** (Deaths/Cases) is an unexploited opportunity to gain insight into how the number and timing of ***Vaccination Events*** can best reduces the chance of death. Boosters, whose sequential impact on **COVID-19** ***Lethality*** we shall examine below, have been given in limited numbers and at limited intervals. Infections occur in some unfortunate individuals in large numbers and at a great variety of intervals. Extracting essential information about how the number of these infections, and their timing, affect **COVID-19** ***Lethality***, should tell us much useful information about how we can get the most out of future boosters.

***Gompertzian Analysis*** of the impact of the number of **COVID-19 *Vaccination Events*** (Primary & Boosters)

Let us now examine how **COVID-19** changes with sequential vaccination.

I. ISRAEL, Boosters

Studies from Israel provides valuable data in the impact of 1st and 2nd boosters on **COVID-19 *Gompertzian Lethality*** (Deaths/Cases) and ***Malthusian Lethality*** (Deaths/Population). The first study captured data on deaths and cases by age, making ***Gompertzian Analysis*** of ***Gompertzian Lethality*** (Deaths/Cases) possible.[40]

***Gompertzian Lethality*** (Deaths/Cases), by age, on ***Gompertz Plots***, can be seen in APPENDIX-FIGURE 23.

***Gompertzian Reductions***, relative to the unvaccinated, for patients with just primary vaccination (2 Doses), 1 Booster (3 Doses) or 2 Boosters (4 Doses), can be seen in APPENDIX-FIGURE 24.

***Gompertzian Lethality*** (Deaths/Cases), for each age group, by number of ***Vaccination Events*** can be seen in APPENDIX-FIGURE 25: unvaccinated ("0" on the X-axis), vaccinated with just the primary ("1" on the X-axis), vaccinated with 1 Booster ("2" on the X-axis), and vaccinated with 2 Boosters ("3" on the X-axis).

All of these calculations show that with each booster, ***Gompertzian Lethality*** declines, again pointing to very low lethality at ~3 or 4 boosters, as we shall see below for the USA CDC and Hungarian booster data. The quantification of this decline will be examined below.

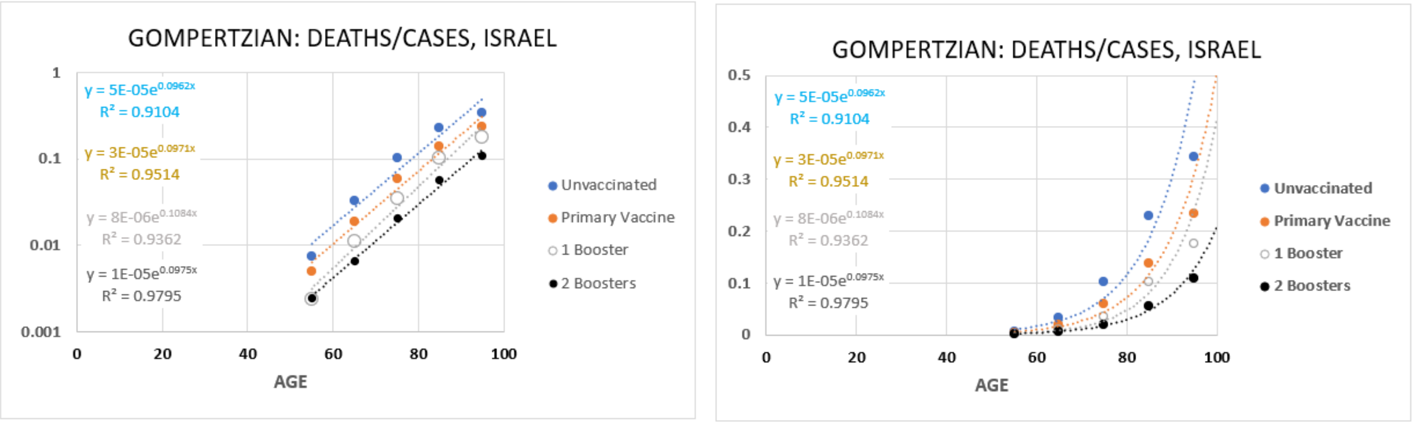

APPENDIX-FIGURE 23

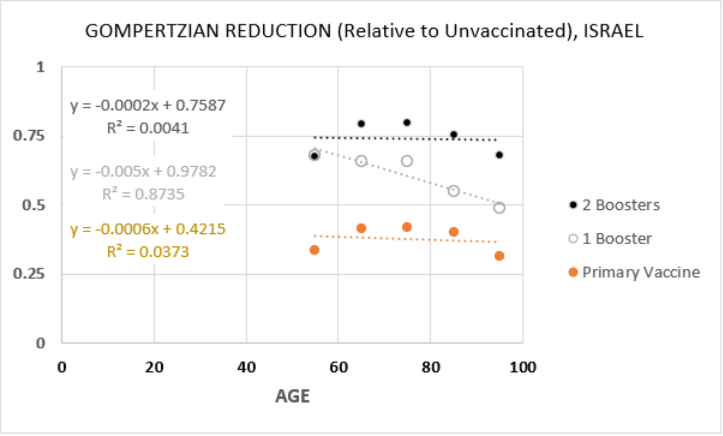

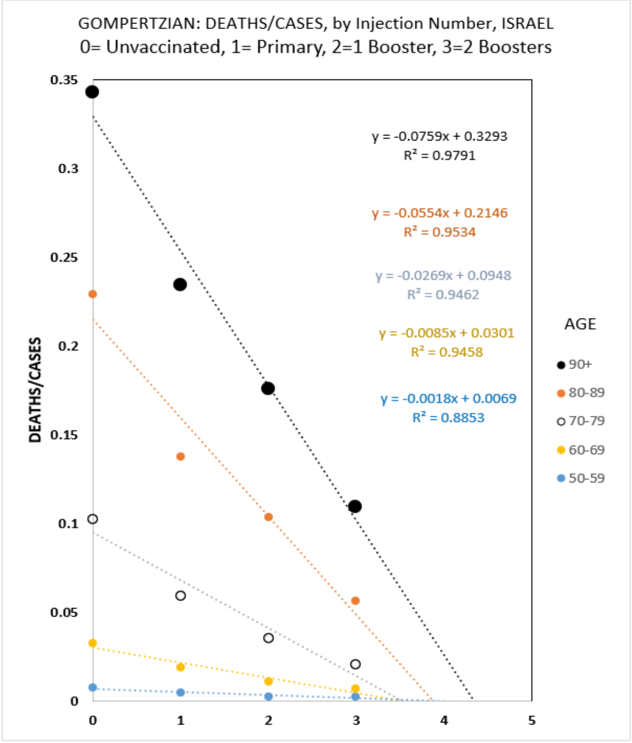

APPENDIX-FIGURE 24 APPENDIX-FIGURE 25

A second study from Israel data captured the number of **COVID-19** Deaths, per populations, in the Pre-vaccination era (March 23 2020-28 March 2021),[41] while the third study examined **COVID-19** deaths, per population, among people over age 60 who had had a primary vaccination with one booster (3 doses) or two boosters (4 doses) (APPENDIX-FIGURE 26).[42] Combing these on ***Gompertz Plots*** of ***Malthusian Lethality***, reveals that an unvaccinated 80-year-old at the beginning on the epidemic would have had a 0.34% chance of death, declining to 0.16% after vaccination and 1 booster, and declining yet further to 0.014% after vaccination and two boosters, a twelve-fold reduction in the chance of death (APPENDIX-FIGURE 26)

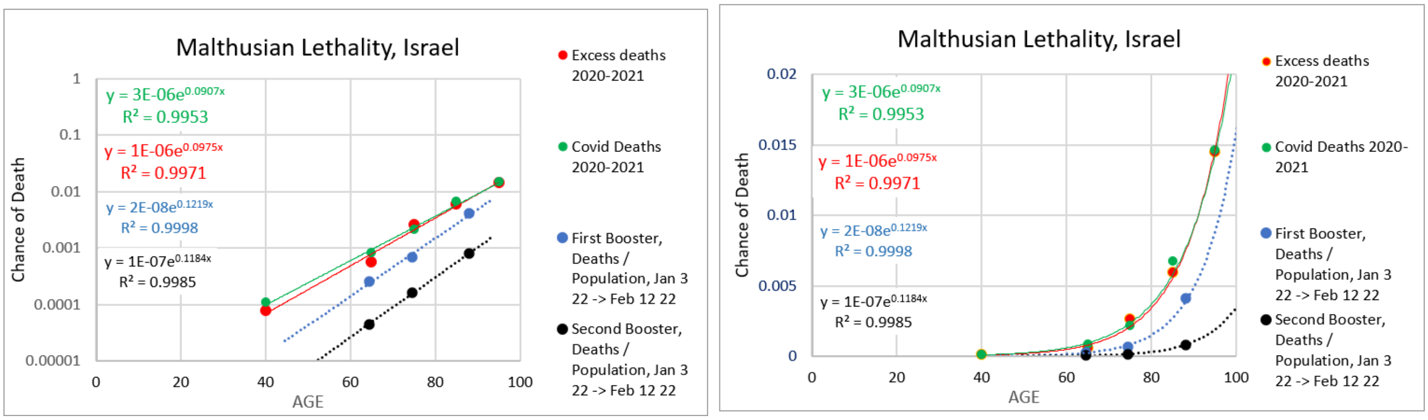

APPENDIX-FIGURE 26

Yet another study from Israel, provided data which revealed lower death rates for the those with 2 boosters (4 doses), than those with only 1 Booster (3 doses).[43] There were 35 deaths in among the 9,021 cases among those who had had 3 doses (1st booster) (0.38%), and 9 deaths among the 5,040 cases among those who had 4 doses (2 boosters) (0.18%). These deaths occurred among three age groups, and using the risk of death values that would have been expected for each age group, as calculated from the USA CDC data, for patients with a 4th dose (2 boosters), yielded an expect 0.09% risk of death, resulting in an expected of 5 deaths, which is in the same general area as the 9 deaths that actually occurred in this study group. Furthermore, patients who had had a previous COVID infections were excluded from analysis, so this study reveals the impact of sequential vaccination without the confounding immunizing effect of infection.

Finally, yet another study from Israel collected data on severe **COVID-19**. The data from this study also revealed a better outcome for those with 4 doses (2 boosters) than those who had only 3 doses (1 booster) (APPENDIX-FIGURE 27).[44]

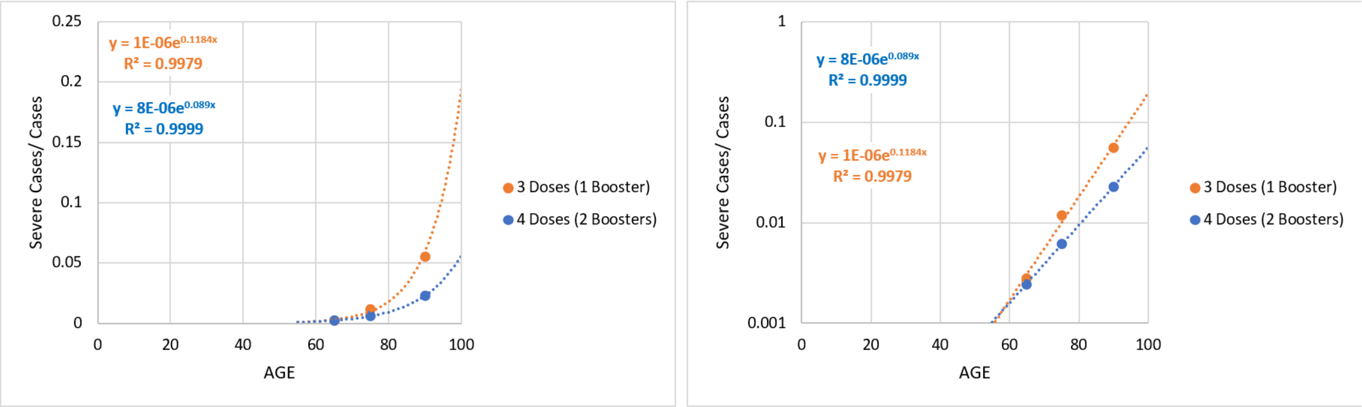

APPENDIX-FIGURE 27

Each ***Vaccination Event*** decreased **COVID-19 *Lethality*** (Deaths/Cases) by about 1/3rd.

Returning to the data introduced in APPENDIX FIGURE 23-25, in APPENDIX FIGURE 28 (FIGURE 2 in text) are shown the values for **COVID-19 *Lethality*** (Deaths/Cases), by age, for groups of Israeli patients: "unvaccinated"; "Only Primary Vaccination"; or "Primary Vaccination +1 Booster"; or "Primary Vaccination +2 Boosters".[45] This dataset excludes patients which have had infections, which allows us to see the impact of only vaccination on **COVID-19 *Lethality*** (Deaths/Cases).

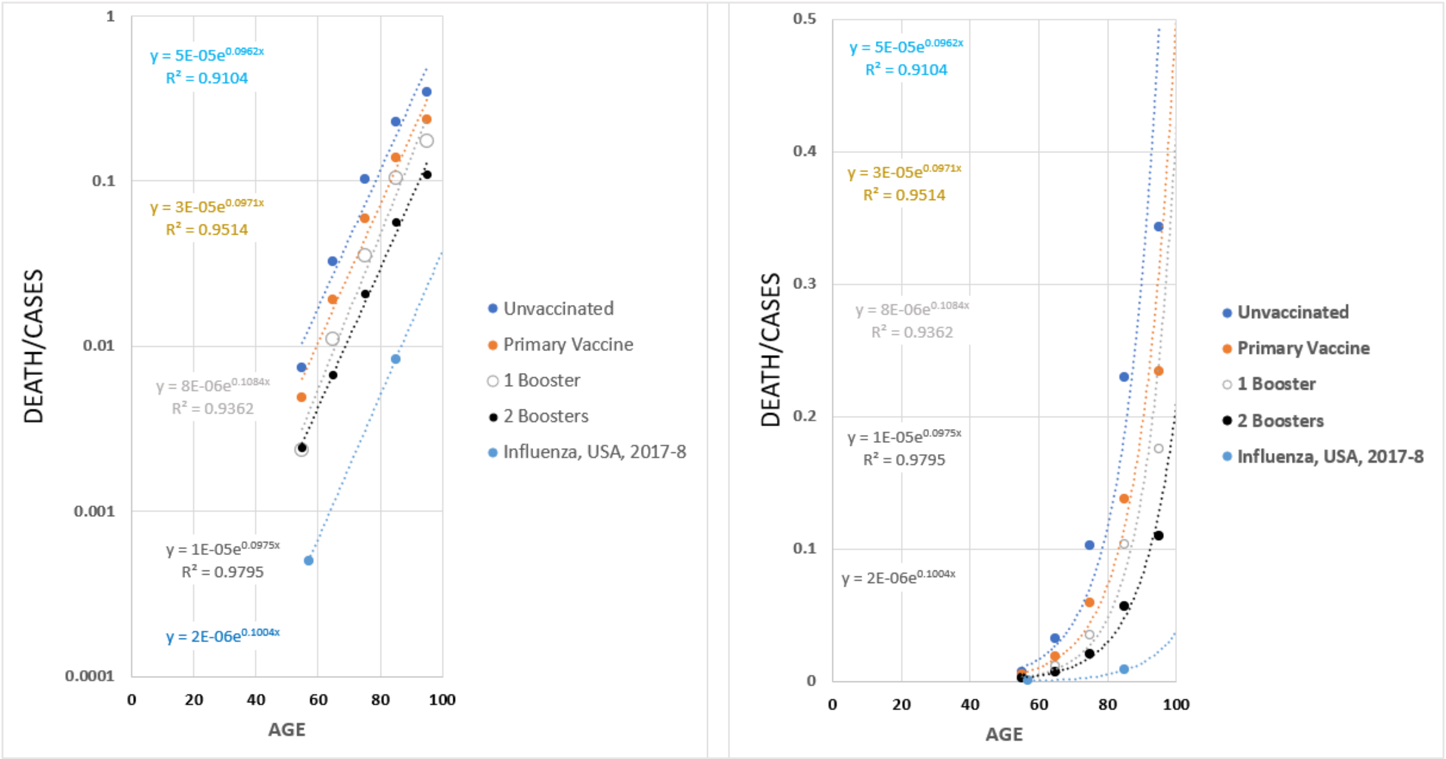

APPENDIX FIGURE 28 (**FIGURE 2**). **COVID-19 *Lethality*** (Deaths/Cases), by age, for groups of Israeli patients who have chosen one of four opportunities for vaccination: those who are "unvaccinated"; those who have had "Only Primary vaccination"; or "Primary vaccination +1 Booster"; or "Primary vaccination +2 Boosters", in comparison to the ***Gompertz Lines*** for groups of patients with influenza. This dataset excludes patients which have had infections, thus containing the impact of only vaccination on **COVID-19 *Lethality***.

For each ***Vaccination Event***, the ***Gompertz Line***, and thus ***Gompertzian Height***, ***$G_H$***, of **COVID-19 *Gompertzian Lethality*** (Deaths/Cases), declined cumulatively. Patients who had had only primary vaccination received a 38% reduction in **COVID-19 *Lethality*** (Deaths/Cases), captured by ***$G_H$***, relative to the unvaccinated. Those with the first booster received an additional 38% reduction. Those with a second booster received an additional 32% reduction (FIGURE 2 (APPENDIX FIGURE 29), TABLE I).

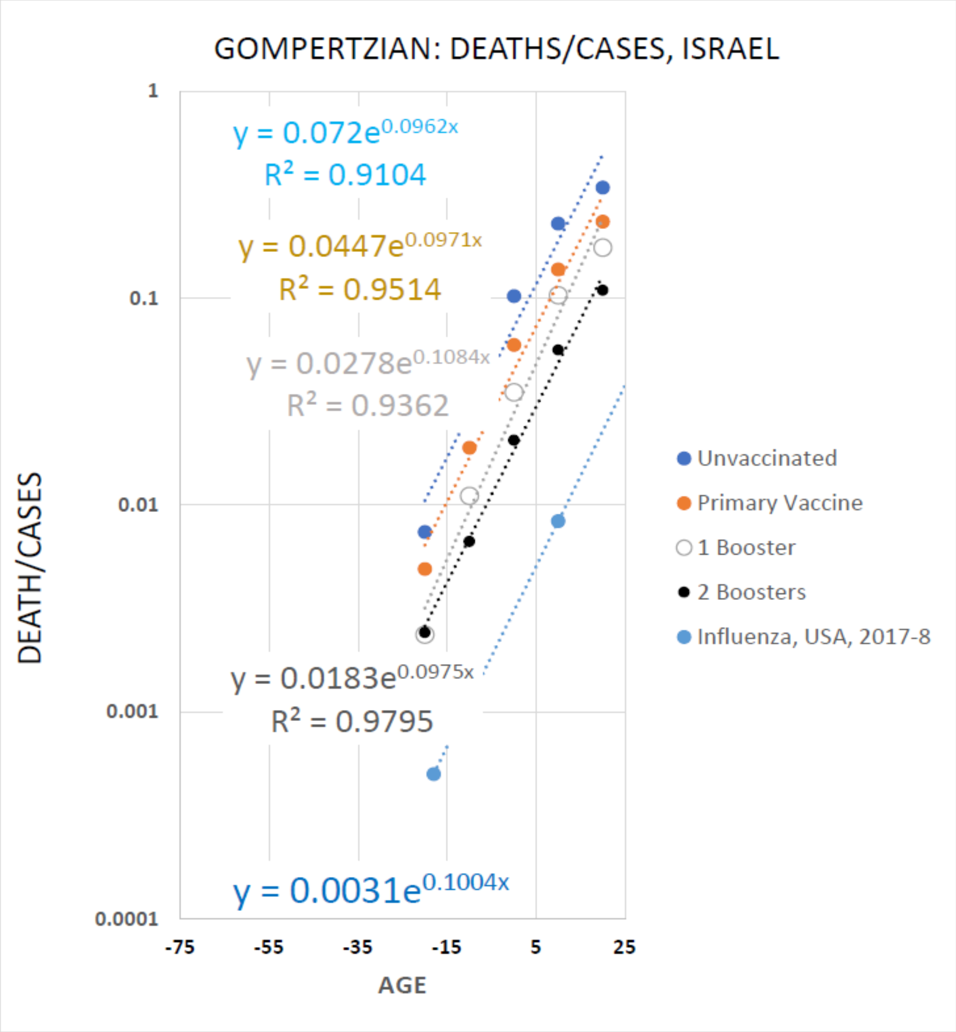

APPENDIX-FIGURE 29

| Event | Reduction in ***Gompertzian Height***, ***$G_H$***, of **COVID-19 *Lethality*** |
|---|---|
| Primary Vaccination | 38% |
| 1st Booster | 38% |
| 2nd Booster | 32% |

TABLE I

The vertical axis is formatted to where the ***Gompertz Mortality Equation*** intersects it at age 75.

We can examine the statistical strength of these measures of various numbers of ***Vaccination Events*** by applying the method described above, with which we examined the Hungary data in the APPENDIX-TABLES I and II above. Furthermore, this approach allows us to make such calculations without using the extreme age groups -age 10 and younger and age 90 and older -, whose precise age distributions are not known. By doing such a calculation, it was seen that the 95% confidence interval for the unvaccinated was 0.2%. Patients who had had only primary vaccination received a 36% reduction in **COVID-19 *Lethality*** (Deaths/Cases), captured by ***$G_H$***, relative to the unvaccinated (+/- 0.4%). Those with the first booster received an aggregate 59% reduction, relative to the unvaccinated (+/- 0.2%). Those with a second booster received an aggregate 75% reduction (+/- 0.6%).

Below, we shall also examine the impact of 1, 2, and 3 ***Vaccination Events*** in the USA. As we shall see, comparison of the impact of 1, 2, and 3 ***Vaccination Events*** in the USA and Israel reveals that the ***Gompertzian Reductions*** for each off the 3 ***Vaccination Events*** were remarkably similar in the two countries. (See "IV. Comparison of USA and Israel Booster Studies" below)

Curiously, the reduction in **COVID-19 *Gompertzian Lethality*** (Deaths/Cases) caused by vaccination can be seen, as noted above, by a reduction in the ***Gompertzian Height***, ***$G_H$***, of **COVID-19 *Gompertzian Lethality*** (Deaths/Cases), or, vaccination can be viewed as reducing in the operational age of the patient, by a reduction in **COVID-19 *Gompertzian Lethality*** age of about 4 years for each ***Vaccination Event***. Vaccination appears to make us functionally younger.

2 more boosters could push **COVID-19 *Lethality*** down to the familiar level of **Influenza *Lethality***.

What can be expected from additional ***Vaccination Events***, presuming they would again lead to additional ~1/3rd reductions in ***Gompertzian Height***, ***$G_H$***, of **COVID-19 *Lethality*** (Deaths/Cases)? Five ***Vaccination Events*** (Primary, 1st, 2nd, and 3rd Booster, and vaccination in Fall of 2023) have already been approved by the FDA and other organizations. We have already seen that primary plus 2 boosters has led to a 5-fold reduction **COVID-19 *Lethality*** (Deaths/Cases) (APPENDIX-FIGURE 29, TABLE 2). Below, we shall see some partial data that suggest that there was further reduction in **COVID-19 *Gompertzian Lethality*** by the third booster in the USA (APPENDIX-FIGURE-40 and APPENDIX-FIGURE-41 in section: "5. Boosters, October 2021 to Dec 2022: 3rd Booster (***Bivalent***) decreases **COVID-19 *Lethality*** further."). By considering two more ***Vaccination Events***, these calculations point to the possibility of a 25-fold reduction in **COVID-19 *Lethality*** (Deaths/Cases) (APPENDIX-FIGURES 30 and 31 and main text FIGURES 3 and 4). These values in tabular form can be seen in TABLE 2, below.

The ***Gompertz Mortality Equation*** for influenza is also shown on APPENDIX-FIGURE 30 (main text FIGURE 3) and lies essentially where the calculated **COVID-19 *Lethality*** (Deaths/Cases) would roughly be after these additional ***Vaccination Events***. Thus, approval of just two more ***Vaccination Events*** would appear to offer the chance of bringing the lethal burden of **COVID-19** down to the familiar level we see for influenza (APPENDIX-FIGURES 30 and 31 and main text FIGURES 3 and 4).

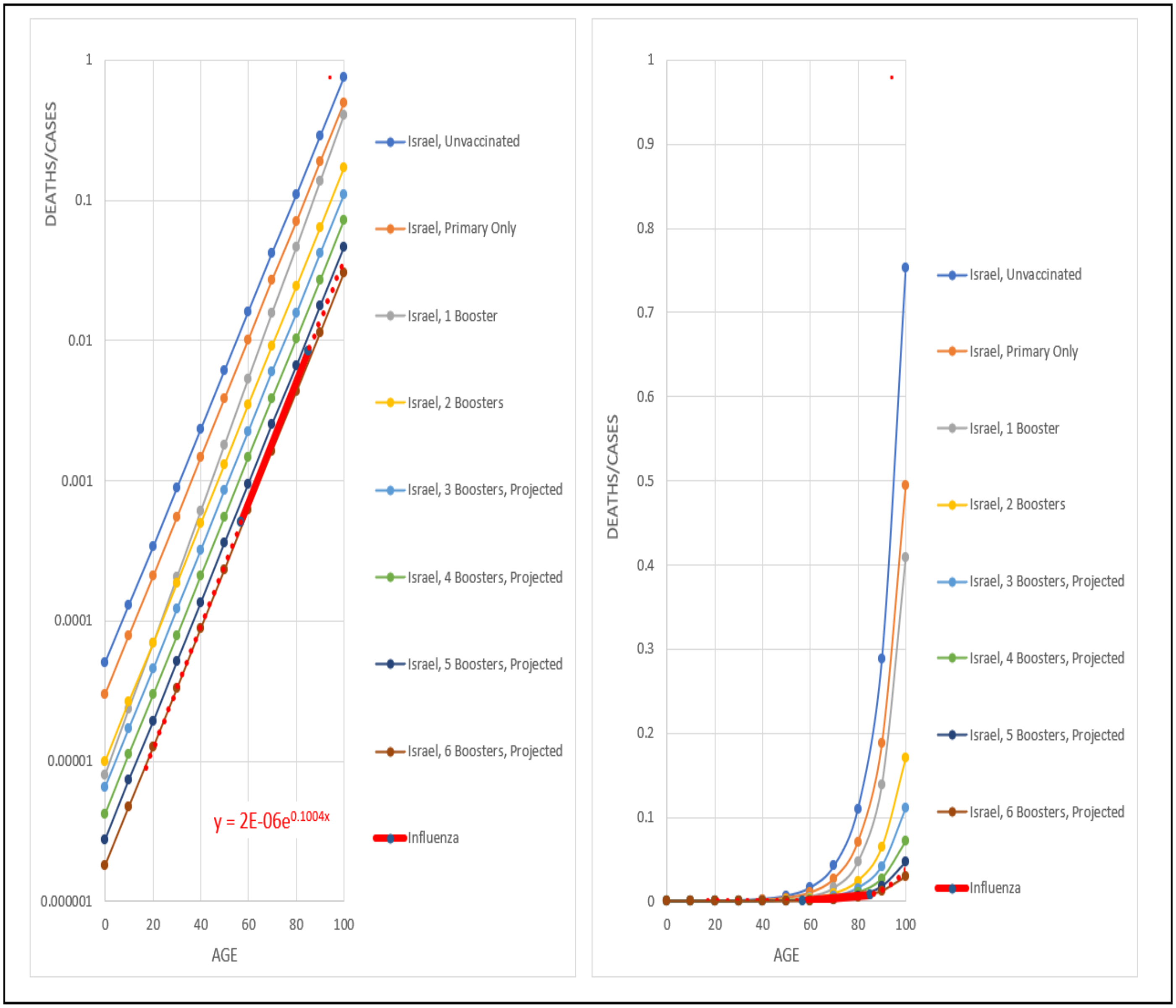

FIGURE 3 (APPENDIX-FIGURE 30)

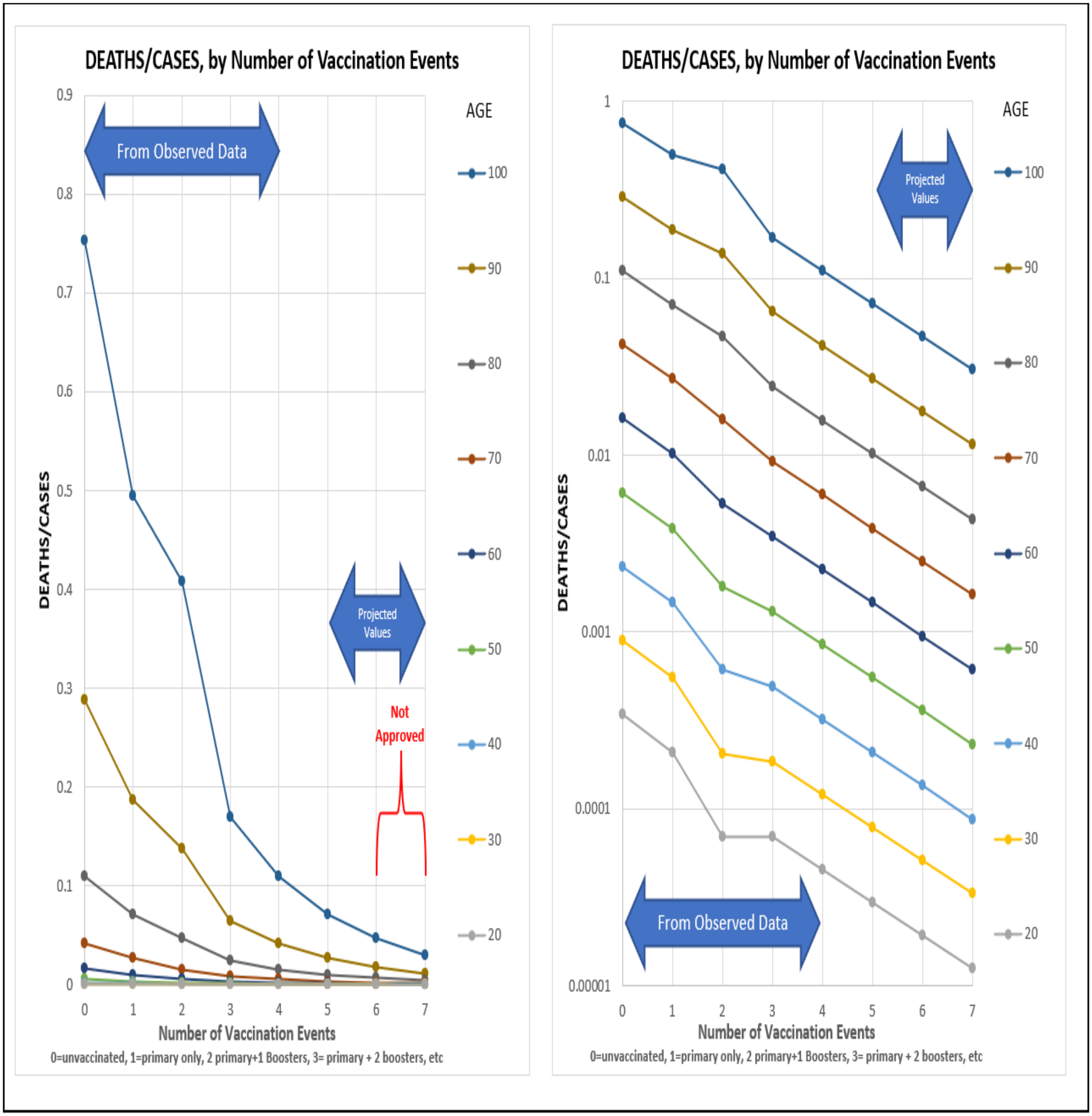

APPENDIX-FIGURE 31(FIGURE 4)

#### II. USA(CDC): March 2022 to July 2022, and October 2021 to Dec 2022, Boosters.

The second CDC dataset, from March 2022 through July 2022, thus one month beyond the first dataset described above, contains additional information on booster status[46] (APPENDIX-FIGURE 32). The first boosters were approved in late September 2021, the second boosters at the end of March 2022, and the third boosters in September 2022. The CDC dataset of roughly 65 million Americans shows that as of June 2022, about 10 million had been vaccinated to 2 boosters, ~20 million had been vaccinated to 1 booster, ~57 million vaccinated but no boosters yet, and ~10 million were unvaccinated. Thus, about 87% of the population had any vaccination, 39% had one booster, 17% had two boosters, 31% had vaccination but no boosters, and 13% had no vaccination. This dataset has two age groups (50-64 and 65+).

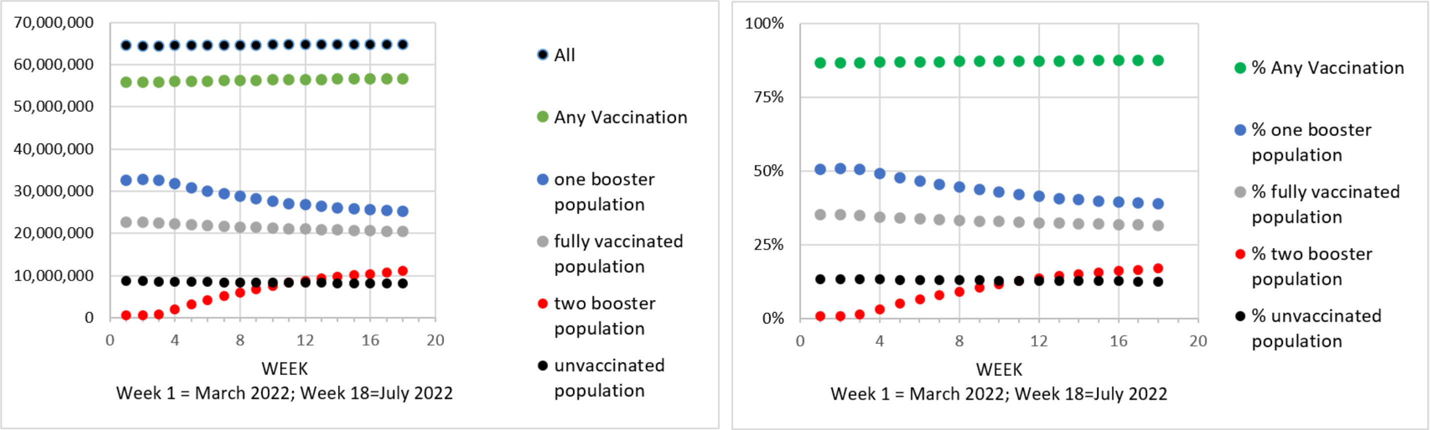

APPENDIX-FIGURE 32

#### 1. Boosters: March 2022 to July 2022: ***Gompertzian Lethality*** (Deaths/Cases)

The graphs in APPENDIX-FIGURE 33 below show ***Gompertzian Lethality*** (Deaths/Cases), from the CDC dataset of March 2022 to July 2022.[46] Having only two age groups (50-64 and 65+) makes it impossible to examine fit to the ***Gompertz Mortality Equation***. However, the two data points can be placed on a ***Gompertz Plots***, displaying a reduction in the ***Gompertzian Height***, **$G_H$**, in the presence of a roughly constant ***Gompertzian Slope***, **$G_S$**, as we have seen in so many examples above.

These values show that the chance of death after infection, ***Gompertzian Lethality*** (Deaths/Cases), was highest for those who are unvaccinated, lower for those vaccinated without boosters, even lower for those vaccinated, followed by 1 booster, and lowest yet for those with 2 boosters. The magnitude of these beneficial effects of boosters for each of these two age groups (50-64 and 80+) are shown in APPENDIX-FIGURE 33.

- For those vaccinated without boosters, the ***Gompertzian Reduction*** was 77%, a 1.3-fold reduction in (Deaths/Cases) in comparison to the unvaccinated.
- For those vaccinated followed by 1 booster, the additional ***Gompertzian Reduction*** was 41%, an accumulated 2.5-fold reduction in (Deaths/Cases) in comparison to the unvaccinated.
- For those vaccinated followed by 2 boosters, the additional ***Gompertzian Reduction*** was 32%, an accumulated 3-fold reduction in (Deaths/Cases) in comparison to the unvaccinated.

However, because the data includes an unknown number of individuals who have been immunized by infection, and is available in only two age groups, these values are likely to be less accurate that the Israeli data not above. Nonetheless, comparison of the impact of 1, 2, and 3 ***Vaccination Events*** in the USA and Israel reveals that the ***Gompertzian Reductions*** for each off the 3 ***Vaccination Events*** were remarkably similar in the two countries. (See "IV. Comparison of USA and Israel Booster Studies" below)

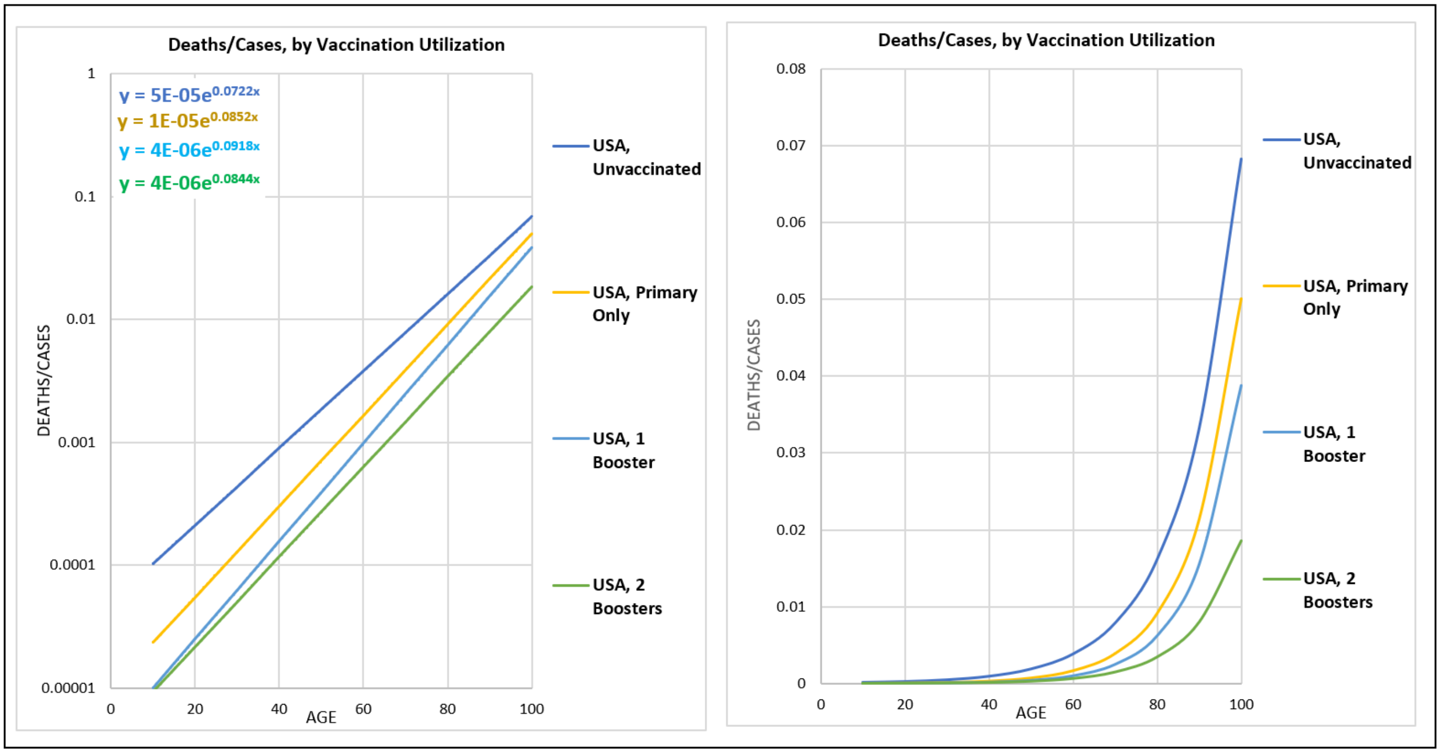

APPENDIX-FIGURE 33

The ***Gompertz Mortality Equations*** for the relationship between **COVID-19** Lethality, ***D***, the risk of death if infected, in units of (deaths/million cases), vs age, ***t***, for groups of USA patients with various numbers of ***Vaccination Events***. [46]

2. Boosters: March 2022 to July 2022: Persistence of ***Gompertzian Lethality*** (Deaths/Cases)

The CDC dataset provided the data in weekly increments over the 18-week period from March 2022 to July 2022 (APPENDIX-FIGURES 34-36). The “1” booster group is perhaps most informative, as its numbers decline over time, no doubt because some of them become “2” boosters (APPENDIX-FIGURE 34). Thus, we suspect that few individuals are added to the “1” booster group as the study proceeded. The data show that the level of **COVID-19 *Lethality*** (Deaths/Cases) for both age groups remained remarkably constant over the 18-week period.

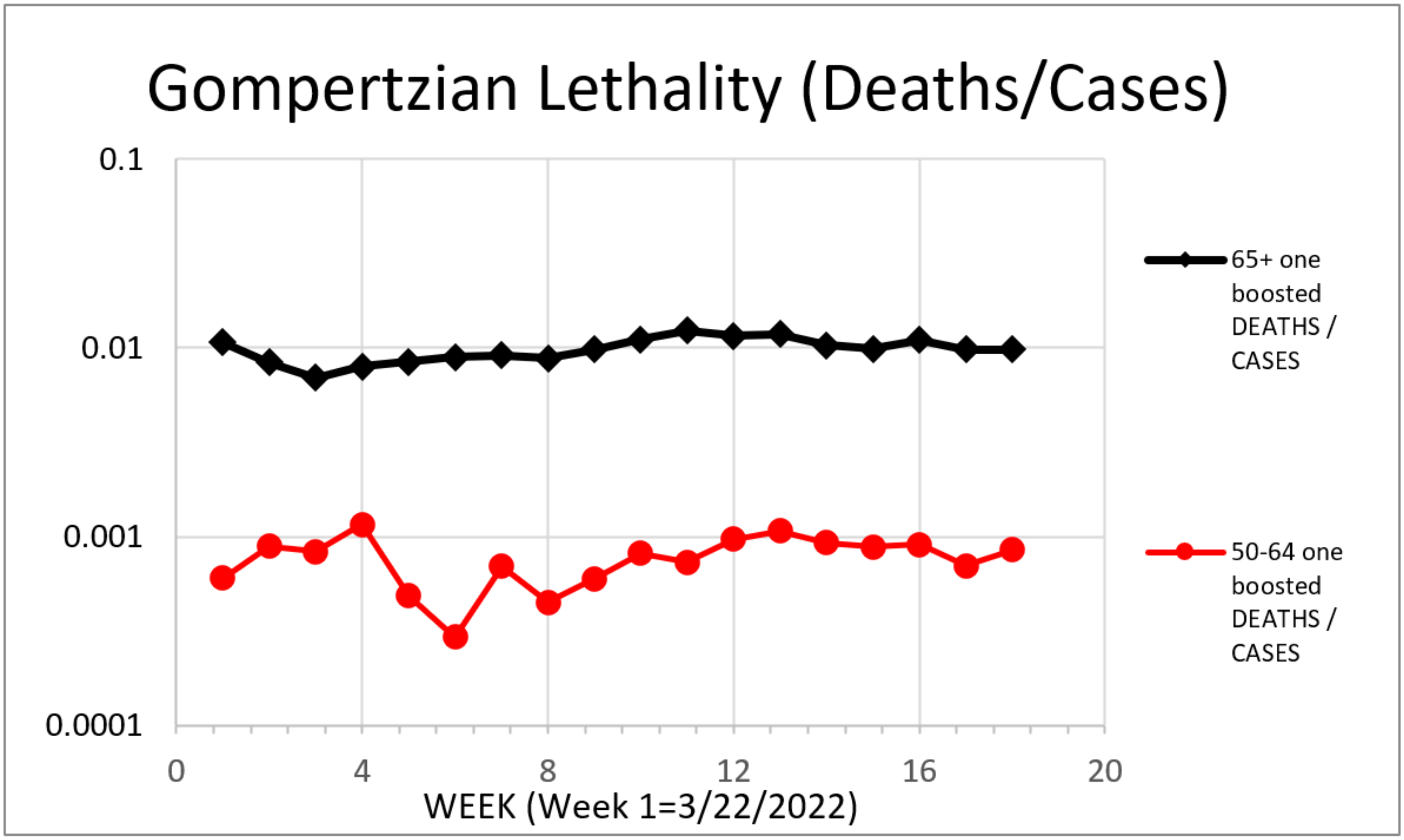

APPENDIX-FIGURE 34

3. Boosters: March 2022 to July 2022: Lack of Persistence of ***Pasteurian Infectivity*** (Cases/Population)

The graphs in APPENDIX-FIGURE 35 below show ***Pasteurian Infectivity*** (Cases/Population) for each of the 18 weeks, covered between March 2022 and July 2022. These values reveal that the chance of infection is highest for those who are unvaccinated and lower for those vaccinated without boosters. Boosters did not appear to have added any obvious additional benefit, and, in fact, those with 1 or 2 boosters showed slightly poorer reduction in ***Pasteurian Infectivity*** than those that have simply been vaccinated, although the differences are so small that they may be irrelevant.

The magnitude of these beneficial effects of vaccination and boosters are measured by their ***Pasteurian Reductions*** (1-[vaccinated/unvaccinated]; Cases/Population), shown in (APPENDIX-FIGURE 35), averaged for the two age groups (50-64 and 80+):

For those vaccinated without boosters, the ***Pasteurian Reduction*** was 73%, a 3.6-fold reduction in (Cases/Population), in comparison to the unvaccinated.

For those vaccinated followed by 1 booster ***Pasteurian Reduction*** was 53%, a 2.1-fold reduction in (Cases/Population), in comparison to the unvaccinated.

For those vaccinated followed by 2 boosters ***Pasteurian Reduction*** was 63%, a 2.7-fold reduction in (Cases/Population), in comparison to the unvaccinated.

See below for more visualizations, in APPENDIX-FIGURES 36-39.

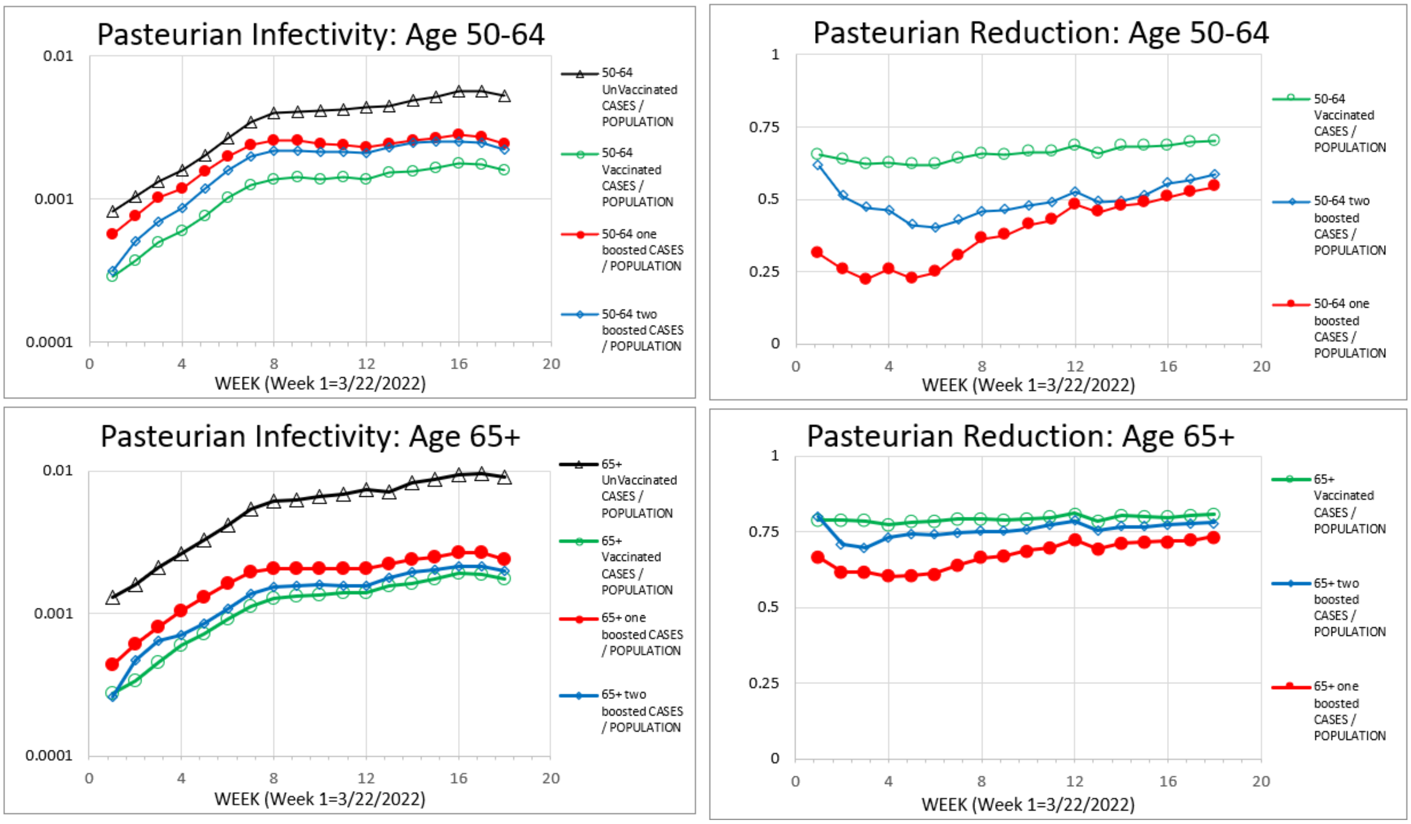

APPENDIX-FIGURE 35

4. Boosters: ***Gompertzian Lethality***, ***Pasteurian Infectivity***, ***Malthusian Lethality*** compared

In APPENDIX-FIGURES 36-38, the ***Gompertzian Lethality*** (Deaths/Cases), ***Pasteurian Infectivity*** (Cases/Population), and ***Malthusian Lethality*** (Deaths/Population), from the USA CDC dataset averaged over all 18 weeks of March 2022 to July 2022, are displayed by the number of ***Vaccination Events***. On these graphs, "0" identifies no vaccination, "1" identifies vaccination without boosters, "2" identifies vaccination followed by 1 booster, "3" identifies vaccination followed by 2 boosters:

For ***Gompertzian Lethality*** (Deaths/Cases, APPENDIX-FIGURE 36), as noted above, the chance of death if infected, relative to the unvaccinated, was reduced by each ***Vaccination Event*** (Primary & Boosters), cumulatively.

For ***Pasteurian Infectivity*** (Cases/Population, APPENDIX-FIGURE 37), the chance of infection, relative to the unvaccinated, was reduced by the initial primary vaccination, but this initial benefit became less after the first booster, and even less after the second booster.

Whether these changes to ***Pasteurian Infectivity*** were due to weakening action of vaccines among the vaccinated, changed variants, or improved resistance among the unvaccinated due to infection, is not known from these data, although resolving this would be doable, and of considerable value.

For ***Malthusian Lethality*** (Deaths/Population, APPENDIX-FIGURE 38), the combined benefits of ***Gompertzian*** and ***Pasteurian Reduction***, despite the disappointing impact of sequential boosters on ***Pasteurian Infectivity*** (Cases/Population), the favorable reductions in ***Gompertzian Lethality*** (Deaths/Cases), added up together to an overall favorable population-wide reduction of ***Malthusian Lethality*** with each ***Vaccination Event***, leading to progressively greater reductions in population-wide rates of death.

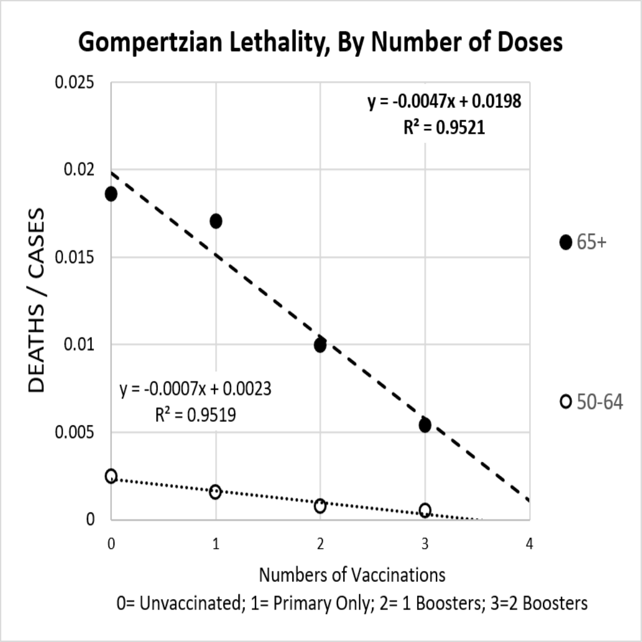

APPENDIX-FIGURE 36

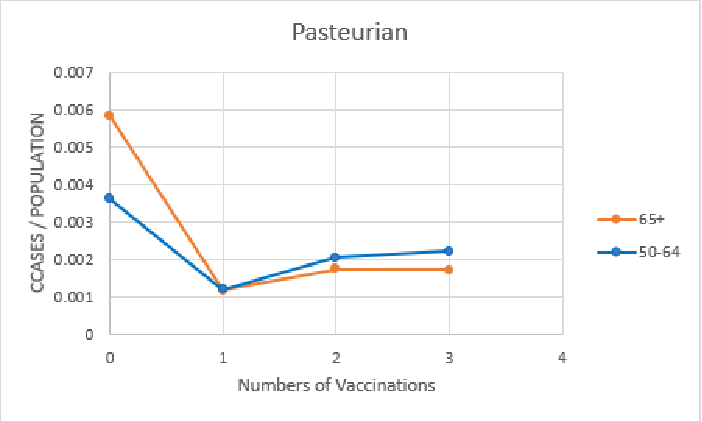

APPENDIX-FIGURE 37

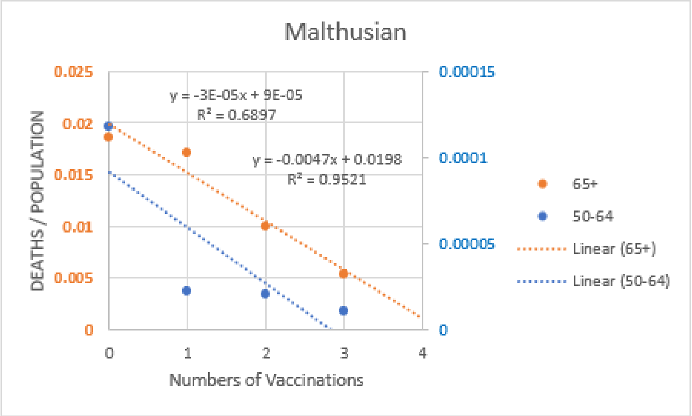

APPENDIX-FIGURE 38

5. Boosters, October 2021 to Dec 2022: 3rd Booster (***Bivalent***) decreases **COVID-19 *Lethality*** further.

In February 2023, the CDC published a study on the on the impact of the primary vaccination, monovalent, and bivalent boosters[47]. Unfortunately, the CDC has yet to release data broken down by number of boosters, but considerable insight can still be gained.

Most strikingly, as can be seen in APPENDIX-FIGURE-40 and APPENDIX-FIGURE-41, the ***Gompertz Line*** for **COVID-19 (*Gompertzian*) *Lethality*** (Deaths/Cases) for patients administered the ***Bivalent*** booster was indistinguishable from the ***Gompertz Line*** for the **COVID-19 (*Gompertzian*) *Lethality*** (Deaths/Cases) noted above for patients who had received 2 monovalent boosters (APPENDIX-FIGURE 33). This could only have been the case if those who had had 3 boosters, the last of which had the ***Bivalent*** formulation, had had a lower ***Gompertz Line***, that is, lower **COVID-19 (*Gompertzian*) *Lethality***, than those who had had 2 boosters. To picture why this is the case is a bit complicated, so bear with me.

Note that because these patients were not sorted by the number of booster shots that they had had, the group of patients who received the ***Bivalent*** booster included patients who had had 2 boosters, and patients who had had 1 booster, and patients who had not had any booster. Thus, before this mix of patients go their ***Bivalent*** booster, they had to have had a lower level of **COVID-19 (*Gompertzian*) *Lethality*** (Deaths/Cases) (that is, a lower ***Gompertz Line***, with a lower ***Gompertzian Height***, $G_H$) than the group of patients shown in APPENDIX-FIGURE 33, all of whom had had 2 boosters. If the ***Bivalent*** booster added no additional reduction in **COVID-19 (*Gompertzian*) *Lethality***, their ***Gompertz Line*** would have been higher than the ***Gompertz Line*** of the group of patients shown in APPENDIX-FIGURE 33, all of whom had had 2 boosters. However, those two ***Gompertz Lines*** were indistinguishable, indicating that the ***Bivalent*** booster did, indeed, push down the **COVID-19 (*Gompertzian*) *Lethality*** for patients.

Of course, precisely how much more the 3rd booster reduced the ***Gompertzian Height***, $G_H$, of **COVID-19 (*Gompertzian*) *Lethality*** beyond that achieved by the 2nd booster awaits the release of data on patients sorted by the number of boosters, as was made available for the March 2022 to July 2022 dataset of patients with 1 booster and 2 boosters (APPENDIX-FIGURE 33). Indeed, we await this valuable information with much appreciation and enthusiasm. But, for now, the data that has been made available so far are most definitely consistent with the occurrence of the ***Bivalent*** booster decreasing **COVID-19 *Lethality***, just as the 1st and 2nd boosters had decreased **COVID-19 *Lethality***.

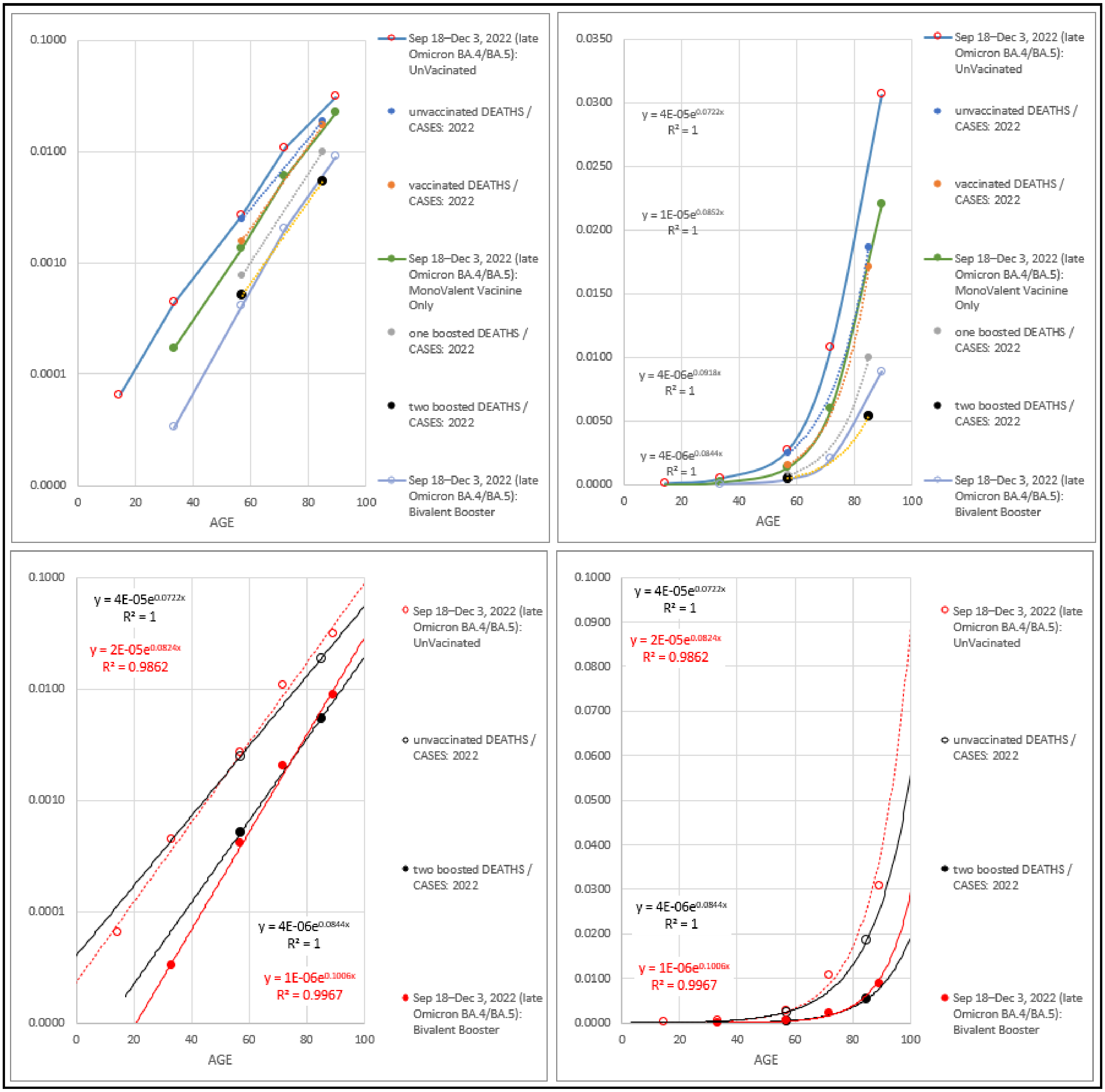

APPENDIX-FIGURE-40

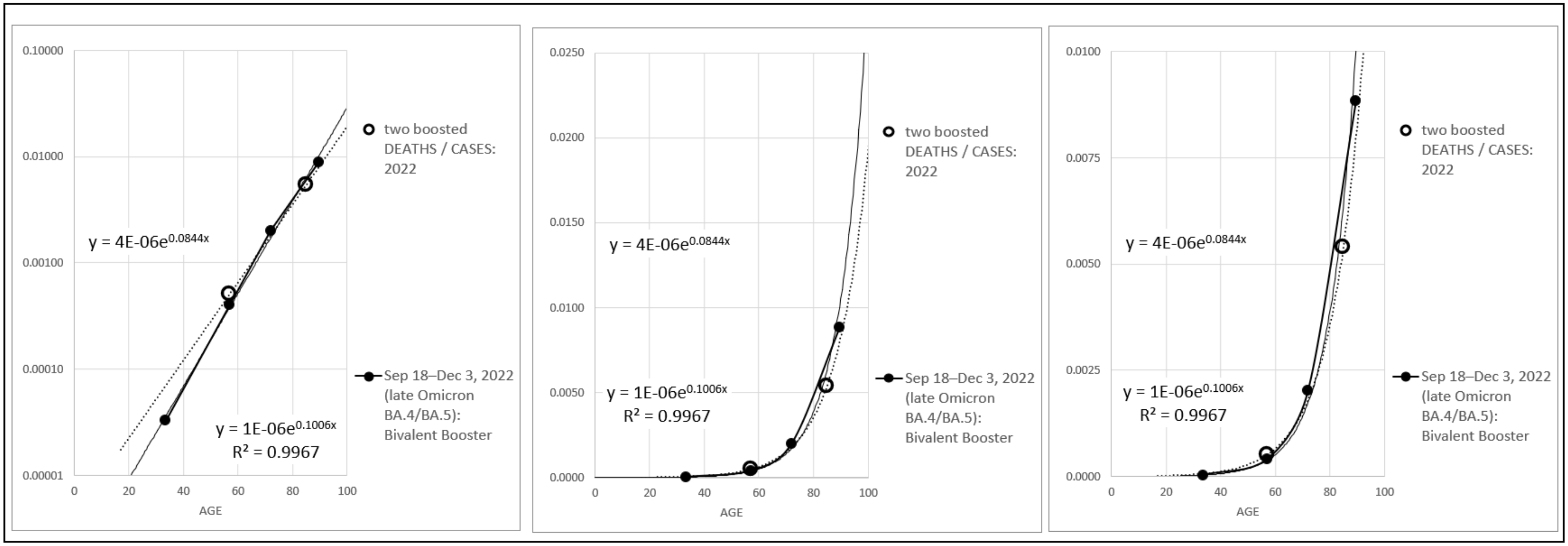

APPENDIX-FIGURE-41

**Sept 1, 2022**

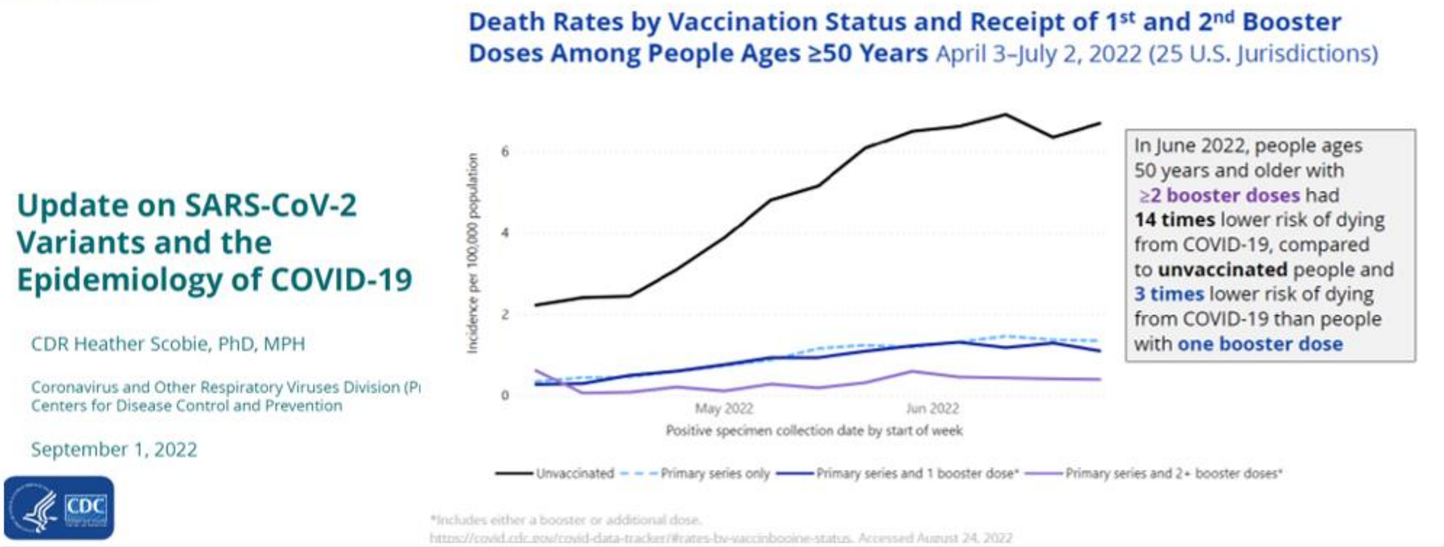

**March 6, 2023**

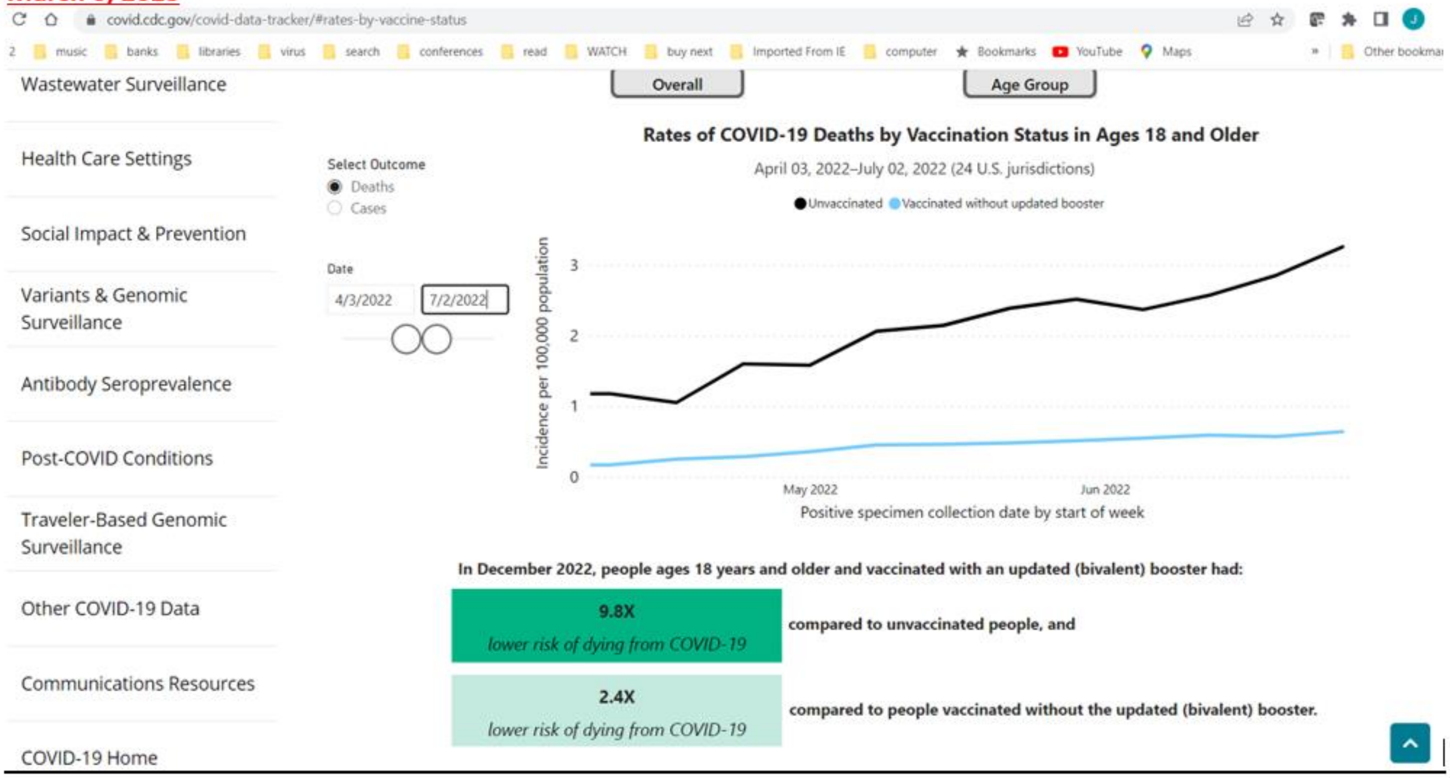

APPENDIX-FIGURE-42[48]

6. Boosters: The value of sorting patients by number of ***Vaccination Events***, infections, and age.

Fortunately, it's clear that our CDC colleagues, who have heroically created and analyzed this national treasure of a dataset, are aware of the information that can be gained from information on patients sorted by number of ***Vaccination Events***, infections, and age. We await this valuable data, for the potential that it has for comprehending, and minimizing, the lethal burden of **COVID-19**.

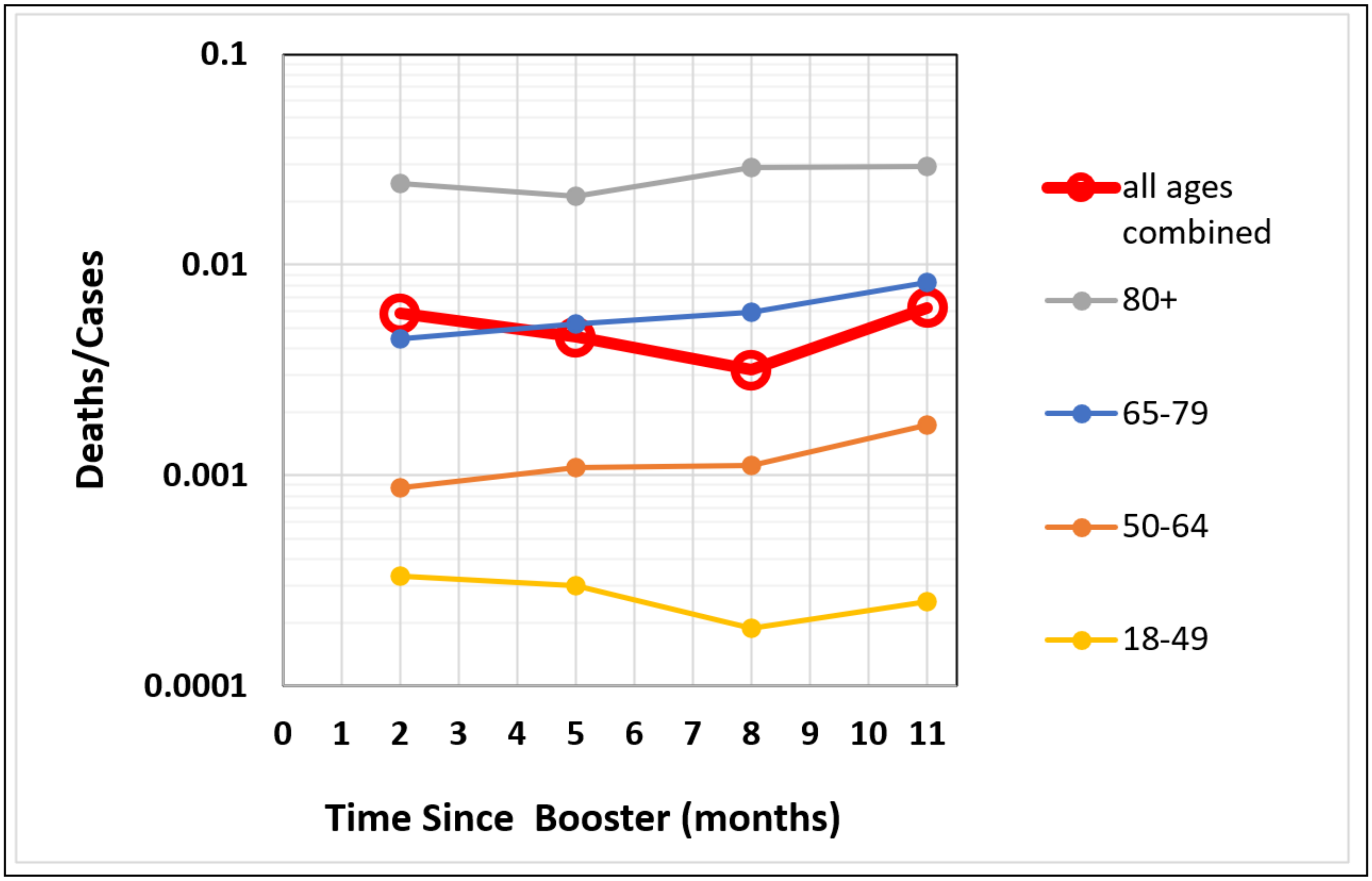

APPENDIX-FIGURE-43. (FIGURE 3).

**COVID-19 (*Gompertzian*) *Lethality*** (Deaths/Cases), for each age group, by time from last injection till infection.[47]

<u>7. Boosters: July 2021-June 2022: **COVID-19 *Lethality*** shows no waning for at least 11 months.</u>

The February 2023 CDC study[47] also observed that waning of **COVID-19 (*Pasteurian*) *Infectivity*** (Cases/Population) occurred in the months following injection. In addition, this study provided values, with which it was possible for us to calculate **COVID-19 (*Gompertzian*) *Lethality*** (Deaths/Cases), for each age group, by time from injection till infection (APPENDIX-FIGURE-43). These values, shown in APPENDIX-FIGURE-43, reveal that there was no evident waning of **COVID-19 (*Gompertzian*) *Lethality*** over the 11-months following booster administration. Booster administration in this dataset occurred from July 2021-June 2022, with **COVID-19 (*Gompertzian*) *Lethality*** values (Deaths/Cases) observed for patients who were infected in the March 20–June 25, 2022 (Omicron BA.2) period. Note, for example, that **COVID-19 *Lethality*** calculations for all ages combined, (Deaths/Cases) yielded a value at 2 months of 0.0059, and at 11 months of 0.63, a mere 6% difference. Yet again, the data provide us with a picture of vaccination's impact on **COVID-19 *Lethality*** (Deaths/Cases) being persistent, while vaccination's impact on **COVID-19 *Infectivity*** (cases/population) is fleeting.

#### III. HUNGARY, Boosters

The Hungary group followed the HUN-VE study,[34] analyzed above, with two more studies, HUN-VE 2,[35] and HUN-VE 3,[35] the first of which is of most relevance here, as it examined the impact of the first and second boosters that followed the primary vaccinations. Values of deaths and cases were provided for patients who had receive no vaccine (0 in APPENDIX-FIGURE-44), primary vaccination (1), primary vaccination plus just one booster (2), or two boosters (3) in the Omicron Wave, January 1, 2021- February 23, 2021. With their data on cases and deaths, sorted by age, we again could calculate the ***Gompertzian Reductions***: 56% (a 1.80-fold reduction in the lethality) after the primary vaccination ("1" on the x-axis, APPENDIX-FIGURE-44, Left); 92%, (an 11.81-fold reduction in the chance of death after infection, ("2" on the x-axis) after the First Booster; 98% (a 55.16-fold reduction in the chance of death after infection) after the Second Booster ("3" on the x-axis).

The practical implications of these reductions in **COVID-19 *Lethality*** can be seen by graphing the ***Gompertzian*** chance of death, relative to the unvaccinated (APPENDIX-FIGURE-44, Right). These numbers show the events from vaccination ("1" on the x-axis) to first booster ("2" on the x-axis) point to a very low risk of death, as realized by the second booster ("3" on the x-axis), which show progressively, additive, reductions in **COVID-19 (*Gompertzian*) *Lethality*** (Deaths/Cases), as we saw above for the ISRAEL and USA data.

These data again reveal that boosters elicit a cumulative, progressively greater, and lasting, resistance to **COVID-19 (*Gompertzian*) *Lethality*** with each sequential ***Vaccination Event*** (Primary & Boosters).

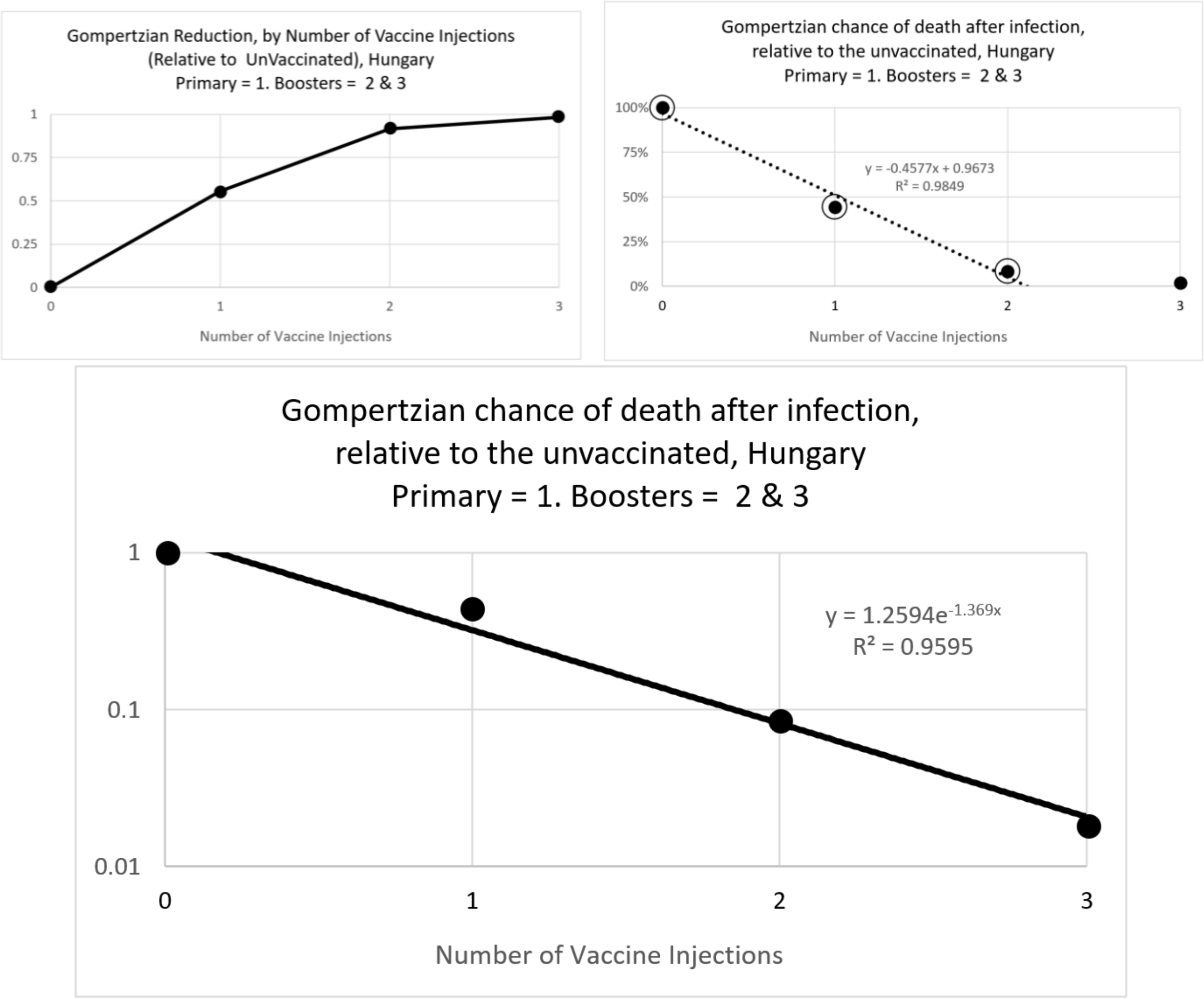

APPENDIX-FIGURE-44

| Gompertzian Reduction | | 1 Booster | 2 Boosters | Primary | BOTH |
|---|---|---|---|---|---|
| | Observed CFR/Expected CFR-> | 0.08468 | 0.01813 | 0.55573 | 0.08418 |
| | +/- 95% -> | 0.0014 | 0.0126 | 0.0123 | 0.0014 |
| | Reduction, as a % (1-v/un) | 92% | 98% | 56% | 92% |
| | **#-Fold Reduction in Death, once infected ->** | **11.81** | **55.16** | **1.80** | **11.88** |

These ***Gompertzian Reductions*** translated into massive saving in life, that becomes progressively more dramatic with age. As we can see on the ***Gompertz Plots***, and their linear visualizations, shown above, while an unvaccinated 80-year-old at the beginning on the epidemic would have had a 40% chance of death if infected, that number declined to 3% after vaccination and one booster, and 1% after vaccination and two boosters (APPENDIX-FIGURE 45).

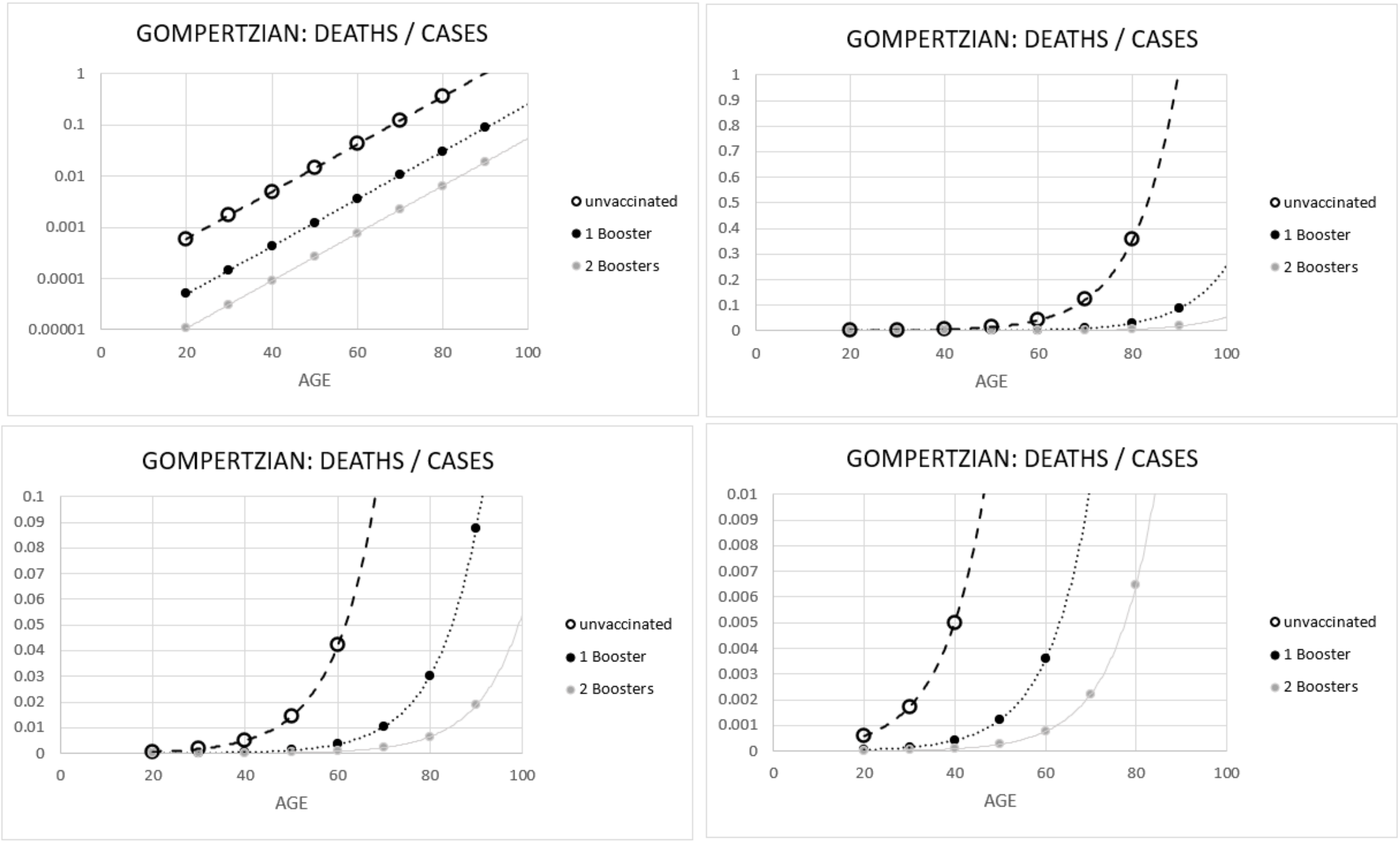

(APPENDIX-FIGURE-45)

Unlike the Israel data, individuals with infections were not excluded, so quantifying the impact of vaccination alone is problematic, but the encouragingly cumulative reduction in death with each ***Vaccination Event*** is certainly made clear. Hungary had an unusually high number of infections at the start of the pandemic, so it may be difficult to disentangle the impact of infection and vaccination. Nonetheless, the **COVID-19 *Gompertzian Height***, $G_H$, of **COVID-19 *Lethality*** (Deaths/Cases) in Hungary after 2 Boosters (APPENDIX-FIGURE-45) is quite close to the ***Gompertzian Height***, $G_H$, of **Influenza *Lethality*** (APPENDIX-FIGURE 30), and with an additional 1/3$^{rd}$ reduction that would be expected from a third booster, would reach that familiar level.

Regrettably, also, for the earlier phase, where primary only individuals, and unvaccinated individuals, were also studied, case numbers were not published. Thus, calculations for the impacts of the sequential vaccinations of primary vaccination followed by boosters cannot yet be calculated.

#### IV. Comparison of USA and Israel Booster Studies

\ Comparison of the impact of 1, 2, and 3 ***Vaccination Events*** in the USA and Israel reveals that the ***Gompertzian Reductions*** for each off the 3 ***Vaccination Events*** were remarkably similar in the two countries.

| Vaccination Events-> | 1 | 2 | 3 |
|---|---|---|---|
| **Gompertzian Reduction (USA)** | | | |
| +/- 95% -> | 0.0009 | 0.0009 | 0.0018 |
| Reduction, as a % (1-v/un)-> | 13% | 49% | 71% |
| **#-Fold Reduction in Death, once infected ->** | **1.15** | **1.96** | **3.50** |
| **Gompertzian Reduction (Israel)** | | | |
| +/- 95% -> | 0.0037 | 0.0024 | 0.0058 |
| Reduction, as a % (1-v/un)-> | 31% | 59% | 75% |
| **#-Fold Reduction in Death, once infected ->** | **1.45** | **2.44** | **3.93** |

#### V. More recent 2nd Booster Studies

A number of more recent studies have examined the relative rate of COVID death in those with 4 doses (2 boosters) vs those who had had 3 doses (1 booster), and while these studies have not provided fine-grained enough datapoints for ***Gompertzian Analysis***, all of these studies have shown the superior death-reducing potential of the additional dose. Two studies were of **COVID-19** deaths among patients in in old age homes,[49,50] another of death in a long-term care facility,[51] another of rates of admission to intensive care or death among elderly (isolated death values not provided),[52] and another of infection, hospitalization, or death in nursing home residents[53] All of these studies have found a superior reduction in death for 2 boosters (4 doses) over 1 booster (3 doses).

***Gompertzian Analysis*** provides useful values for the benefit, and marginal benefit, of vaccination, by age.

The values shown in APPENDIX-FIGURE 30 are also shown in TABLE 2 as Deaths/Cases, as percentages, by age, in decades, for patients who have availed themselves of the various numbers of ***Vaccination Events***. From these values, the number of deaths per million infections, are shown in TABLE 3. From these values, the number of deaths prevented, by all ***Vaccination Events***/million infections, were calculated (TABLE 4) by subtracting the number of deaths in each vaccination category in TABLE 3 from the number of deaths among the unvaccinated, on the first line in TABLE 3. In TABLE 5, are shown the number of deaths prevented, by last ***Vaccination Event***/million infections; these values are the ***Marginal Benefit*** of each ***Vaccination Event***. As we shall see next, such ***Marginal Benefit*** values provide a way to make age-related guidelines, which may be offered the public, to help in the rational choice of whether or not get a vaccination.

***Gompertzian Analysis*** gives us rational age-based vaccination guidelines.

At what ages would it be logical for each of us to avail ourselves of a ***Vaccination Opportunity***, and at what ages would it be logical for each of us to pass the opportunity by? The ~10,000-fold difference, from our teenage years to our life's end, in the number of deaths that ***Gompertzian Analysis*** shows us would be prevented by vaccination (APPENDIX-FIGURE 7, TABLES 4 and 5) makes clear that being able to decide such choices wisely would be exceedingly valuable.

The simple fact is that if a ***Vaccination Opportunity*** would give each of us the savings in life that we would expect in return for the trouble we take, or the price we pay, for that ***vaccination***, then we would want to get the shot. However, if a ***Vaccination Opportunity*** wouldn't give us the savings in life worth the trouble, or the expense, that we would be willing to pay out, then we would want to stay home. The values in TABLE 5, derived by ***Gompertzian Analysis*** from counting actual cases and deaths, give each of us the data we need for making such choices rationally.

Let us examine how individuals could use this information to decide on the four ***Vaccination Opportunities*** already approved (Primary and three boosters), and the ***contemplated*** 6$^{th}$, and 7$^{th}$ ***Vaccination Opportunities***, whose potential benefit we have characterized above. One basis by which individuals make such choices is the level of risk each of us is comfortable with. Let us examine a couple of vivid examples, which can make clear how these individual desires can be transformed into rational, individual decisions, for each ***Vaccination Opportunity*** at each age.

The ***Marginal Benefit*** values derived by ***Gompertzian Analysis*** (FIGURES 3&4, APPENDIX-FIGURES 30 &31, TABLE 5) make it possible to create rational, data-driven, age-specific vaccination guidelines, for reducing the risk of **COVID-19** death to the everyday risks we all accept. For example, most American accept the risk of driving (~120 automotive deaths/million-Americans/year). Should this risk level define our choices for vaccination, then primary vaccination and 1$^{st}$ booster would be wise if ~age 20 or older, the 2$^{nd}$ and 3$^{rd}$ boosters if ~age 40 or older, and the 4$^{th,}$ 5$^{th}$, and 6$^{th}$ boosters if ~age 50 or older (TABLE 5, yellow highlights).

On the other hand, one of the most dangerous professions, fishing, comes with ~1,200 work-related deaths/million fisherpersons. For those of us who would have such an acceptance of the higher level of risk that going to sea would give, who would get a shot only if it prevented at least 1,200 deaths/million infected Americans, then the values shown in blue highlights in TABLE 5 help us see which ***Vaccination Events***, at which ages, would be rational: primary immunization and the 1$^{st}$ booster as well if ~age 50 or older, then get the 2$^{nd}$ booster and 3$^{rd}$ booster if ~age 60 or older, then attend 4$^{th}$ and 5$^{th}$ boosters if ~age 70 or older, and then get the 6$^{th}$ booster if ~age 80 or older (TABLE 5, blue highlights)..

While these vivid examples illustrate how knowledge of ***Marginal Benefit*** values assembled by ***Gompertzian Analysis*** of cases and deaths can help us make rational choices, on a more fundamental, and mathematical level, it has long been appreciated that societies get the greatest benefit out of some action when everyone utilizes the resource to the same ***Marginal Benefit***, which are the values shown in TABLE 5. This principle, known as ***Pareto Optimality***, allows policy makers to design decision suggestions that that can give the greatest possible savings in life that match the resources available and the risk levels most of us accept. Other tools for generating the best possible guidelines, such as considering not lives saved, but years of life saved, or reduction in **COVID-19** ***Lethality*** as a fraction of all-cause lethality, may also be examined with these fundamental ***Marginal Benefit*** values. Practical considerations of vaccine distribution can also be taken into account.

| TABLE 2 Deaths/Cases: Percentages | | | | | | | | | | |
|---|---|---|---|---|---|---|---|---|---|---|
| Age-> | 0 | 10 | 20 | 30 | 40 | 50 | 60 | 70 | 80 | 90 | 100 |
| Israel, Unvaccinated | 0.0050% | 0.0131% | 0.0342% | 0.0896% | 0.2345% | 0.6137% | 1.6059% | 4.2025% | 10.9977% | 28.7801% | 75.3152% |
| Israel, Primary Only | 0.0030% | 0.0079% | 0.0209% | 0.0552% | 0.1459% | 0.3851% | 1.0170% | 2.6855% | 7.0912% | 18.7250% | 49.4448% |
| Israel, 1st Booster | 0.0008% | 0.0024% | 0.0070% | 0.0207% | 0.0611% | 0.1807% | 0.5342% | 1.5795% | 4.6697% | 13.8060% | 40.8171% |
| Israel, 2nd Booster | 0.0010% | 0.0026% | 0.0070% | 0.0186% | 0.0492% | 0.1303% | 0.3452% | 0.9142% | 2.4212% | 6.4125% | 16.9835% |
| 3rd Booster, Projected | 0.0007% | 0.0017% | 0.0046% | 0.0121% | 0.0320% | 0.0847% | 0.2244% | 0.5942% | 1.5738% | 4.1681% | 11.0393% |
| 4th Booster, Projected | 0.0004% | 0.0011% | 0.0030% | 0.0078% | 0.0208% | 0.0551% | 0.1458% | 0.3862% | 1.0229% | 2.7093% | 7.1755% |
| 5th Booster, Projected | 0.0003% | 0.0007% | 0.0019% | 0.0051% | 0.0135% | 0.0358% | 0.0948% | 0.2510% | 0.6649% | 1.7610% | 4.6641% |
| 6th Booster, Projected | 0.0002% | 0.0005% | 0.0013% | 0.0033% | 0.0088% | 0.0233% | 0.0616% | 0.1632% | 0.4322% | 1.1447% | 3.0317% |

| TABLE 3. Deaths/Million Infections | | | | | | | | | | | |
|---|---|---|---|---|---|---|---|---|---|---|---|
| Age-> | 0 | 10 | 20 | 30 | 40 | 50 | 60 | 70 | 80 | 90 | 100 |
| Israel, Unvaccinated | 50 | 131 | 342 | 896 | 2,345 | 6,137 | 16,059 | 42,025 | 109,977 | 287,801 | 753,152 |
| Israel, Primary Only | 30 | 79 | 209 | 552 | 1,459 | 3,851 | 10,170 | 26,855 | 70,912 | 187,250 | 494,448 |
| Israel, 1st Booster | 8 | 24 | 70 | 207 | 611 | 1,807 | 5,342 | 15,795 | 46,697 | 138,060 | 408,171 |
| Israel, 2nd Booster | 10 | 26 | 70 | 186 | 492 | 1,303 | 3,452 | 9,142 | 24,212 | 64,125 | 169,835 |
| 3rd Booster, Projected | 7 | 17 | 46 | 121 | 320 | 847 | 2,244 | 5,942 | 15,738 | 41,681 | 110,393 |
| 4th Booster, Projected | 4 | 11 | 30 | 78 | 208 | 551 | 1,458 | 3,862 | 10,229 | 27,093 | 71,755 |
| 5th Booster, Projected | 3 | 7 | 19 | 51 | 135 | 358 | 948 | 2,510 | 6,649 | 17,610 | 46,641 |
| 6th Booster, Projected | 2 | 5 | 13 | 33 | 88 | 233 | 616 | 1,632 | 4,322 | 11,447 | 30,317 |

| TABLE 4. Deaths Prevented, by all ***Vaccination Events***/Million Infections | | | | | | | | | | | |
|---|---|---|---|---|---|---|---|---|---|---|---|
| Age-> | 0 | 10 | 20 | 30 | 40 | 50 | 60 | 70 | 80 | 90 | 100 |
| Israel, Primary Only | 20 | 52 | 133 | 344 | 886 | 2,285 | 5,889 | 15,170 | 39,064 | 100,551 | 258,704 |
| Israel, 1st Booster | 42 | 107 | 272 | 689 | 1,734 | 4,330 | 10,717 | 26,230 | 63,279 | 149,741 | 344,981 |
| Israel, 2nd Booster | 40 | 104 | 272 | 710 | 1,853 | 4,833 | 12,607 | 32,884 | 85,765 | 223,676 | 583,317 |
| 3rd Booster, Projected | 44 | 114 | 297 | 775 | 2,025 | 5,289 | 13,815 | 36,083 | 94,239 | 246,119 | 642,759 |
| 4th Booster, Projected | 46 | 120 | 313 | 818 | 2,137 | 5,586 | 14,601 | 38,163 | 99,747 | 260,708 | 681,397 |
| 5th Booster, Projected | 47 | 124 | 323 | 845 | 2,210 | 5,779 | 15,111 | 39,515 | 103,328 | 270,190 | 706,511 |
| 6th Booster, Projected | 48 | 126 | 330 | 863 | 2,257 | 5,904 | 15,443 | 40,393 | 105,655 | 276,354 | 722,836 |

| TABLE 5. Deaths Prevented, by Last ***Vaccination Event***/Million Infections; Risk Levels Highlighted | | | | | | | | | | | |
|---|---|---|---|---|---|---|---|---|---|---|---|
| Age-> | 0 | 10 | 20 | 30 | 40 | 50 | 60 | 70 | 80 | 90 | 100 |
| Israel, Primary Only | 20 | 52 | 133 | 344 | 886 | 2,285 | 5,889 | 15,170 | 39,064 | 100,551 | 258,704 |
| Israel, 1st Booster | 22 | 56 | 139 | 346 | 847 | 2,044 | 4,828 | 11,060 | 24,215 | 49,190 | 86,277 |
| Israel, 2nd Booster | 0 | 0 | 0 | 21 | 119 | 504 | 1,891 | 6,653 | 22,486 | 73,935 | 238,336 |
| 3rd Booster, Projected | 4 | 9 | 25 | 65 | 172 | 456 | 1,208 | 3,200 | 8,474 | 22,444 | 59,442 |
| 4th Booster, Projected | 2 | 6 | 16 | 42 | 112 | 296 | 785 | 2,080 | 5,508 | 14,588 | 38,638 |
| 5th Booster, Projected | 1 | 4 | 10 | 27 | 73 | 193 | 510 | 1,352 | 3,580 | 9,482 | 25,114 |
| 6th Booster, Projected | 1 | 3 | 7 | 18 | 47 | 125 | 332 | 879 | 2,327 | 6,164 | 16,324 |

Who's dying now?

The data reviewed above make clear that people who have had 2 boosters have a better chance of surviving after infection than people with 1 booster, while people with 1 booster have a better chance than people with just primary vaccination, and people with primary vaccination have a better chance than the unvaccinated. The data also show that the vaccinated have a lower chance of getting infected than the unvaccinated.

So how does all this translate into who is dying now? The CDC vaccination and booster status data on people with **COVID-19** infections from March 2022 through July 2022 give us a picture of this matter (APPENDIX-TABLE VI).

- 38% of the deaths occurred among the unvaccinated, even though they made up only 12% of the population.
- Only 5% of the deaths occurred among those who have had two boosters, even though they made up 17% of the population.
- Those with only primary vaccination have lowered their chance of dying of **COVID-19**, by ~1/4$^{th}$, when compared with those without vaccination.
- Those with primary vaccination and one booster have lowered their chance of dying of **COVID-19**, by ~1/6$^{th}$, when compared with those without vaccination.
- Those with primary vaccination and two boosters have lowered their chance of dying of **COVID-19**, by ~1/10$^{th}$, when compared with those without vaccination.

| | deaths | % of all deaths | cases | population 7/22/22 | % | CASES/ Population (7/22) | DEATHS/ Population (7/22) | deaths/ million/ week | % |
|---|---|---|---|---|---|---|---|---|---|
| unvaccinated | 6,559 | 38% | 707,447 | 8,902,210.65 | 12% | 7.9% | 0.074% | 41 | 63% |
| vaccinated | 3,906 | 23% | 511,859 | 22,898,297 | 31% | 2.2% | 0.017% | 9 | 15% |
| one booster | 5,755 | 34% | 1,129,984 | 29,289,472 | 40% | 3.9% | 0.012% | 11 | 17% |
| two boosters | 892 | 5% | 244,979 | 12,667,677 | 17% | 1.9% | 0.007% | 4 | 6% |
| Total | 17,112 | 100% | 2,594,269 | 73,757,657 | 100% | 3.5% | 0.117% | 65 | 100% |

APPENDIX-TABLE VI

There were 41 deaths per million Americans per week among the unvaccinated, but only 4 deaths per million Americans per week among those who had had 2 boosters. Of course, the additional boosters now being offered rase the possibility of further savings in life. Clearly, we shall be watching the data to see whether these hopeful possibilities are realized. However, the lesson is clear: It's the unvaccinated who are dying now, as are those who have not had all of the available boosters. For those who are most fully boosted, **COVID-19** death is becoming a memory.

Unfortunately, it is not such an encouraging story for **COVID-19** infection. As can be seen in FIGURES throughout this APPENDIX, for those who are fully boosted, the chance of death, ***Gompertzian Lethality***, may be drifting into the past, but the chance of becoming infected, ***Pasteurian Infectivity***, is not.

Benefits of higher vaccination compliance and additional ***Vaccination Opportunities***.

With these values derived from ***Gompertzian Analysis*** of actual data, we can derive plausible estimates of how death would have been reduced, and can be reduced in the future, with greater use of vaccination (FIGURE 5, APPENDIX-FIGURE-46, APPENDIX-TABLE V). Even without taking into account any vaccine driven reduction in infections, that is, in **COVID-19 *Infectivity*** (Cases/Population), these estimates make clear that many lives could be saved simply by wider adoption of the currently approved ***Vaccination Opportunities***, and that even more lives might be saved by additional ***Vaccination Opportunities***. The practical execution of these calculations were made by taking the actual numbers of cases and deaths, sorted by vaccination status, as assembled by the CDC for a sample of the USA population in 2022 (APPENDIX-TABLE V), and then replacing the values in cells for which we wish to examine the death rates had these groups adopted a higher level of vaccine utilization (grey cells in APPENDIX-TABLE V). These calculations reveal that had all Americans utilized at least primary vaccination, the reduction in **COVID-19 *Lethality*** (Deaths/Cases) would have resulted in the ~267,000 **COVID-19** deaths that occurred in 2022 being cut by ~13% to ~232,00 deaths. Had all Americans utilized at least primary vaccination and 1 booster, then **COVID-19** deaths that occurred in 2022 would have been cut by ~29% to ~189,00. Had all Americans utilized at least primary vaccination and 2 boosters, then death would have been cut by ~52% to ~129,00. Had all Americans utilized primary vaccination and 3 boosters, then lives lost would likely have been cut by ~68% to ~85,00 deaths. A total of 6 boosters yields a value of a ~91% reduction in lives lost, to ~25,000 deaths, far fewer lives than are lost each year in automobile accidents.

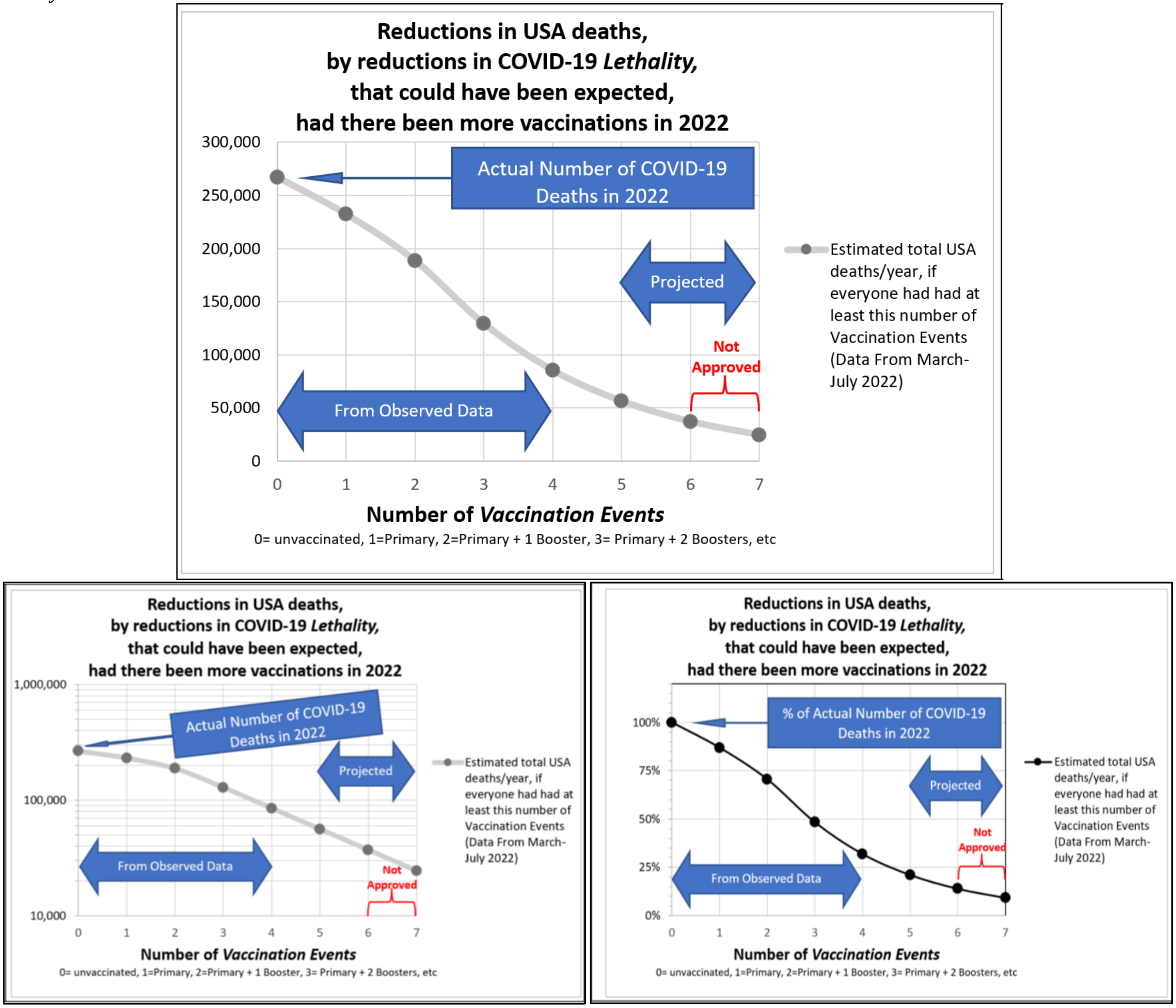

APPENDIX-FIGURE-46

| | deaths | % of all deaths | cases | population 7/22/22 | % | CASES/ Population (7/22) | DEATHS/ Population (7/22) | deaths/ million/ week | % | deaths | deaths, if they had had at least primary vac | deaths, if they had had at least primary vac + 1 Booster | deaths, if they had had at least primary vac + 2 Boosters | Projected deaths, if they had had at least primary vac + 3 Boosters | Projected deaths, if they had had at least primary vac + 4 Boosters | Projected deaths, if they had had at least primary vac + 5 Boosters | Projected deaths, if they had had at least primary vac + 6 Boosters | | % deaths, if they had had at least primary vac | %deaths, if they had had at least primary vac + 1 Booster | % deaths, if they had had at least primary vac + 2 Boosters | % Projected deaths, if they had had at least primary vac + 3 Boosters | % Projected deaths, if they had had at least primary vac + 4 Boosters | % Projected deaths, if they had had at least primary vac + 5 Boosters | % Projected deaths, if they had had at least primary vac + 6 Boosters |
|---|---|---|---|---|---|---|---|---|---|---|---|---|---|---|---|---|---|---|---|---|---|---|---|---|---|
| unvaccinated | 6,559 | 38% | 707,447 | 8,902,210.65 | 12% | 7.90% | 0.07% | 41 | 63% | 6,559 | 4,329 | 2,857 | 1,886 | 1,245 | 821 | 542 | 358 | | 66% | 44% | 29% | 19% | 13% | 8% | 5% |
| vaccinated | 3,906 | 23% | 511,859 | 22,898,297 | 31% | 2.20% | 0.02% | 9 | 15% | 3,906 | 3,906 | 2,578 | 1,701 | 1,123 | 741 | 489 | 323 | | 100% | 66% | 44% | 29% | 19% | 13% | 8% |
| one booster | 5,755 | 34% | 1,129,984 | 29,289,472 | 40% | 3.90% | 0.02% | 11 | 17% | 5,755 | 5,755 | 5,755 | 3,798 | 2,507 | 1,655 | 1,092 | 721 | | 100% | 100% | 66% | 44% | 29% | 19% | 13% |
| two boosters | 892 | 5% | 244,979 | 12,667,677 | 17% | 1.90% | 0.01% | 4 | 6% | 892 | 892 | 892 | 892 | 589 | 389 | 256 | 169 | | 100% | 100% | 100% | 66% | 44% | 29% | 19% |
| Total | 17,112 | 100% | 2,594,269 | 73,757,657 | 100% | 3.50% | 0.02% | 65 | 100% | 17,112 | 14,882 | 12,082 | 8,277 | 5,463 | 3,606 | 2,380 | 1,571 | | 87% | 71% | 48% | 32% | 21% | 14% | 9% |
| | | | | | | | | | | 0 | 1 | 2 | 3 | 4 | 5 | 6 | 7 | 0 | 1 | 2 | 3 | 4 | 5 | 6 | 7 |
| | | | | | | | | | Estimated Total USA deaths / year -> | 267,000 | 232,204 | 188,518 | 129,154 | 85,241 | 56,259 | 37,131 | 24,507 | 100% | 87% | 71% | 48% | 32% | 21% | 14% | 9% |

| | deaths | % of all deaths | cases | population 7/22/22 | % | CASES/ Population (7/22) | DEATHS/ Population (7/22) | deaths/ million/ week | % |
|---|---|---|---|---|---|---|---|---|---|
| unvaccinated | 6,559 | 38% | 707,447 | 8,902,210.65 | 12% | 7.90% | 0.07% | 41 | 63% |
| vaccinated | 3,906 | 23% | 511,859 | 22,898,297 | 31% | 2.20% | 0.02% | 9 | 15% |
| one booster | 5,755 | 34% | 1,129,984 | 29,289,472 | 40% | 3.90% | 0.02% | 11 | 17% |
| two boosters | 892 | 5% | 244,979 | 12,667,677 | 17% | 1.90% | 0.01% | 4 | 6% |
| Total | 17,112 | 100% | 2,594,269 | 73,757,657 | 100% | 3.50% | 0.02% | 65 | 100% |
| | | | | | | | | | Estimated Total USA deaths / year -> |

| | deaths | deaths, if they had had at least primary vac | deaths, if they had had at least primary vac + 1 Booster | deaths, if they had had at least primary vac + 2 Boosters | Projected deaths, if they had had at least primary vac + 3 Boosters | Projected deaths, if they had had at least primary vac + 4 Boosters | Projected deaths, if they had had at least primary vac + 5 Boosters | Projected deaths, if they had had at least primary vac + 6 Boosters |
|---|---|---|---|---|---|---|---|---|
| unvaccinated | 6,559 | 4,329 | 2,857 | 1,886 | 1,245 | 821 | 542 | 358 |
| vaccinated | 3,906 | 3,906 | 2,578 | 1,701 | 1,123 | 741 | 489 | 323 |
| one booster | 5,755 | 5,755 | 5,755 | 3,798 | 2,507 | 1,655 | 1,092 | 721 |
| two boosters | 892 | 892 | 892 | 892 | 589 | 389 | 256 | 169 |
| Total | 17,112 | 14,882 | 12,082 | 8,277 | 5,463 | 3,606 | 2,380 | 1,571 |
| | 0 | 1 | 2 | 3 | 4 | 5 | 6 | 7 |
| Estimated Total USA deaths / year -> | 267,000 | 232,204 | 188,518 | 129,154 | 85,241 | 56,259 | 37,131 | 24,507 |

| | | % deaths, if they had had at least primary vac | %deaths, if they had had at least primary vac + 1 Booster | % deaths, if they had had at least primary vac + 2 Boosters | % Projected deaths, if they had had at least primary vac + 3 Boosters | % Projected deaths, if they had had at least primary vac + 4 Boosters | % Projected deaths, if they had had at least primary vac + 5 Boosters | % Projected deaths, if they had had at least primary vac + 6 Boosters |
|---|---|---|---|---|---|---|---|---|
| unvaccinated | | 66% | 44% | 29% | 19% | 13% | 8% | 5% |
| vaccinated | | 100% | 66% | 44% | 29% | 19% | 13% | 8% |
| one booster | | 100% | 100% | 66% | 44% | 29% | 19% | 13% |
| two boosters | | 100% | 100% | 100% | 66% | 44% | 29% | 19% |
| Total | | 87% | 71% | 48% | 32% | 21% | 14% | 9% |
| | 0 | 1 | 2 | 3 | 4 | 5 | 6 | 7 |
| | 100% | 87% | 71% | 48% | 32% | 21% | 14% | 9% |

APPENDIX-TABLE V

***Gompertzian Analysis*** of the Impact of Non-Immunological Factors On **COVID-19** Outcome

Vaccines and other things change **COVID-19** ***Lethality*** by pushing up or down ***Gompertz Lines***

Many comorbidity factors have been associated with mortality generally, and **COVID-19** ***Lethality*** specifically. ***Gompertzian Analysis*** gives us some insights, and a single mathematical framework, for assessing, and integrating, the impact of such factors.

The impact of Sex on All-Cause Mortality

Women live longer than men. Census Bureau data, displayed on the ***Gompertz Plot*** shown in APPENDIX-FIGURE-47 reveals that this sex-associated survival is roughly captured by a difference in ***Gompertzian Height***, $G_H$, with a somewhat similar ***Gompertzian Slope***, $G_S$. That is, at every age, men have a greater chance of death than women. This gives the appearance of roughly parallel ***Gompertz Lines***. (The subtle departure and curviness from absolute linearity and parallelism that the observant reader will note in APPENDIX-FIGURE-47 is a topic of continued analysis by my colleagues and I, but, fortunately, this quality does not affect the rough generalizations we shall discuss here.)

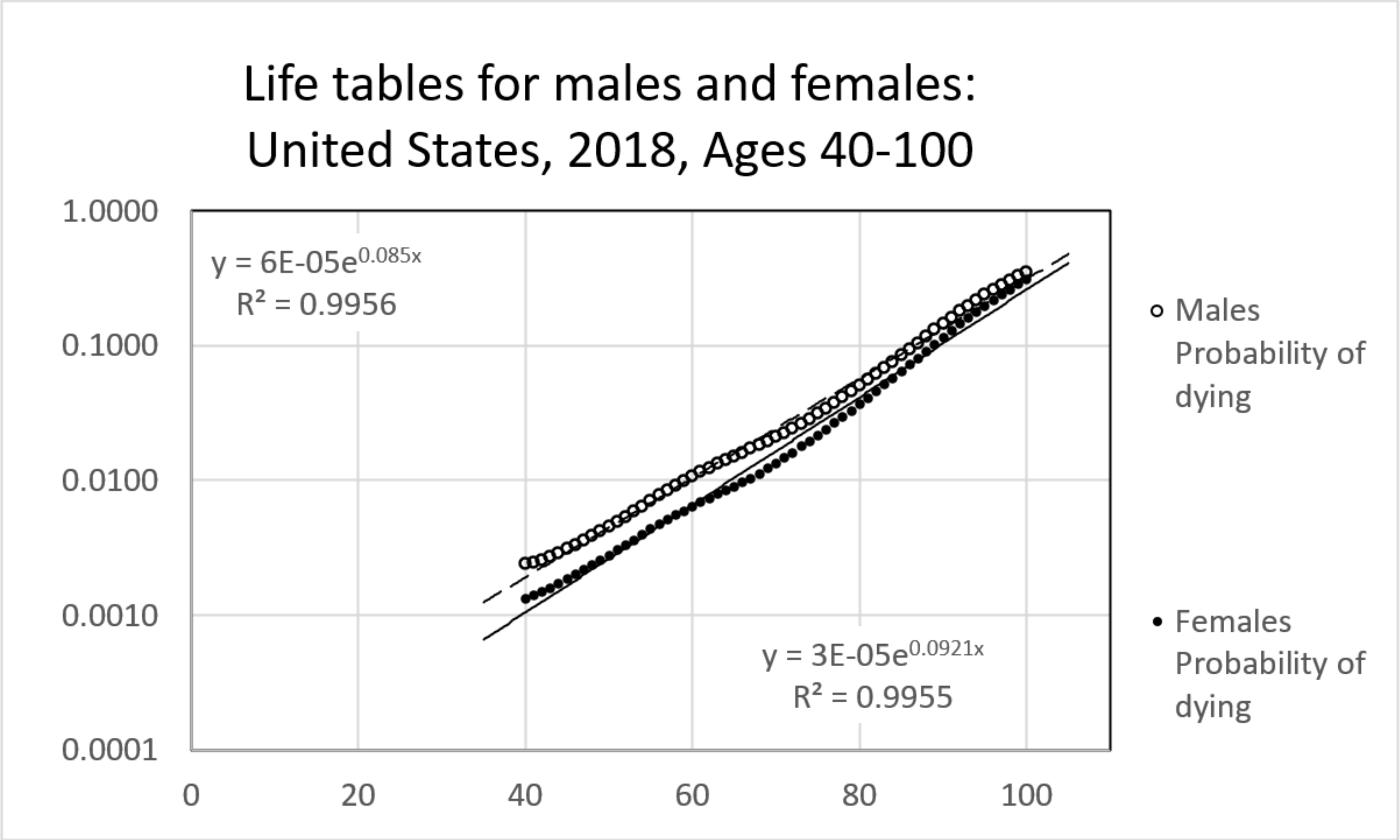

APPENDIX-FIGURE-47

The impact of Sex on **COVID-19 *Infectivity*** and ***Lethality***.

Women also have a lower chance of dying of **COVID-19** than men. A study from patients in Mexico provided age-sorted data on cases and death, with which it was possible to carry out ***Gompertzian Analysis*** of the impact of sex on **COVID 19** outcome (APPENDIX-FIGURE-48)[54]. These data revealed lower ***Gompertzian Lethality*** for women in comparison to men. The difference is captured, yet again, by a difference in ***Gompertzian Height***, $G_H$, with both sexes expressing the somewhat similar ***Gompertzian Slope***, $G_S$, as we have seen in so many instances above. That is, ***Gompertzian Analysis*** reveals that sex-associated differences in **COVID-19 *Gompertzian Lethality*** (Deaths/Cases) occurs similarly across all ages. There was no evident male/female difference in ***Pasteurian Infectivity*** (Cases/Population).

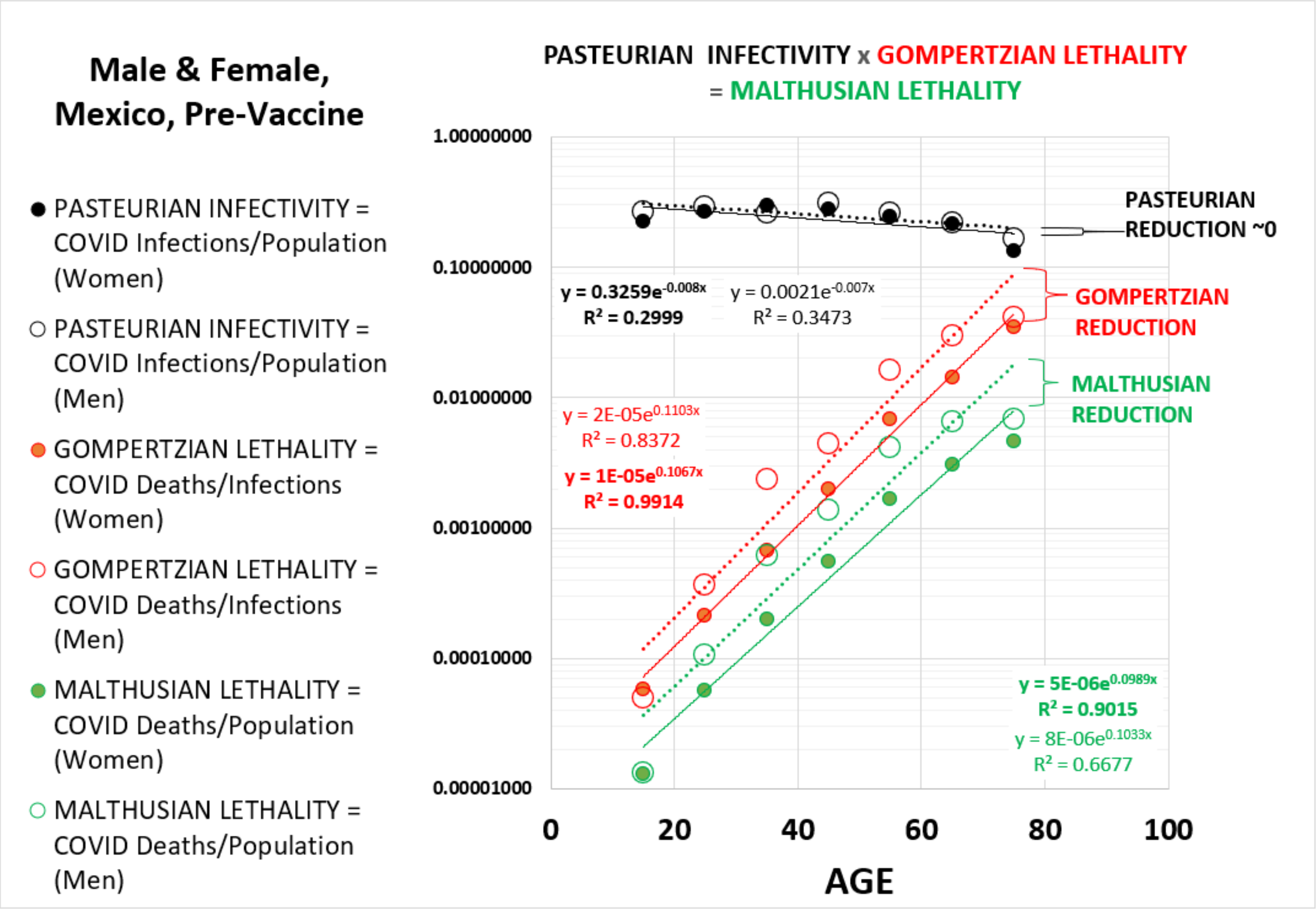

APPENDIX-FIGURE-48

The impact of Diabetes on **COVID-19** ***Infectivity*** and ***Lethality***.

Diabetes has also been known to increase the lethal burden of **COVID-19**. A study from patients in Sweden provided age-sorted data on cases and death, with which it was possible to carry out ***Gompertzian Analysis*** of the impact of diabetes on **COVID-19** outcome (APPENDIX-FIGURE-49).[55] These data revealed higher ***Gompertzian Lethality*** for diabetics in comparison to controls. The difference is captured, yet again, by a difference in ***Gompertzian Height***, $G_H$, with both diabetics and controls expressing the somewhat similar ***Gompertzian Slope***, $G_S$, as we have seen in so many instances above. There was also a strikingly evident diabetic/control difference in ***Pasteurian Infectivity*** (Cases/Population), with the two lines on the ***Gompertzian Plot*** being of a similar (but not identical) slope. That is, ***Gompertzian Analysis*** reveals that diabetes-associated differences in **COVID-19** ***Gompertzian Lethality*** (Deaths/Cases) and ***Pasteurian Infectivity*** (Cases/Population), both occurring across all ages, leading to a disturbing negative ***Malthusian Reduction*** in ***Malthusian Lethality*** (Deaths//Population).

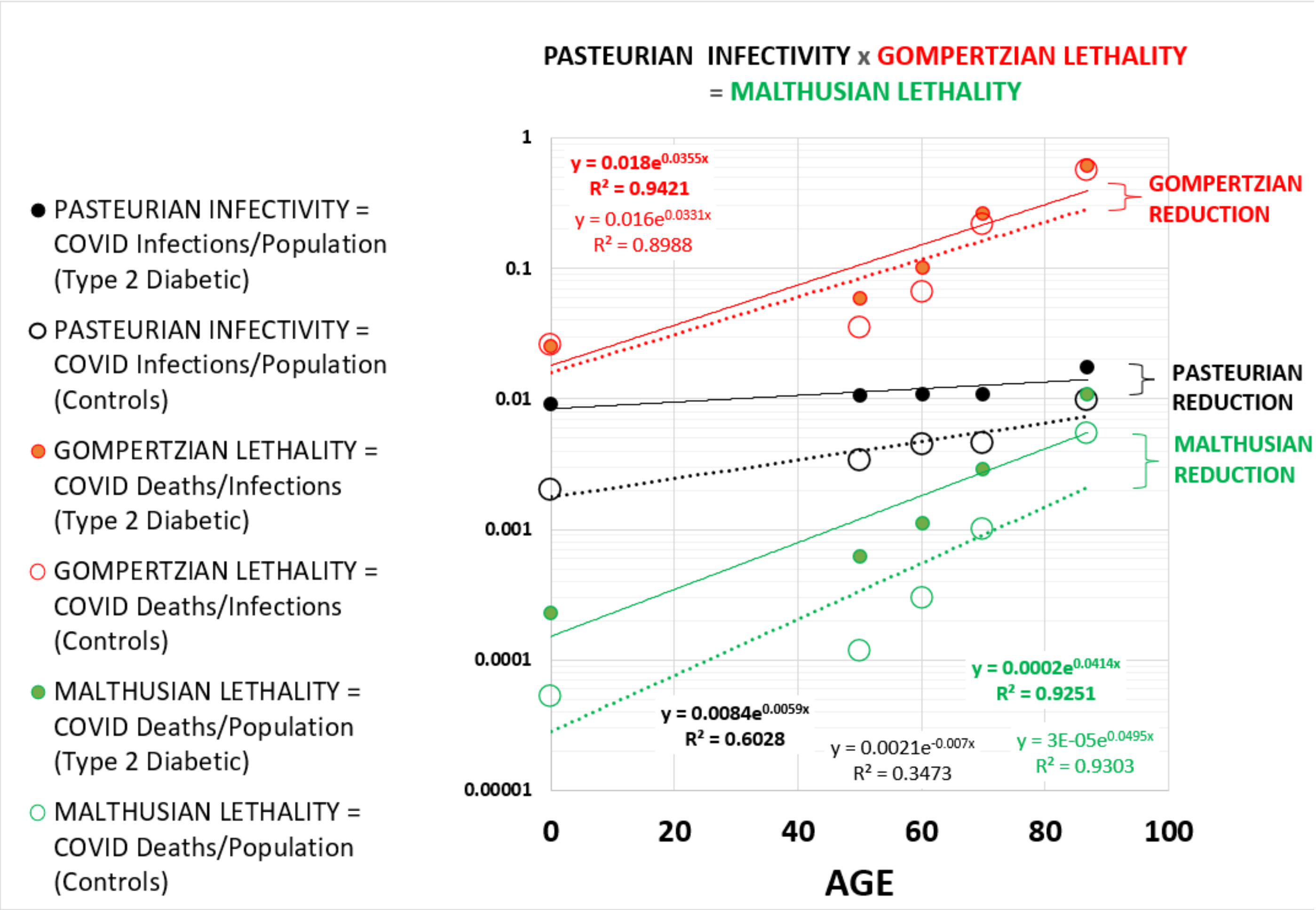

APPENDIX-FIGURE-49

***Gompertzian Lethality***, ***Pasteurian Infectivity***, and ***Malthusian Lethality*** and their negative ***Reductions*** associated with diabetes.

***<u>Gompertzian Analysis</u>*** <u>of LONG COVID</u>

**COVID**-19 manifests in multiple symptoms, which appear and last after infection,[56,57] as does pandemic associated excess mortality.[58,59,60,61] ***Gompertzian Analysis*** has found that unlike ***Gompertzian Lethality***, ***Long COVID*** occurs at relatively equivalent rates across ages, as reported in studies from Turkey[62] and UK[63] (APPENDIX-FIGURE-50), and elsewhere. Whether this age-equivalence occurs across each of the many Long-COVID symptoms is not clear and would be most worthy of a closer look.

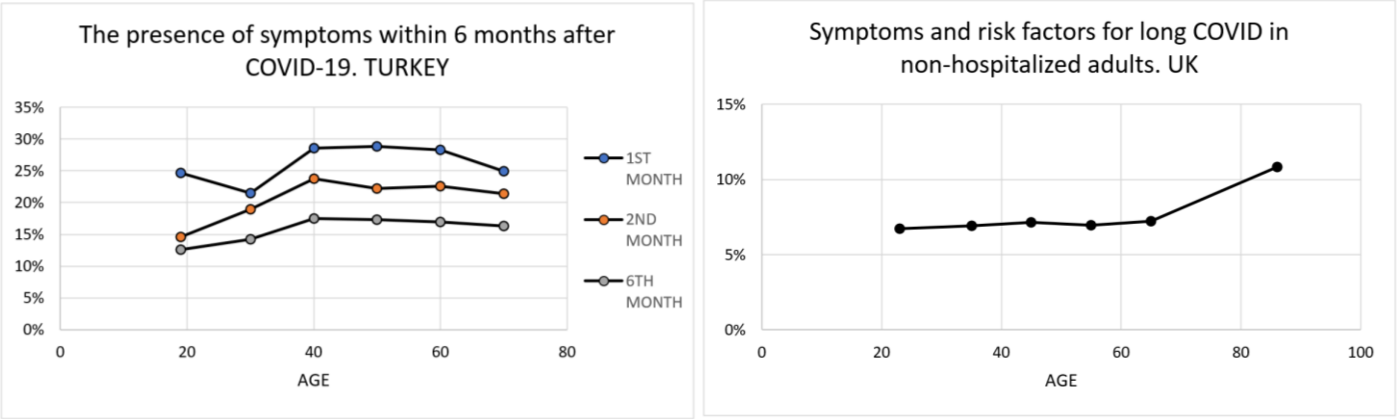

APPENDIX-FIGURE-50

Fortunately, vaccination has been found to protect against Long COVID,[64,6566,67], including several studies that have found progressive greater protected as the numbers of ***Vaccination Events*** (Primary & Boosters) are accumulated, as can be seen in one study whose values are shown APPENDIX-FIGURE-51[68] A large study of Long COVID in Scotland found vaccination reduced some Long COVID cases associated with 7 symptoms, although all were in fewer than 5% of Long COVID patients, and vaccine reduction was not reported of the most common symptoms: tiredness, headache, muscle aches/weakness, joint pain, breathless.[69] Al-Aly, Bowe, and Xie,[70] using USA VA data, found that vaccination (number of ***Vaccination Events*** not noted) reduced the risk of Long COVID, including increased risk death, 30 days after infection, but only partially. As they stated: "the findings suggest that vaccination before infection confers only partial protection in the post-acute phase of the disease; hence, reliance on it as a sole mitigation strategy may not optimally reduce long-term health consequences of SARS-CoV-2 infection." Clearly, whether additional boosters continue to reduce Long COVID, and precisely which symptoms/illnesses are reduced by vaccination, are topics most worthy of continued analysis.

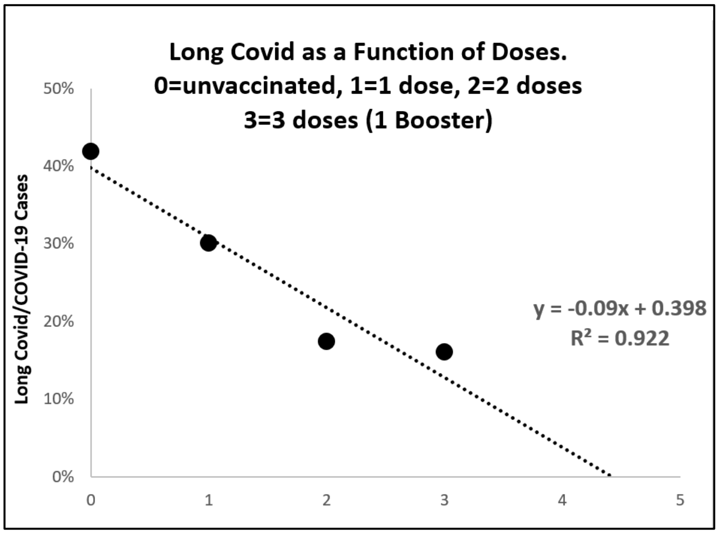

APPENDIX-FIGURE-51[50]

The impact of VARIANTS on **COVID-19 *Infectivity*** and ***Lethality***.

Unlike **COVID-19 *Pasteurian Infectivity***, new variants don't increase **COVID-19 *Gompertzian Lethality***. Two studies provided age-sorted data on cases and death, one in the UK,[71] and the other in the USA[72], with which it was possible to carry out ***Gompertzian Analysis*** of **COVID-19** variants. In both cases, ***delta*** was associated with lower ***Gompertzian Lethality*** than the initial variant, and ***omicron*** with and even lower level of ***Gompertzian Lethality*** (APPENDIX-FIGURES 48 AND 49). These differences are captured, yet again, by differences in ***Gompertzian Height***, $\boldsymbol{G_H}$, with each expressing the somewhat similar ***Gompertzian Slope***, $\boldsymbol{G_S}$, as we have seen in so many instances above. That is, ***Gompertzian Analysis*** reveals that the same variant-associated differences in **COVID-19 *Gompertzian Lethality*** (Deaths/Cases) occurs similarly across all ages.

Of course, the measures of variant-associated differences in **COVID-19 *Gompertzian Lethality*** (Deaths/Cases) come from separate points in time, when different levels vaccination, infection-acquire immunity, and other forces are likely to have been at work, in addition to the variant's properties themselves. Thus, the data shown here don't tell us which of these forces are at work, although this would certainly be a worthwhile exercise, and a perfectly achievable goal, should the underlying data be available. At the very least, however, we can see that the emergence of these new variants was not correlated with an increased chance of death after infection.

In contrast to the decline in **COVID-19 *Gompertzian Lethality*** (Deaths/Cases) with each new variant, **COVID-19 *Pasteurian Infectivity*** (Cases/Population) has increased with each new variant. Previous experience teaches that viral Darwinism would select for variants to evolve to be more infectious (**COVID-19 *Pasteurian Infectivity***), but not necessarily more deadly (**COVID-19 *Gompertzian Lethality***), and that is what appears to have been the case so far.

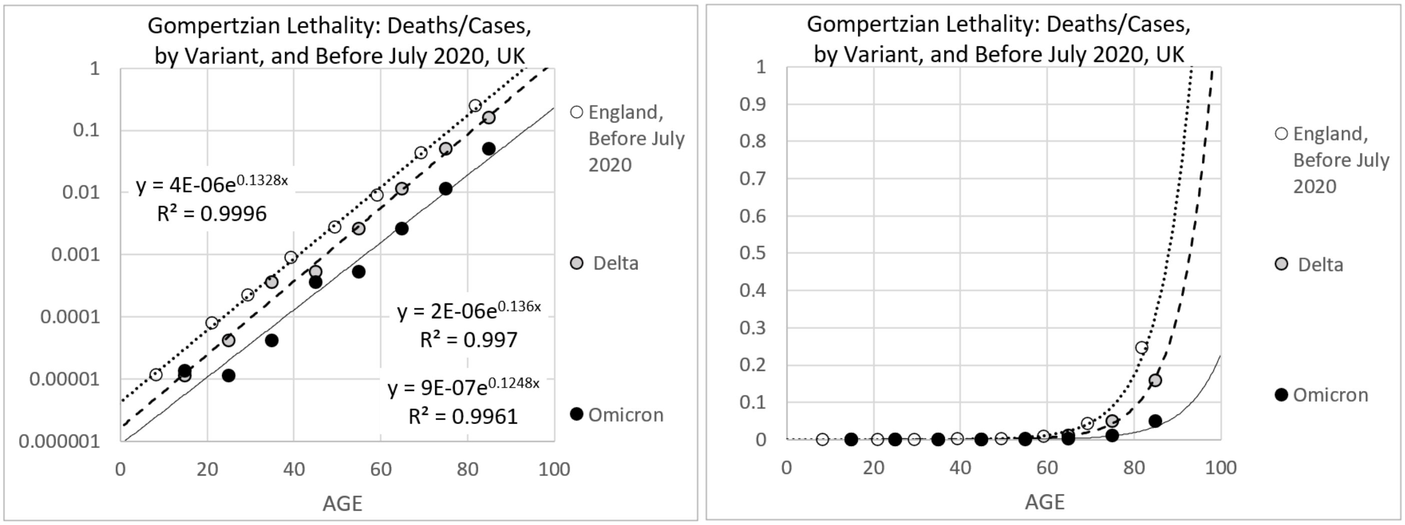

APPENDIX-FIGURE-52 **COVID-19 *Lethality*** (Deaths/Cases) By Variant, UK[71]

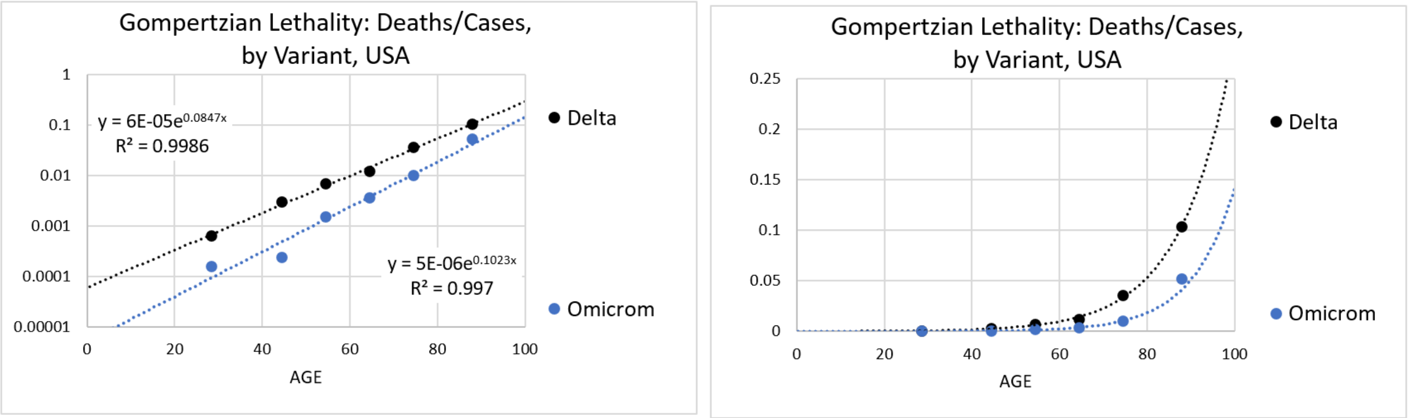

APPENDIX-FIGURE-53 **COVID-19 *Lethality*** (Deaths/Cases) By Variant, USA[72]

***Gompertzian Analysis*** can aid in the formation of **COVID-19** vaccination policy.

An examination of **COVID-19** vaccination policy statements, thoughtfully created by a number of national and international authorities, makes clear that a single approach for identifying which individuals, at which ages, should receive which vaccines, has yet to emerge.[73,74,75,76,77,78,79,80] ***Gompertzian Analysis*** thus gives a method for molding population-wide policy so as to achieve the greatest possible reduction in **COVID-19** deaths, as well as to create methods for tracking individual's accumulation of ***Vaccination Events***, to monitor the success of such efforts, and to modify guidance if policy does not achieve the goals expected.

15 Makeham, W. M. (1860). "On the Law of Mortality and the Construction of Annuity Tables". J. Inst. Actuaries and Assur. Mag. 8 (6): 301–310. doi:10.1017/S204616580000126X. JSTOR 41134925.

16 COVID-19 Forecasting Team. Variation in the COVID-19 infection-fatality ratio by age, time, and geography during the pre-vaccine era: a systematic analysis. Lancet. 2022 Apr 16;399(10334):1469-1488. doi: 10.1016/S0140-6736(21)02867-1. Epub 2022 Feb 24. Erratum in: Lancet. 2022 Apr 16;399(10334):1468. PMID: 35219376; PMCID: PMC8871594.

17 Citi L, Su, J, Huang, L, Michaelson, J. Counting Cells By Age Tells Us How, and Why, and When, We Grow, and Become Old and Ill. 2023. https://www.medrxiv.org/content/10.1101/2023.01.05.23284244v1.

18 Stadler T, Pybus OG, Stumpf MPH. Phylodynamics for cell biologists. Science. 2021 Jan 15;371(6526):eaah6266. doi: 10.1126/science.aah6266. PMID: 33446527.

19 Michaelson J. Cell selection in development. Biol Rev Camb Philos Soc. 1987 May;62(2):115-39. doi: 10.1111/j.1469-185x.1987.tb01264.x. PMID: 3300794.

20 Michaelson J. Cellular selection in the genesis of multicellular organization. Lab Invest. 1993 Aug;69(2):136-51. PMID: 8350596.

21 Michaelson J. The role of molecular discreteness in normal and cancerous growth. Anticancer Res. 1999 Nov-Dec;19(6A):4853-67. PMID: 10697599.

22 Cagan A, Baez-Ortega A, Brzozowska N, Abascal F, Coorens THH, Sanders MA, Lawson ARJ, Harvey LMR, Bhosle S, Jones D, Alcantara RE, Butler TM, Hooks Y, Roberts K, Anderson E, Lunn S, Flach E, Spiro S, Januszczak I, Wrigglesworth E, Jenkins H, Dallas T, Masters N, Perkins MW, Deaville R, Druce M, Bogeska R, Milsom MD, Neumann B, Gorman F, Constantino-Casas F, Peachey L, Bochynska D, Smith ESJ, Gerstung M, Campbell PJ, Murchison EP, Stratton MR, Martincorena I. Somatic mutation rates scale with lifespan across mammals. Nature. 2022 Apr;604(7906):517-524. doi: 10.1038/s41586-022-04618-z. Epub 2022 Apr 13. PMID: 35418684; PMCID: PMC9021023.

23 Gladyshev VN et al. Molecular damage in aging. Nature Aging. volume 1, pages 1096–1106 (2021)

24 Gladyshev VN. On the cause of aging and control of lifespan: heterogeneity leads to inevitable damage accumulation, causing aging; control of damage composition and rate of accumulation define lifespan. Bioessays. 2012 Nov;34(11):925-9. doi: 10.1002/bies.201200092. Epub 2012 Aug 23. PMID: 22915358; PMCID: PMC3804916.

25 Glynn JR, Moss PAH. Systematic analysis of infectious disease outcomes by age shows lowest severity in school-age children. Sci Data. 2020 Oct 15;7(1):329. doi: 10.1038/s41597-020-00668-y. PMID: 33057040; PMCID: PMC7566589.

26 Jones HB, A SPECIAL CONSIDERATION OF THE AGING PROCESS, DISEASE, AND LIFE EXPECTANCY November 1, 1955 CLEVELAND PUBLIC LIBRARY TECHNOLOGY DIVISION MAR 2 1956 SERIAL DOCS OARDS Printed for the U. S. Atomic Energy Commission UNIVERSITY OF CALIFORNIA Radiation Laboratory Berkeley, California Contract No. W - 7405 - eng – 48. https://www.google.com/books/edition/A_Special_Consideration_of_the_Aging_Pro/xLL4pko-4hoC?hl=en

27 Tartof SY, Qian L, Hong V, Wei R, Nadjafi RF, Fischer H, Li Z, Shaw SF, Caparosa SL, Nau CL, Saxena T, Rieg GK, Ackerson BK, Sharp AL, Skarbinski J, Naik TK, Murali SB. Obesity and Mortality Among Patients Diagnosed With COVID-19: Results From an Integrated Health Care Organization. Ann Intern Med. 2020 Nov 17;173(10):773-781. doi: 10.7326/M20-3742. Epub 2020 Aug 12. PMID: 32783686; PMCID: PMC7429998.

28 Scobie H. Update on Current Epidemiology of the COVID-19 Pandemic and SARS-CoV-2 Variants. https://www.fda.gov/media/164814/download 2022

29 L D Mueller, T J Nusbaum, M R Rose. The Gompertz equation as a predictive tool in demography. Exp Gerontol . 1995 Nov-Dec;30(6):553-69. doi: 10.1016/0531-5565(95)00029-1.

30 CDC COVID Data Tracker: Rates of COVID-19 Cases and Deaths by Vaccination Status
https://covid.cdc.gov/covid-data-tracker/#rates-by-vaccine-status
https://data.cdc.gov/Public-Health-Surveillance/Rates-of-COVID-19-Cases-or-Deaths-by-Age-Group-and/ukww-au2k

31 CDC COVID Data Tracker: Rates of COVID-19 Cases and Deaths by Vaccination Status
https://covid.cdc.gov/covid-data-tracker/#rates-by-vaccine-status
https://data.cdc.gov/Public-Health-Surveillance/Rates-of-COVID-19-Cases-or-Deaths-by-Age-Group-and/ukww-au2k

32 Ioannou GN, Locke ER, O'Hare AM, Bohnert ASB, Boyko EJ, Hynes DM, Berry K. COVID-19 Vaccination Effectiveness Against Infection or Death in a National U.S. Health Care System : A Target Trial Emulation Study. Ann Intern Med. 2022 Mar;175(3):352-361. doi: 10.7326/M21-3256. Epub 2021 Dec 21. PMID: 34928700; PMCID: PMC8697485.

33 Ioannou GN, Locke ER, Green PK, Berry K. Comparison of Moderna versus Pfizer-BioNTech COVID-19 vaccine outcomes: A target trial emulation study in the U.S. Veterans Affairs healthcare system. EClinicalMedicine. 2022 Mar 5;45:101326. doi: 10.1016/j.eclinm.2022.101326. PMID: 35261970; PMCID: PMC8896984.

34 Vokó Z, Kiss Z, Surján G, Surján O, Barcza Z, Pályi B, Formanek-Balku E, Molnár GA, Herczeg R, Gyenesei A, Miseta A, Kollár L, Wittmann I, Müller C, Kásler M. Nationwide effectiveness of five SARS-CoV-2 vaccines in Hungary-the HUN-VE study. Clin Microbiol Infect. 2022 Mar;28(3):398-404. doi: 10.1016/j.cmi.2021.11.011. Epub 2021 Nov 25. PMID: 34838783; PMCID: PMC8612758.

35 Kiss Z, Wittmann I, Polivka L, Surján G, Surján O, Barcza Z, Molnár GA, Nagy D, Müller V, Bogos K, Nagy P, Kenessey I, Wéber A, Pálosi M, Szlávik J, Schaff Z, Szekanecz Z, Müller C, Kásler M, Vokó Z. Nationwide Effectiveness of First and Second SARS-CoV2 Booster Vaccines During the Delta and Omicron Pandemic Waves in Hungary (HUN-VE 2 Study). Front Immunol. 2022 Jun 23;13:905585. doi: 10.3389/fimmu.2022.905585. PMID: 35812442; PMCID: PMC9260843.

36 Vokó Z, Kiss Z, Surján G, Surján O, Barcza Z, Wittmann I, Molnár GA, Nagy D, Müller V, Bogos K, Nagy P, Kenessey I, Wéber A, Polivka L, Pálosi M, Szlávik J, Rokszin G, Müller C, Szekanecz Z, Kásler M. Effectiveness and Waning of Protection With Different SARS-CoV-2 Primary and Booster Vaccines During the Delta Pandemic Wave in 2021 in Hungary (HUN-VE 3 Study). Front Immunol. 2022 Jul 22;13:919408. doi: 10.3389/fimmu.2022.919408. PMID: 35935993; PMCID: PMC9353007..

37 Rearte A, Castelli JM, Rearte R, Fuentes N, Pennini V, Pesce M, Barbeira PB, Iummato LE, Laurora M, Bartolomeu ML, Galligani G, Del Valle Juarez M, Giovacchini CM, Santoro A, Esperatti M, Tarragona S, Vizzotti C. Effectiveness of rAd26-rAd5, ChAdOx1 nCoV-19, and BBIBP-CorV vaccines for risk of infection with SARS-CoV-2 and death due to COVID-19 in people older than 60 years in Argentina: a test-negative, case-control, and retrospective longitudinal study. Lancet. 2022 Mar 26;399(10331):1254-1264. doi: 10.1016/S0140-6736(22)00011-3. Epub 2022 Mar 15. Erratum in: Lancet. 2022 Jun 11;399(10342):2190. PMID: 35303473; PMCID: PMC8923678.

38 Goldberg Y, Mandel M, Woodbridge Y, Fluss R, Novikov I, Yaari R, Ziv A, Freedman L, Huppert A. Similarity of Protection Conferred by Previous SARS-CoV-2 Infection and by BNT162b2 Vaccine: A 3-Month Nationwide Experience From Israel. Am J Epidemiol. 2022 Jul 23;191(8):1420-1428. doi: 10.1093/aje/kwac060. PMID: 35355048; PMCID: PMC8992290.

39 Goldberg Y, Mandel M, Woodbridge Y, Fluss R, Novikov I, Yaari R, Ziv A, Freedman L, Huppert A. Similarity of Protection Conferred by Previous SARS-CoV-2 Infection and by BNT162b2 Vaccine: A 3-Month Nationwide Experience From Israel. Am J Epidemiol. 2022 Jul 23;191(8):1420-1428. doi: 10.1093/aje/kwac060. PMID: 35355048; PMCID: PMC8992290.

40 Saban M, Myers V, Wilf-Miron R. Changes in infectivity, severity and vaccine effectiveness against delta COVID-19 variant ten months into the vaccination program: The Israeli case. Prev Med. 2022 Jan;154:106890. doi: 10.1016/j.ypmed.2021.106890. Epub 2021 Nov 17. PMID: 34800471; PMCID: PMC8596646.

41 Chava Peretz 1, Naama Rotem 2, Lital Keinan-Boker 3 4, Avner Furshpan 5, Manfred Green 3, Michal Bitan 6, David M Steinberg. Excess mortality in Israel associated with COVID-19 in 2020-2021 by age group and with estimates based on daily mortality patterns in 2000-2019 Int J Epidemiol. 2022 Jun 13;51(3):727-736. doi: 10.1093/ije/dyac047.

42 Ronen Arbel, Ruslan Sergienko, Michael Friger, Alon Peretz, Tanya Beckenstein, Shlomit Yaron, Doron Netzer, Ariel Hammerman Effectiveness of a second BNT162b2 booster vaccine against hospitalization and death from COVID-19 in adults aged over 60 years

Nat Med . 2022 Jul;28(7):1486-1490. doi: 10.1038/s41591-022-01832-0. Epub 2022 Apr 25.

43 Magen O, Waxman JG, Makov-Assif M, et al. Fourth dose of BNT162b2 mRNA Covid-19 vaccine in a nationwide setting. N Engl J Med 2022; 386: 1603-14 Epub 2022 Apr 13.

44 Yinon M Bar-On, Yair Goldberg, Micha Mandel, Omri Bodenheimer, Ofra Amir, Laurence Freedman, Sharon Alroy-Preis, Nachman Ash, Amit Huppert, Ron Milo Protection by a Fourth Dose of BNT162b2 against Omicron in Israel N Engl J Med . 2022 May 5;386(18):1712-1720. doi: 10.1056/NEJMoa2201570. Epub 2022 Apr 5.

45 Saban M, Myers V, Wilf-Miron R. Changes in infectivity, severity and vaccine effectiveness against delta COVID-19 variant ten months into the vaccination program: The Israeli case. Prev Med. 2022 Jan;154:106890. doi: 10.1016/j.ypmed.2021.106890. Epub 2021 Nov 17. PMID: 34800471; PMCID: PMC8596646.

46 CDC. Rates of COVID-19 Cases or Deaths by Age Group and Vaccination Status and Booster Dose; https://covid.cdc.gov/covid-data-tracker/#rates-by-vaccine-status; Updated 9/20/2022: Contact Dr Michaelson for dataset downloaded on that date; https://data.cdc.gov/Public-Health-Surveillance/Rates-of-COVID-19-Cases-or-Deaths-by-Age-Group-and/d6p8-wqjm

47 Johnson AG, Linde L, Ali AR, DeSantis A, Shi M, Adam C, Armstrong B, Armstrong B, Asbell M, Auche S, Bayoumi NS, Bingay B, Chasse M, Christofferson S, Cima M, Cueto K, Cunningham S, Delgadillo J, Dorabawila V, Drenzek C, Dupervil B, Durant T, Fleischauer A, Hamilton R, Harrington P, Hicks L, Hodis JD, Hoefer D, Horrocks S, Hoskins M, Husain S, Ingram LA, Jara A, Jones A, Kanishka FNU, Kaur R, Khan SI, Kirkendall S, Lauro P, Lyons S, Mansfield J, Markelz A, Masarik J 3rd, McCormick D, Mendoza E, Morris KJ, Omoike E, Patel K, Pike MA, Pilishvili T, Praetorius K, Reed IG, Severson RL, Sigalo N, Stanislawski E, Stich S, Tilakaratne BP, Turner KA, Wiedeman C, Zaldivar A, Silk BJ, Scobie HM. COVID-19 Incidence and Mortality Among Unvaccinated and Vaccinated Persons Aged ≥12 Years by Receipt of Bivalent Booster Doses and Time Since Vaccination - 24 U.S. Jurisdictions, October 3, 2021-December 24, 2022. MMWR Morb Mortal Wkly Rep. 2023 Feb 10;72(6):145-152. doi: 10.15585/mmwr.mm7206a3. PMID: 36757865; PMCID: PMC9925136.

48 Scobie H. Update on SARS-CoV-2 Variants and the Epidemiology of COVID-19. https://www.cdc.gov/vaccines/acip/meetings/downloads/slides-2022-09-01/02-covid-scobie-508.pdf

49 Muhsen K, Maimon N, Mizrahi AY, Boltyansky B, Bodenheimer O, Diamant ZH, Gaon L, Cohen D, Dagan R. Association of Receipt of the Fourth BNT162b2 Dose With Omicron Infection and COVID-19 Hospitalizations Among Residents of Long-term Care Facilities. JAMA Intern Med. 2022 Aug 1;182(8):859-867. doi: 10.1001/jamainternmed.2022.2658. PMID: 35737368; PMCID: PMC9227688

50 Nordström P, Ballin M, Nordström A. Effectiveness of a fourth dose of mRNA COVID-19 vaccine against all-cause mortality in long-term care facility residents and in the oldest old: A nationwide, retrospective cohort study in Sweden. Lancet Reg Health Eur. 2022 Oct;21:100466. doi: 10.1016/j.lanepe.2022.100466. Epub 2022 Jul 13. PMID: 35855494; PMCID: PMC9277096.

51 Grewal R, Kitchen SA, Nguyen L, Buchan SA, Wilson SE, Costa AP, Kwong JC. Effectiveness of a fourth dose of covid-19 mRNA vaccine against the omicron variant among long term care residents in Ontario, Canada: test negative design study. BMJ. 2022 Jul 6;378:e071502. doi: 10.1136/bmj-2022-071502. PMID: 35793826; PMCID: PMC9257064.

52 Tan CY, Chiew CJ, Lee VJ, Ong B, Lye DC, Tan KB. Effectiveness of a Fourth Dose of COVID-19 mRNA Vaccine Against Omicron Variant Among Elderly People in Singapore. Ann Intern Med. 2022 Nov;175(11):1622-1623. doi: 10.7326/M22-2042. Epub 2022 Sep 13. PMID: 36095316; PMCID: PMC9578545.

53 McConeghy KW, White EM, Blackman C, Santostefano CM, Lee Y, Rudolph JL, Canaday D, Zullo AR, Jernigan JA, Pilishvili T, Mor V, Gravenstein S. Effectiveness of a Second COVID-19 Vaccine Booster Dose Against Infection, Hospitalization, or Death Among Nursing Home Residents - 19 States, March 29-July 25, 2022. MMWR Morb Mortal Wkly Rep. 2022 Sep 30;71(39):1235-1238. doi: 10.15585/mmwr.mm7139a2. PMID: 36173757; PMCID: PMC9533729.

54 Torres-Ibarra L, Basto-Abreu A, Carnalla M, Torres-Alvarez R, Reyes-Sanchez F, Hernández-Ávila JE, Palacio-Mejia LS, Alpuche-Aranda C, Shamah-Levy T, Rivera JA, Barrientos-Gutierrez T. SARS-CoV-2 infection fatality rate after the first epidemic wave in Mexico. Int J Epidemiol. 2022 May 9;51(2):429-439. doi: 10.1093/ije/dyac015. PMID: 35157072; PMCID: PMC8903396.

55 Rawshani A, Kjölhede EA, Rawshani A, Sattar N, Eeg-Olofsson K, Adiels M, Ludvigsson J, Lindh M, Gisslén M, Hagberg E, Lappas G, Eliasson B, Rosengren A. Severe COVID-19 in people with type 1 and type 2 diabetes in Sweden: A nationwide retrospective cohort study. Lancet Reg Health Eur. 2021 May;4:100105. doi: 10.1016/j.lanepe.2021.100105. Epub 2021 Apr 30. PMID: 33969336; PMCID: PMC8086507.

56 Global Burden of Disease Long COVID Collaborators, Wulf Hanson S, Abbafati C, Aerts JG, Al-Aly Z, Ashbaugh C, Ballouz T, Blyuss O, Bobkova P, Bonsel G, Borzakova S, Buonsenso D, Butnaru D, Carter A, Chu H, De Rose C, Diab MM, Ekbom E, El Tantawi M, Fomin V, Frithiof R, Gamirova A, Glybochko PV, Haagsma JA, Haghjooy Javanmard S, Hamilton EB, Harris G, Heijenbrok-Kal MH, Helbok R, Hellemons ME, Hillus D, Huijts SM, Hultström M, Jassat W, Kurth F, Larsson IM, Lipcsey M, Liu C, Loflin CD, Malinovschi A, Mao W, Mazankova L, McCulloch D, Menges D, Mohammadifard N, Munblit D, Nekliudov NA, Ogbuoji O, Osmanov IM, Peñalvo JL, Petersen MS, Puhan MA, Rahman M, Rass V, Reinig N, Ribbers GM, Ricchiuto A, Rubertsson S, Samitova E, Sarrafzadegan N, Shikhaleva A, Simpson KE, Sinatti D, Soriano JB, Spiridonova E, Steinbeis F, Svistunov AA, Valentini P, van de Water BJ, van den Berg-Emons R, Wallin E, Witzenrath M, Wu Y, Xu H, Zoller T, Adolph C, Albright J, Amlag JO, Aravkin AY, Bang-Jensen BL, Bisignano C, Castellano R, Castro E, Chakrabarti S, Collins JK, Dai X, Daoud F, Dapper C, Deen A, Duncan BB, Erickson M, Ewald SB, Ferrari AJ, Flaxman AD, Fullman N, Gamkrelidze A, Giles JR, Guo G, Hay SI, He J, Helak M, Hulland EN, Kereselidze M, Krohn KJ, Lazzar-Atwood A, Lindstrom A, Lozano R, Malta DC, Månsson J, Mantilla Herrera AM, Mokdad AH, Monasta L, Nomura S, Pasovic M, Pigott DM, Reiner RC Jr, Reinke G, Ribeiro ALP, Santomauro DF, Sholokhov A, Spurlock EE, Walcott R, Walker A, Wiysonge CS, Zheng P, Bettger JP, Murray CJL, Vos T. Estimated Global Proportions of Individuals With Persistent Fatigue, Cognitive, and Respiratory Symptom Clusters Following Symptomatic COVID-19 in 2020 and 2021. JAMA. 2022 Oct 25;328(16):1604-1615. doi: 10.1001/jama.2022.18931. PMID: 36215063; PMCID: PMC9552043..

57 Al-Aly Z, Bowe B, Xie Y. Long COVID after breakthrough SARS-CoV-2 infection. Nat Med. 2022 Jul;28(7):1461-1467. doi: 10.1038/s41591-022-01840-0. Epub 2022 May 25. PMID: 35614233; PMCID: PMC9307472..

58 https://data.gov.scot/coronavirus-covid-19/detail.html#excess_deaths

59 Paglino E, Lundberg DJ, Cho A, Wasserman JA, Raquib R, Luck AN, Hempstead K, Bor J, Elo IT, Preston SH, Stokes AC. Excess all-cause mortality across counties in the United States, March 2020 to December 2021. medRxiv [Preprint]. 2022 May 17:2022.04.23.22274192. doi: 10.1101/2022.04.23.22274192. PMID: 35547848; PMCID: PMC9094106. https://www.ncbi.nlm.nih.gov/pmc/articles/PMC9094106/

60 Excess Deaths Associated with COVID-19 https://www.cdc.gov/nchs/nvss/vsrr/covid19/excess_deaths.htm

61 Excess mortality in England analysis https://app.powerbi.com/view?r=eyJrIjoiYmUwNmFhMjYtNGZhYS00NDk2LWFlMTAtOTg0OGNhNmFiNGM0IiwidCI6ImVlNGUxNDk5LTRhMzUtNGIyZS1hZDQ3LTVmM2NmOWRlODY2NiIsImMiOjh9

62 Emecen AN, Keskin S, Turunc O, Suner AF, Siyve N, Basoglu Sensoy E, Dinc F, Kilinc O, Avkan Oguz V, Bayrak S, Unal B. The presence of symptoms within 6 months after COVID-19: a single-center longitudinal study. Ir J Med Sci. 2022 Jun 17:1–10. doi: 10.1007/s11845-022-03072-0. Epub ahead of print. PMID: 35715663; PMCID: PMC9205653.

63 Subramanian A, Nirantharakumar K, Hughes S, Myles P, Williams T, Gokhale KM, Taverner T, Chandan JS, Brown K, Simms-Williams N, Shah AD, Singh M, Kidy F, Okoth K, Hotham R, Bashir N, Cockburn N, Lee SI, Turner GM, Gkoutos GV, Aiyegbusi OL, McMullan C, Denniston AK, Sapey E, Lord JM, Wraith DC, Leggett E, Iles C, Marshall T, Price MJ, Marwaha S, Davies EH, Jackson LJ, Matthews KL, Camaradou J, Calvert M, Haroon S. Symptoms and risk factors for long COVID in non-hospitalized adults. Nat Med. 2022 Aug;28(8):1706-1714. doi: 10.1038/s41591-022-01909-w. Epub 2022 Jul 25. PMID: 35879616; PMCID: PMC9388369.

64 Gao P, Liu J, Liu M. Effect of COVID-19 Vaccines on Reducing the Risk of Long COVID in the Real World: A Systematic Review and Meta-Analysis. Int J Environ Res Public Health. 2022 Sep 29;19(19):12422. doi: 10.3390/ijerph191912422. PMID: 36231717; PMCID: PMC9566528.

65 Kuodi P, Gorelik Y, Zayyad H, Wertheim O, Wiegler KB, Abu Jabal K, Dror AA, Nazzal S, Glikman D, Edelstein M. Association between BNT162b2 vaccination and reported incidence of post-COVID-19 symptoms: cross-sectional study 2020-21, Israel. NPJ Vaccines. 2022 Aug 26;7(1):101. doi: 10.1038/s41541-022-00526-5. PMID: 36028498; PMCID: PMC9411827.

66 Ayoubkhani D, Bosworth ML, King S, Pouwels KB, Glickman M, Nafilyan V, Zaccardi F, Khunti K, Alwan NA, Walker AS. Risk of Long COVID in People Infected With Severe Acute Respiratory Syndrome Coronavirus 2 After 2 Doses of a Coronavirus Disease 2019 Vaccine: Community-Based, Matched Cohort Study. Open Forum Infect Dis. 2022 Sep 12;9(9):ofac464. doi: 10.1093/ofid/ofac464. PMID: 36168555; PMCID: PMC9494414.

67 Brannock MD, Chew RF, Preiss AJ, Hadley EC, McMurry JA, Leese PJ, Girvin AT, Crosskey M, Zhou AG, Moffitt RA, Funk MJ, Pfaff ER, Haendel MA, Chute CG; N3C and RECOVER Consortia. Long COVID Risk and Pre-COVID Vaccination: An EHR-Based

Cohort Study from the RECOVER Program. medRxiv [Preprint]. 2022 Oct 7:2022.10.06.22280795. doi: 10.1101/2022.10.06.22280795. PMID: 36238713; PMCID: PMC9558440.

68 Azzolini E, Levi R, Sarti R, Pozzi C, Mollura M, Mantovani A, Rescigno M. Association Between BNT162b2 Vaccination and Long COVID After Infections Not Requiring Hospitalization in Health Care Workers. JAMA. 2022 Aug 16;328(7):676-678. doi: 10.1001/jama.2022.11691. PMID: 35796131; PMCID: PMC9250078.

69 Hastie CE, Lowe DJ, McAuley A, Winter AJ, Mills NL, Black C, Scott JT, O'Donnell CA, Blane DN, Browne S, Ibbotson TR, Pell JP. Outcomes among confirmed cases and a matched comparison group in the Long-COVID in Scotland study. Nat Commun. 2022 Oct 12;13(1):5663. doi: 10.1038/s41467-022-33415-5. Erratum in: Nat Commun. 2022 Nov 1;13(1):6540. PMID: 36224173; PMCID: PMC9556711.

70 Al-Aly Z, Bowe B, Xie Y. Long COVID after breakthrough SARS-CoV-2 infection. Nat Med. 2022 Jul;28(7):1461-1467. doi: 10.1038/s41591-022-01840-0. Epub 2022 May 25. PMID: 35614233; PMCID: PMC9307472.

71 Nyberg T, Ferguson NM, Nash SG, Webster HH, Flaxman S, Andrews N, Hinsley W, Bernal JL, Kall M, Bhatt S, Blomquist P, Zaidi A, Volz E, Aziz NA, Harman K, Funk S, Abbott S; COVID-19 Genomics UK (COG-UK) consortium, Hope R, Charlett A, Chand M, Ghani AC, Seaman SR, Dabrera G, De Angelis D, Presanis AM, Thelwall S. Comparative analysis of the risks of hospitalisation and death associated with SARS-CoV-2 omicron (B.1.1.529) and delta (B.1.617.2) variants in England: a cohort study. Lancet. 2022 Apr 2;399(10332):1303-1312. doi: 10.1016/S0140-6736(22)00462-7. Epub 2022 Mar 16. PMID: 35305296; PMCID: PMC8926413.

72 Skarbinski J, Wood MS, Chervo TC, Schapiro JM, Elkin EP, Valice E, Amsden LB, Hsiao C, Quesenberry C, Corley DA, Kushi LH. Risk of severe clinical outcomes among persons with SARS-CoV-2 infection with differing levels of vaccination during widespread Omicron (B.1.1.529) and Delta (B.1.617.2) variant circulation in Northern California: A retrospective cohort study. Lancet Reg Health Am. 2022 Aug;12:100297. doi: 10.1016/j.lana.2022.100297. Epub 2022 Jun 16. PMID: 35756977; PMCID: PMC9212563.

[73] FDA. Coronavirus (COVID-19) Update: FDA Authorizes Second Booster Dose of Two COVID-19 Vaccines for Older and Immunocompromised Individuals. https://www.fda.gov/news-events/press-announcements/coronavirus-covid-19-update-fda-authorizes-second-booster-dose-two-covid-19-vaccines-older-and

[79] COVID-19 vaccine: Canadian Immunization Guide. https://www.canada.ca/en/public-health/services/publications/healthy-living/canadian-immunization-guide-part-4-active-vaccines/page-26-covid-19-vaccine.html#t3

[80] JCVI statement on spring 2023 COVID-19 vaccinations, 22 February 2023. Published 7 March 2023. https://www.gov.uk/government/publications/spring-2023-covid-19-vaccination-programme-jcvi-advice-22-february-2023/jcvi-statement-on-spring-2023-covid-19-vaccinations-22-february-2023
